# Who Publishes During Residency? A Systematic Review and Meta-Analysis of Peer-Reviewed Publication Among Residents in the United States

**DOI:** 10.64898/2026.10.29.26366831

**Authors:** Pei-Lun Chen, Wei-Lin Tsai, Wan-Chun Liao, I-Chen Tsai

## Abstract

**Background/Objectives:** The Accreditation Council for Graduate Medical Education requires US residents to participate in scholarship, but no review has pooled how many physician residents publish during training. We estimated the proportion who published at least 1 peer-reviewed article during residency and compared specialties.

**Methods:** In October 2026, we searched PubMed, Europe PMC, OpenAlex, ERIC, and ClinicalTrials.gov (protocol, osf.io/eupxr). AI agents screened records, two AI models from different developers independently extracted outcome data, and an author decided every disagreement against the source. Proportions were pooled with random-effects generalized linear mixed models.

**Results:** Of 172 included studies, 32 US studies (13,450 residents) reported the primary outcome. The pooled proportion was 59.6% (95% CI, 48.7%–69.6%). Surgical specialties had 68.7% and medical specialties 29.6%. Adjusted for data source and pre-intervention status, the odds of publishing in medical cohorts were less than one-third of those in surgical cohorts: odds ratio (OR), 0.28 (95% CI, 0.12–0.63).

Conclusions:

About 60% of US residents publish during training, but specialties differ widely, and medical specialties have the most room for improvement.

**Key Points:** *Question:* What proportion of US residents publish at least 1 peer-reviewed article during residency, and does it differ by specialty?

*Findings:* In 32 US studies with 13,450 residents, the pooled proportion was 59.6%: 68.7% in surgical and 29.6% in medical specialties. After adjustment for data source and pre-intervention status, the odds of publishing in medical cohorts were less than one-third of those in surgical cohorts (OR, 0.28).

*Meaning:* About 6 in 10 US residents publish during training, and medical specialties have the most room for improvement.

## 1. Introduction

The Accreditation Council for Graduate Medical Education (ACGME) requires residents to participate in scholarship [1]. Scholarship can take many forms, including quality improvement projects, case reports, and conference presentations. A peer-reviewed publication is only one of them, but it is the easiest to verify from outside a program. It has also passed review by independent peers and remains a permanent, searchable record of the work. It is also used when residents apply for fellowship positions [2,3]. Program directors, residents, and accreditors therefore have reason to ask a simple question. How many residents publish during training?

No single answer exists across specialties. Estimates are scattered across specialty-specific reports. Some count publications from program records [4,5], some search a bibliographic database such as PubMed for each resident [6,7], and some ask residents in surveys [8]. Definitions of the training window also differ. For a program director who wants to know whether a program performs well, no publicly available reference exists.

Earlier systematic reviews have explored this field. Hebert and colleagues reviewed resident research curricula [9]. Stevenson and colleagues reviewed 80 studies of interventions to increase scholarly productivity [10]. Wood and colleagues pooled the effects of scholarly activity initiatives, and Zimmerman and colleagues reviewed interventions to increase trainee publications [11,12]. These reviews focused on research curricula and changes in output associated with interventions. The proportion of physician residents who publish at least one article, pooled across specialties, is not yet available from this literature. Behal and colleagues pooled publication of pharmacy resident projects, with the project as the unit of analysis [13]. This review provides a pooled estimate for physician residents, with the resident as the unit.

We asked what proportion of residents in the United States (US) publish at least one peer-reviewed article during residency, and conversely how many finish training without one; how much this proportion differs between specialties once study design is taken into account; whether these differences reflect differences in training length; and whether the proportion has clearly risen over the past 40 years.

## 2. Materials and Methods

We conducted and report this review in accordance with the Preferred Reporting Items for Systematic Reviews and Meta-Analyses (PRISMA) 2020 statement [14] (Supplementary Material 1, Table S1). We registered the protocol on Open Science Framework (OSF) Registries on October 22, 2026, before screening began (https://osf.io/eupxr), and updated the registration before submission to match the methods reported here. Ethical review board approval and participant informed consent were not required.

### 2.1. Database Searches and Study Identification

We searched PubMed, Europe PMC, OpenAlex, the Education Resources Information Center (ERIC), and ClinicalTrials.gov using the following keywords, adapted to the search syntax of each database: (‘resident’ OR ‘residents’ OR ‘residency’ OR ‘house staff’ OR ‘trainee’) AND (‘publication’ OR ‘publish’ OR ‘scholarly activity’ OR ‘scholarly output’ OR ‘research productivity’ OR ‘research output’ OR ‘peer-reviewed’ OR ‘bibliometric’ OR ‘h-index’) AND (‘rate’ OR ‘proportion’ OR ‘percent’ OR ‘prevalence’ OR ‘number’ OR ‘per resident’). The search spanned the period from the first available entry in each database to the search date (October 22, 2026). The exact search strings, limits, and numbers of records for each source are given in Supplementary Material 1, Table S2.

In PubMed and Europe PMC, records indexed as editorials, letters, or comments were removed by publication-type filter; research letters reporting original data were eligible and were identified through the other databases and citation searching. We also searched the forward and backward citations of the included studies and of 4 earlier reviews [9–12] through OpenAlex.

Screening had three steps. First, two AI agents (Claude, Anthropic) independently screened titles and abstracts and then full texts against the eligibility criteria, recording the reason and source passage for each decision. A record went to full-text review if either agent judged it possibly eligible, and disagreements at full-text review were resolved against the source and confirmed by I.-C.T. Second, to check the exclusions, a model from a different developer (GPT-6 Astra, OpenAI), blinded to the original decisions, re-screened a random 10% of title-and-abstract exclusions and, within each exclusion reason, a random 20% of full-text exclusions, both from the database and citation searches; full-text exclusions were sampled more heavily because reports excluded at full text are closer to inclusion. I.-C.T. decided, against the source, every record on which the second model disagreed with the original decision. Third, I.-C.T. audited a random 10% of the title-and-abstract sample and a random 20% of the full-text sample against the source. This describes screening; data extraction followed a separate process (Section 2.7). Because the review focused on US residents, we limited the language to English.

### 2.2. Inclusion and Exclusion Criteria

The review question was defined by population, exposure, and outcome: (1) population: residents in any country, with the primary analysis restricted to categorical residents in ACGME-accredited US programs; (2) exposure: residency training, compared across specialties; and (3) outcome: publication of at least 1 peer-reviewed article during residency.

The inclusion criteria were: (1) studies that reported, or allowed calculation of, the number of residents with at least 1 peer-reviewed publication during training and the number assessed, or, for the rate outcome, total publications and resident-years; (2) cohorts of at least 10 residents; and (3) cross-sectional, cohort, database-search, or survey designs, including research letters that reported original data. We accepted any peer-reviewed item as each study defined it and recorded the types counted. Denominators restricted to residents who completed training are flagged in Supplementary Material 1, Table S3.

The minimum of 10 residents was set because proportions from very small cohorts are unstable [15]; a cohort of 5 residents, for example, can give proportions only in steps of 20%.

The exclusion criteria were: (1) studies whose outcome was not peer-reviewed publication, such as completed or presented projects; (2) samples selected by a later career outcome, such as residents who later entered fellowship or academic positions, because selection on later output inflates the proportion; (3) studies whose counting period extended into practice after training or covered all of medical school; (4) studies of medical students, fellows, or attending physicians only; (5) editorials, commentaries, correspondence without original data, and conference abstracts; and (6) repeated reports of the same cohort, of which we kept the report with the largest sample and most complete follow-up.

### 2.3. Cohort Selection and Classification

For intervention studies, we used only cohorts not exposed to the intervention. A cohort from the same program that was measured before its residents were exposed to the research intervention was flagged as a pre-intervention cohort. Comparison cohorts that did not receive the intervention were included without a flag: concurrent cohorts from other programs [16,17] and same-program residents who did not enroll in an optional research track [18]. Some comparison cohorts were selected; for example, one consisted of applicants whom the intervention program had ranked in the match [17]. For programs or reports with unusual designs, such as programs with long-standing research curricula, the authors decided how to include them under the same principles. Cohorts that might share a small number of residents with a larger cohort of the same specialty and graduation years, without clear evidence of overlap, were included.

We grouped specialties by a fixed rule. The surgical group comprised general surgery, orthopaedic surgery, neurological surgery, plastic surgery, otolaryngology, urology, ophthalmology, and obstetrics and gynecology. The medical group comprised internal medicine, family medicine, pediatrics, physical medicine and rehabilitation, neurology, and psychiatry. The third group, other specialties, comprised the specialties that are neither medical nor surgical: radiology, radiation oncology, pathology, anesthesiology, emergency medicine, and dermatology. Cohorts that mixed specialties entered only all-specialty analyses. Each specialty was assigned its standard ACGME training length, counted from the first postgraduate year and including any required preliminary year [19]. The unit of comparison was the residency that leads to initial board certification in each specialty; subspecialty fellowships after residency, such as cardiology or nephrology after internal medicine, were outside the scope of the review. Emergency medicine has 3– and 4-year programs, and each study was assigned the length of its stated format; a study that combined 3– and 4-year programs could not be assigned a single length and was not included in the training-length analysis.

We coded data source as database search (a search of each resident’s name in a bibliographic database such as PubMed or Scopus), program records (lists kept by the program, such as portfolios or annual reports), self-report, or mixed.

### 2.4. Methodological Quality Appraisal

We assessed risk of bias with the Joanna Briggs Institute (JBI) checklist for prevalence studies [20], adapted to emphasize the denominator, cohort completeness, coverage of the training window, and database verification of authorship. The two screening agents rated each study in the primary analysis independently, with the source passage for each item, and disagreements were resolved against the source and confirmed by I.-C.T.

### 2.5. Primary Outcome: Proportion of Residents With at Least 1 Peer-Reviewed Publication

The primary outcome was the proportion of residents with at least 1 peer-reviewed publication, as any author, during the training window, which ran from the start of residency to 12 months after graduation; the 12 months after graduation were included because resident research is often published more than a year after it is presented at an institutional research day [21]. Postgraduate year (PGY) refers to the year of graduate medical education training, and graduation means completion of the categorical residency. Dedicated research years taken between the start of residency and graduation fall inside this window, whether or not they count toward accredited training. We classified each study’s window as strict (matching this window), extended beyond this window, truncated (residents still in training), partial (only some training years), undefined, or unrestricted (the count was not limited to residency) (Supplementary Material 1, Table S4). Studies of advanced specialties often started counting at PGY-2, after a preliminary year elsewhere; we classified these windows as strict.

### 2.6. Secondary Outcomes

There were 2 secondary outcomes.

#### 2.6.1. Publications per Resident-Year, Any Author Position

The first secondary outcome was the number of peer-reviewed publications per resident-year, counting publications in any author position, as in the primary outcome. Studies that counted only first-author publications were not included in this outcome. When a study did not report resident-years, or did not state the length of training that its counting period covered, we calculated them from cohort size and the standard training length of the specialty.

#### 2.6.2. Residents With at Least 1 First-Author Publication

The second secondary outcome was the proportion of residents with at least 1 first-author publication. Studies that counted only first-author publications contributed to this outcome.

### 2.7. Data Extraction and Management

For the US studies, two AI models from different developers, Claude Opus 5.5 (Anthropic) and GPT-6 Astra (OpenAI), independently extracted the numbers assessed and published, first-author publications, total publications, resident-years, training window, data source, publication types, and pre-intervention status, following the same written instructions and recording the source location of each value. A third model, Claude Sonnet 5.5 (Anthropic), compared each disagreement with the source and proposed a value with the supporting passage, and I.-C.T. decided every disagreement. I.-C.T. also checked a random 10% of the fields on which both models agreed on a non-missing value against the source. Counts read from figures were checked against the reported means or percentages. Country, specialty, number of programs, graduation years, and protected research time, and all values for studies outside the US, were extracted by the two screening agents and checked in the same way. Each study contributed 1 cohort. When a study reported publication outcomes but not the number of residents who published or the number assessed, we emailed the corresponding author to request the data.

Supplementary Material 1, Table S5, records the source location and derivation of each value. AI tools also generated the R code for the statistical analyses and assisted with English editing of the manuscript; the authors reviewed and edited all code, output, and text.

### 2.8. Statistical Analyses

We used R version 4.5.3 (R Foundation for Statistical Computing) with the meta and metafor packages [22–24] and their default settings. Proportions were pooled with a random-effects generalized linear mixed model on the logit scale, and publication rates with a Poisson random-effects model [25,26]. We report pooled estimates with 95% confidence intervals (CIs), 95% prediction intervals, and the between-study variance (τ²) [27]. The primary analysis included all US studies except those that counted only part of the training window, counted beyond it, or did not limit the count to residency; it included pre-intervention cohorts.

Specialty groups were compared with a subgroup analysis. The medical–surgical difference was further examined with an exploratory meta-regression that included specialty group, data source, and pre-intervention status, and is reported as an odds ratio (OR) between study cohorts. For publications per resident-year, specialties were compared with a rate ratio. Change over time was examined with a random-effects meta-regression of the proportion on the midpoint of each cohort’s graduation years. Training length was examined with a linear model, alone and together with specialty; data source, window class, and number of institutions were examined with subgroup analyses or meta-regression; protected research time was reported descriptively. To describe the spread of published cohorts, we report the median, interquartile range, and 10th to 90th percentiles of the predictive distribution for each specialty group, without pre-intervention cohorts. A two-tailed *p* value below 0.05 was considered statistically significant.

### 2.9. Sensitivity Analyses

Sensitivity analyses repeated the primary analysis after restricting or adding studies: all countries; multi-institution studies only; without pre-intervention cohorts; studies that counted journal articles only; low risk of bias only; adding studies with extended, partial, or unrestricted windows, to examine how the definition of the counting window affects the result; without the program with a mandatory research curriculum, because the requirement may bias the estimate; adding first-author-only studies as a lower bound; without studies whose eligibility or extracted values had been adjudicated; and without smaller cohorts that might share a small number of residents with a larger cohort of the same specialty and graduation years, so that the same residents were not counted twice (double counting). For rates, we repeated the analysis without pre-intervention cohorts, without assumed resident-years, and without counts approximated from summary statistics. For the specialty gap, we refitted the adjusted model with *t*-tests and with data source collapsed to database search vs. other, and compared medical with surgical specialties, without adjustment, under 9 study selections and with leave-one-out analysis. The selections are nested subsets of the same studies, not independent replications.

## 3. Results

### 3.1. Study Identification and Characteristics

The PRISMA flow diagram and the numbers of records retrieved from each source are given in Supplementary Material 1, Figure S1 and Table S2. Searches of databases and registers identified 17,365 records: 16,912 from databases and 453 from ClinicalTrials.gov. After removing 7,533 duplicates and 27 records that PubMed counted but that could not be retrieved on download, we screened 9,805 records and excluded 9,028. Of 777 reports sought, 47 were not retrieved. We assessed 730 reports and excluded 580 (Supplementary Material 1, Table S11), most often because neither the number of residents who published nor the number assessed could be extracted (293 of 580, 50.5%). Four further reports lacked these numbers, but their corresponding authors provided them on request, and the studies were included [28–31] (Supplementary Material 1, Table S6). Citation searching identified 2,608 further records. Of these, 163 reports were assessed and 22 studies were included. In total, we included 172 studies (150 from databases and 22 from other methods). The characteristics of all included studies are given in Supplementary Material 1, Table S3.

In the audit of exclusions, the blinded second model agreed with 1,082 of 1,146 title-and-abstract exclusions (94.4%) and 143 of 145 full-text exclusions (98.6%). I.-C.T. upheld every exclusion in the audit samples (115 and 30). Of the 66 disagreements, I.-C.T. upheld 58 exclusions and sent 8 records to full-text review, where all 8 were excluded: 3 were not original research, 4 gave no extractable resident counts, and 1 did not measure peer-reviewed publication. After the audit, no excluded record had to be reclassified as included (title and abstract, 0 of 1,146; exact 95% CI upper bound, 0.3%; full text, 0 of 145; upper bound, 2.5%). For the 109 US studies, the two extraction models agreed on 830 of 981 extracted fields (84.6%), and I.-C.T. decided the other 151. In the random sample of 50 agreed fields (10% of 487), no error was found (0 of 50; exact 95% CI upper bound, 7.1%).

Thirty-two US studies with 13,450 residents reported the primary outcome. Of the 32 studies, 19 (59.4%) used database search, 4 (12.5%) program records, 5 (15.6%) self-report, and 4 (12.5%) mixed sources. Eighteen covered more than 1 institution, 13 covered 1, and 1 did not report the number. Five (15.6%) were pre-intervention cohorts, all from single programs. Their characteristics are given in Table 1.

**Table 1.** Characteristics of the 32 US studies in the primary analysis, by specialty group.

| Study | Ref. | Specialty | Programs, n | How publications were counted<sup>a</sup> | Window<sup>b</sup> | Residents, n | Published, n (%) |
|---|---|---|---|---|---|---|---|
| **Surgical specialties** |  |  |  |  |  |  |  |
| Thompson 2018 | 185 | Urology | 1 | Database search | Strict | 49 | 49 (100.0) |
| Warren 2025 | 193 | Urology | 42 | Database search | Strict | 228 | 211 (92.5) |
| Plackett 2020 | 150 | General surgery | 6 | Database search | Strict | 18 | 16 (88.9) |
| Donley 2024 | 79 | Orthopaedics | 138 | Database search | Strict | 758 | 629 (83.0) |
| Chen 2015 | 70 | Otolaryngology | 1 | Database search | Strict | 75 | 58 (77.3) |
| Huh 2022 | 108 | Ophthalmology | 10 | Program records | Strict | 237 | 165 (69.6) |
| Crowley 2012 | 73 | Neurosurgery | 87 | Database search | Strict | 717 | 498 (69.5) |
| Chopra 2025 | 6 | Orthopaedics | 208 | Database search | Truncated | 4,320 | 2,873 (66.5) |
| Kreitz 2019 | 123 | Orthopaedics | 13 | Self-report | Truncated | 274 | 178 (65.0) |
| Hang 2022 | 100 | Ophthalmology | NR | Database search | Truncated | 100 | 63 (63.0) |
| Reyes 2023 | 158 | Ophthalmology | 75 | Database search | Strict | 964 | 596 (61.8) |
| Daly 2006 | 5 | Otolaryngology | 1 | Program records | Strict | 36 | 22 (61.1) |
| Motiwala 2024<sup>c,d</sup> | 134 | Neurosurgery | 1 | Database search | Strict | 19 | 11 (57.9) |
| Choudhry 2023 | 7 | Ophthalmology | 110 | Database search | Truncated | 1,206 | 603 (50.0) |
| Thomas 2000<sup>c,d</sup> | 184 | General surgery | 1 | Mixed | Strict | 31 | 15 (48.4) |
| Patel 2018<sup>c</sup> | 146 | General surgery | 1 | Program records | Undefined | 36 | 16 (44.4) |
| Williams 2017 | 196 | Orthopaedics | 125 | Database search | Truncated | 1,690 | 733 (43.4) |
| Macknin 2014 | 129 | Orthopaedics | 1 | Database search | Strict | 74 | 30 (40.5) |
| **Medical specialties** |  |  |  |  |  |  |  |
| Stultz 2024 | 32 | Internal medicine | 1 | Database search | Strict | 522 | 430 (82.4) |
| Al-Shaikhly 2026 | 18 | Internal medicine | 1 | Database search | Strict | 39 | 18 (46.2) |
| Olanipekun 2023 | 139 | Internal medicine | 1 | Mixed | Truncated | 39 | 16 (41.0) |
| West 2011 | 16 | Internal medicine | NR | Database search | Strict | 430 | 155 (36.0) |
| Carter 2019<sup>c</sup> | 65 | Internal medicine | 1 | Mixed | Strict | 63 | 13 (20.6) |
| Taniguchi 1994 | 182 | PM&R | 60 | Self-report | Strict | 283 | 54 (19.1) |
| Abramson 2018 | 44 | Pediatrics | 22 | Self-report | Truncated | 464 | 62 (13.4) |
| Waheed 2025<sup>c</sup> | 40 | Family medicine | 1 | Program records | Strict | 14 | 0 (0.0) |
| **Other specialties** |  |  |  |  |  |  |  |
| Huang 2023 | 107 | Radiation oncology | NR | Database search | Strict | 20 | 18 (90.0) |
| Shi 2018 | 173 | Dermatology | NR | Database search | Strict | 392 | 327 (83.4) |
| Wilson 2005 | 8 | Radiation oncology | NR | Self-report | Truncated | 66 | 48 (72.7) |
| Sharpe 2025 | 170 | Anesthesiology | 3 | Database search | Strict | 149 | 88 (59.1) |
| Gottlieb 2021 | 94 | Emergency medicine | 1 | Mixed | Truncated | 23 | 13 (56.5) |
| Tsyrulnik 2026 | 4 | Emergency medicine | 8 | Self-report | Strict | 114 | 35 (30.7) |

<sup>a</sup> How the original study counted publications: database search, each resident's name searched in a bibliographic database such as PubMed or Scopus; program records, lists kept by the program; self-report, surveys of residents or programs; mixed, more than 1 of these.
<sup>b</sup> Strict, from the start of residency to 12 months after graduation; truncated, residents still in training; undefined, window not stated.
<sup>c</sup> Single-program pre-intervention cohort.
<sup>d</sup> Number with at least 1 publication read from a figure and checked against the reported mean or percentage. NR, not reported; PM&R, physical medicine and rehabilitation; Ref., reference number.

### 3.2. Methodological Quality of the Included Studies

Risk of bias was mixed. The sample frame was appropriate in 28 of 32 studies (87.5%), the outcome was measured validly in 21 (65.6%), and 6 (18.8%) clearly described the reliability of measurement (Supplementary Material 1, Figure S2 and Table S7).

### 3.3. Primary Outcome: About 6 in 10 Residents Published, Only 30% in Medical Specialties

The pooled proportion of residents who published at least 1 peer-reviewed article during residency was 59.6% (95% CI, 48.7%–69.6%; τ² = 1.51; Figure 1), with a prediction interval from 10.3% to 95.0%. Excluding the 5 pre-intervention cohorts, the estimate was 64.2% (27 studies; 95% CI, 53.4%–73.8%), close to the primary estimate, with largely overlapping 95% CIs.

**Figure 1.**
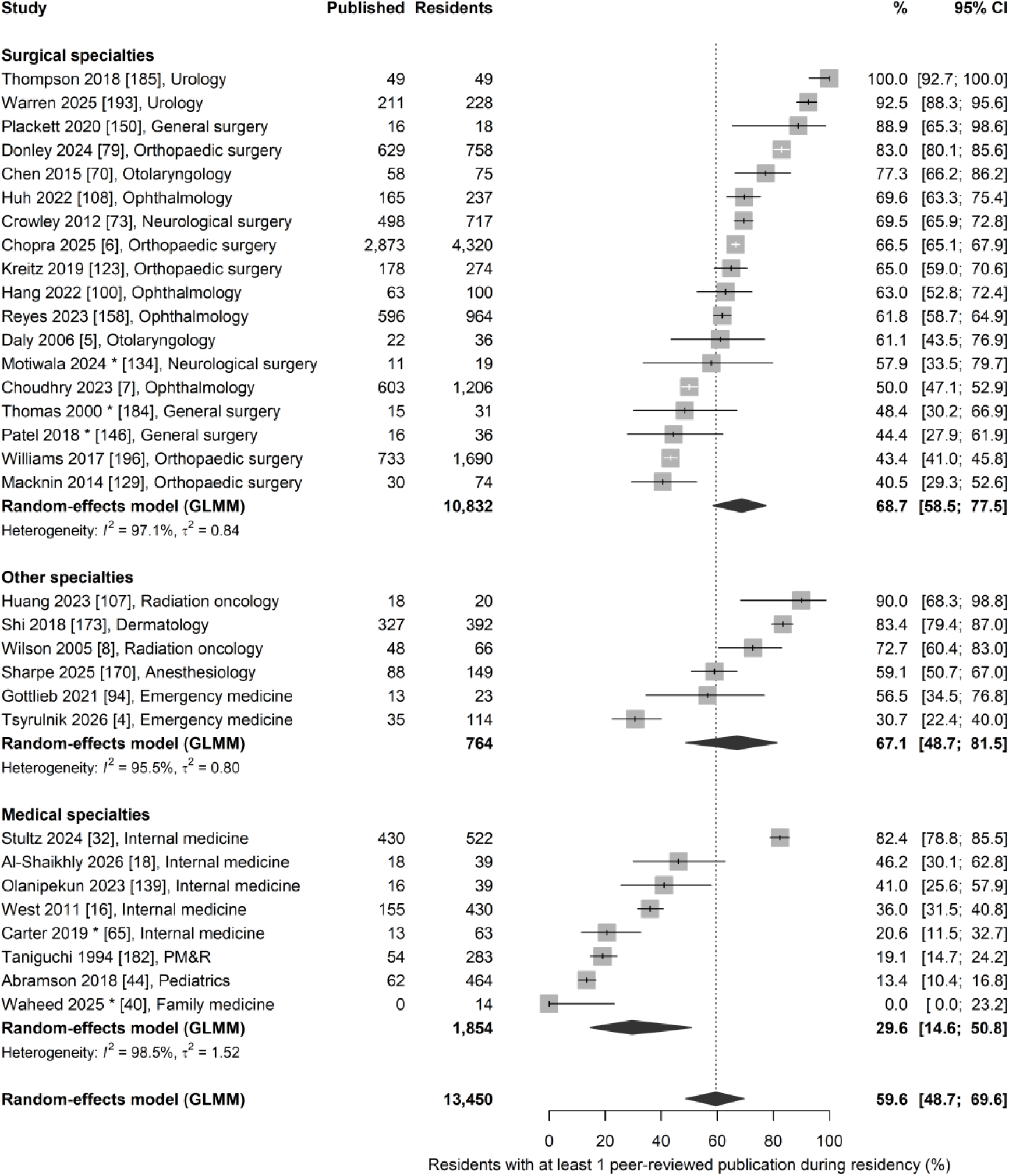
Proportion of residents with at least 1 peer-reviewed publication during residency in 32 US studies (13,450 residents), by specialty group, in studies published 1994–2026. Squares show study estimates and horizontal lines their 95% CIs. Diamonds show pooled estimates from random-effects generalized linear mixed models. The dotted vertical line marks the overall pooled estimate. Asterisks mark single-program pre-intervention cohorts, measured before a research intervention. Each group shows its heterogeneity statistics: I², the share of variation due to between-study differences, and τ², the between-study variance on the logit scale. Within each group, studies are sorted from highest to lowest proportion. Each label gives the first author, year, reference number in square brackets, and the specialty of the cohort. GLMM, generalized linear mixed model; PM&R, physical medicine and rehabilitation.

In the subgroup analysis (Figure 1, Table 2), proportions were 29.6% in medical specialties (8 studies), 67.1% in other specialties (6 studies), and 68.7% in surgical specialties (18 studies; omnibus test across the 3 groups, *p* < 0.001).

**Table 2.** Proportion of residents with at least 1 peer-reviewed publication during residency, by subgroup and sensitivity analysis. Primary analysis: 32 US studies (13,450 residents) published 1994– 2026.

| Analysis | Studies, n | Residents, n | Pooled proportion, % | 95% CI | 95% PI | τ<sup>2</sup> |
|---|---|---|---|---|---|---|
| **Primary analysis, United States** | 32 | 13,450 | 59.6 | 48.7–69.6 | 10.3–95.0 | 1.51 |
| **Specialty group**<sup>a</sup> |  |  |  |  |  |  |
| Surgical specialties | 18 | 10,832 | 68.7 | 58.5–77.5 | 23.0–94.2 | 0.84 |
| Other specialties<sup>b</sup> | 6 | 764 | 67.1 | 48.7–81.5 | 14.2–96.2 | 0.80 |
| Medical specialties | 8 | 1,854 | 29.6 | 14.6–50.8 | 1.8–90.4 | 1.52 |
| **Data source** |  |  |  |  |  |  |
| Database search<sup>c</sup> | 19 | 11,770 | 71.5 | 60.7–80.3 | 21.3–95.9 | 1.06 |
| Mixed<sup>d</sup> | 4 | 156 | 39.5 | 26.1–54.6 | 8.8–81.5 | 0.26 |
| Program records<sup>e</sup> | 4 | 323 | 38.8 | 10.7–76.9 | 0.3–99.4 | 2.28 |
| Self-report | 5 | 1,201 | 37.3 | 18.2–61.6 | 2.0–94.6 | 1.24 |
| **Sensitivity analyses** |  |  |  |  |  |  |
| All countries | 103 | 29,340 | 46.2 | 39.2–53.3 | 4.6–93.8 | 2.07 |
| US, multi-institution only | 18 | 12,410 | 60.6 | 47.9–72.0 | 12.6–94.2 | 1.19 |
| US, excluding pre-intervention cohorts | 27 | 13,287 | 64.2 | 53.4–73.8 | 13.8–95.3 | 1.33 |
| US, journal articles only<sup>f</sup> | 26 | 8,340 | 57.6 | 44.6–69.6 | 7.9–95.5 | 1.72 |
| US, low risk of bias only<sup>g</sup> | 12 | 2,740 | 73.8 | 58.2–85.1 | 16.1–97.6 | 1.36 |
| US, including extended-window studies | 39 | 14,644 | 56.5 | 44.9–67.5 | 6.3–96.2 | 2.08 |
| US, including partial-window studies | 40 | 14,393 | 51.0 | 38.0–63.9 | 3.4–96.9 | 2.74 |
| US, adding unrestricted-period study | 33 | 13,600 | 61.1 | 50.1–71.0 | 10.3–95.6 | 1.60 |
| US, excluding Mayo program<sup>h</sup> | 31 | 12,928 | 58.6 | 47.5–68.9 | 9.9–94.8 | 1.51 |
| US, adding first-author-only studies<sup>i</sup> | 36 | 16,056 | 60.1 | 50.6–69.0 | 12.4–94.1 | 1.32 |
| US, excluding studies whose eligibility or values were adjudicated<sup>j</sup> | 16 | 10,774 | 53.0 | 33.9–71.2 | 3.5–97.2 | 2.42 |
| US, excluding possibly overlapping smaller cohorts<sup>k</sup> | 30 | 12,939 | 59.0 | 47.3–69.8 | 9.1–95.4 | 1.64 |

Abbreviations: CI, confidence interval; PI, prediction interval; τ<sup>2</sup>, between-study variance on the logit scale.
<sup>a</sup> Subgroup τ<sup>2</sup> values come from separate random-effects models within each subgroup.
<sup>b</sup> Radiology, radiation oncology, pathology, anesthesiology, emergency medicine, and dermatology; the primary analysis contained 2 radiation oncology, 1 anesthesiology, 2 emergency medicine, and 1 dermatology study.
<sup>c</sup> Database searches count publications by date and cannot confirm that the work was written during residency; they may include work begun before residency or published in the 12 months after graduation, so proportions may be higher.
<sup>d</sup> Two of the 4 mixed-source studies came from single programs and were measured before a research intervention began.
<sup>e</sup> Half (2) of the 4 program-records studies were pre-intervention cohorts from single programs.
<sup>f</sup> Excluding 6 studies that also counted abstracts or other items.
<sup>g</sup> Appropriate sample frame, complete coverage of the cohort, valid measurement, and a strict training window.
<sup>h</sup> Mayo Clinic internal medicine, which has a mandatory research curriculum.
<sup>i</sup> Studies that counted only first-author publications, added as a lower bound.
<sup>j</sup> In these 16 studies, the AI agents disagreed on eligibility or on extracted values, and I.-C.T. adjudicated against the source; without them, the pooled proportion was 53.0%, similar to the primary analysis, so the conclusions did not depend on these adjudications. Supplementary Material 1, Table S5, records study-level decisions.
<sup>k</sup> A larger cohort of the same specialty and graduation years may include some of the same residents, but there is no clear evidence of overlap; this analysis removes the smaller cohort (1 ophthalmology and 1 orthopaedic surgery study).

In an exploratory meta-regression that included specialty group, data source, and pre-intervention status, medical cohorts had less than one-third the odds of publication of surgical cohorts (logit difference, −1.28; OR, 0.28; 95% CI, 0.12–0.63; *p* = 0.002). In other words, when data source and pre-intervention status were held equal, medical cohorts remained clearly lower. With *t*-tests, the medical OR was 0.28 (95% CI, 0.12–0.65; *p* = 0.005); in the parsimonious model, it was 0.28 (95% CI, 0.13– 0.61; *p* = 0.001). The 3 covariates reduced τ² from 1.51 to 0.68, which still indicates substantial heterogeneity.

The specialty gap did not depend on pre-intervention cohorts. The 5 pre-intervention cohorts in the primary analysis were spread across 2 specialty groups: 3 surgical and 2 medical (1 internal medicine and 1 family medicine). Without all of them, surgical studies had 71.9% (15 studies) and medical studies 38.1% (6 studies, 1,777 residents; 95% CI, 20.2%–60.1%), an unadjusted OR of 0.24 (95% CI, 0.09–0.64; *p* = 0.004), close to the unadjusted OR of 0.20 obtained with them.

Results varied widely within medical specialties. The highest estimate came from Mayo Clinic internal medicine (430 of 522 residents, 82.4%; PubMed-indexed publications) [32], where research has been a mandatory part of training since 1988 [16]. The lowest were family medicine (0 of 14 residents), pediatrics (13.4%), and physical medicine and rehabilitation (19.1%).

### 3.4. Secondary Outcomes

#### 3.4.1. Per Resident-Year, Medical Specialties Still Published Less

Counting publications in any author position, residents published 0.42 articles per resident-year (72 US studies; 95% CI, 0.32–0.56; prediction interval, 0.04–4.43). Three studies were not entered into the model: 1 because only an approximate, non-integer publication total was available, and 2 because they counted only first-author publications. Of the 72 retained studies, 30 used approximate counts derived from summary statistics. Excluding all approximate counts gave 0.27 articles per resident-year (42 studies; 95% CI, 0.18–0.40). Pre-intervention cohorts made up 30 of the 72 studies. Excluding these cohorts gave 0.79 (42 studies; 95% CI, 0.63–1.01). Thirty studies used assumed resident-years; excluding them gave 0.31 (42 studies; 95% CI, 0.23–0.43). Excluding both pre-intervention cohorts and assumed resident-years gave 0.55 (20 studies; 95% CI, 0.39–0.77) (Figure 2).

**Figure 2.**
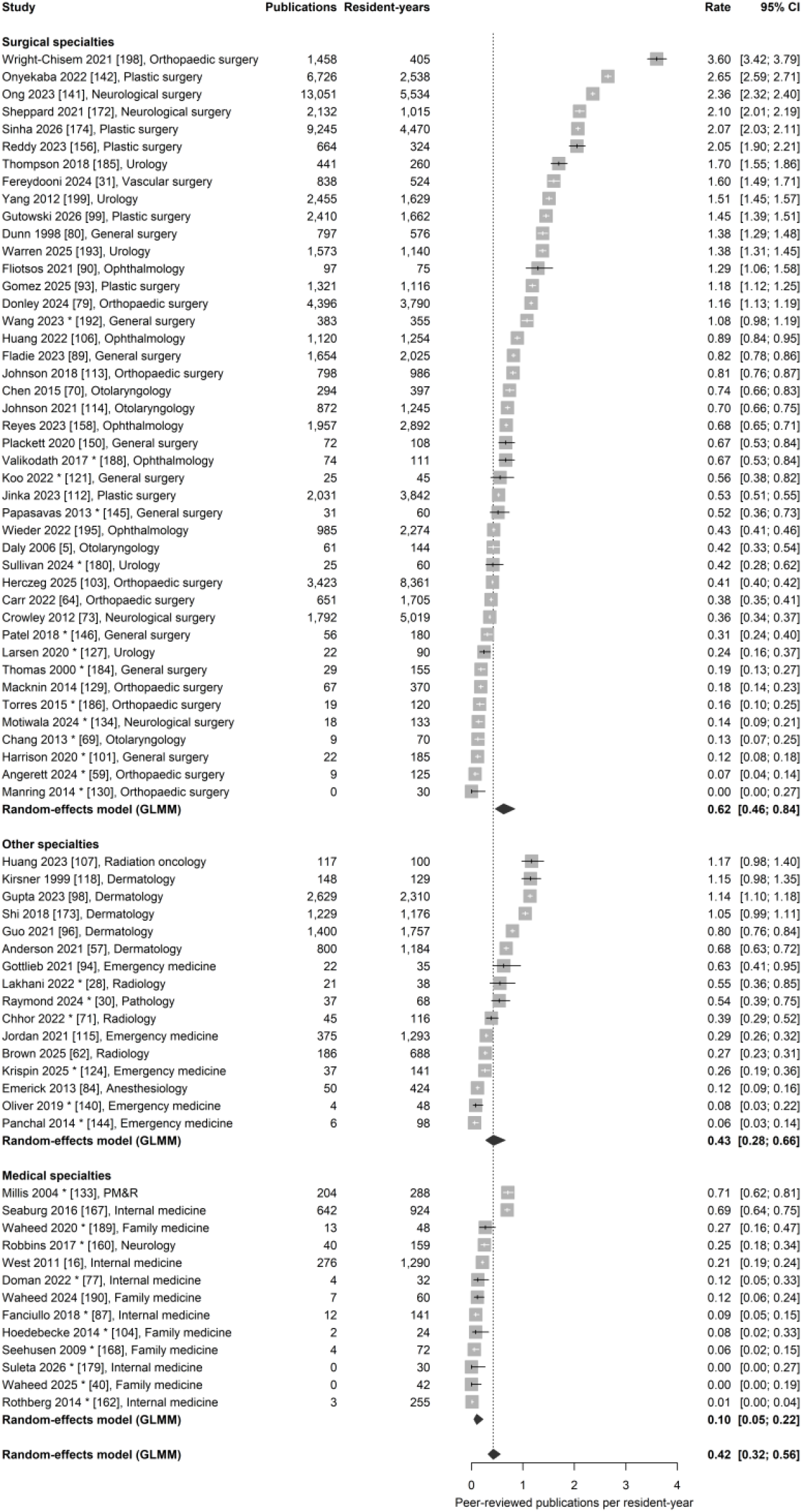
Peer-reviewed publications per resident-year in 72 US studies, by specialty group. Each label gives the first author, year, reference number in square brackets, and the specialty of the cohort. Squares show study rates and horizontal lines their 95% CIs. Rows labeled Random-effects model (GLMM) show pooled estimates from random-effects Poisson-normal generalized linear mixed models. Asterisks mark single-program pre-intervention cohorts. Without pre-intervention cohorts, surgical studies had 0.98 and medical studies 0.27 publications per resident-year (rate ratio, 0.28; 95% CI, 0.12–0.65). GLMM, generalized linear mixed model; PM&R, physical medicine and rehabilitation.

Per resident-year, medical specialties still published less than surgical specialties. With all studies, surgical studies had 0.62 articles per resident-year (43 studies) and medical studies 0.10 (13 studies). Without pre-intervention cohorts, surgical studies had 0.98 (29 studies; 95% CI, 0.76–1.26) and medical studies 0.27 (3 studies; 95% CI, 0.12–0.63), less than one-third of the surgical rate (rate ratio, 0.28; 95% CI, 0.12–0.65; *p* = 0.003).

#### 3.4.2. Residents With at Least 1 First-Author Publication

In 12 studies with 10,098 residents, 46.8% had at least 1 first-author publication during residency (95% CI, 34.5%–59.5%).

### 3.5. No Clear Rise Over 40 Years

Across graduation years from 1985 to 2028, the proportion showed no upward trend (slope, 0.002 logit per year; 95% CI, −0.044 to 0.047; *p* = 0.95; 29 studies with graduation years; Figure 3).

**Figure 3.**
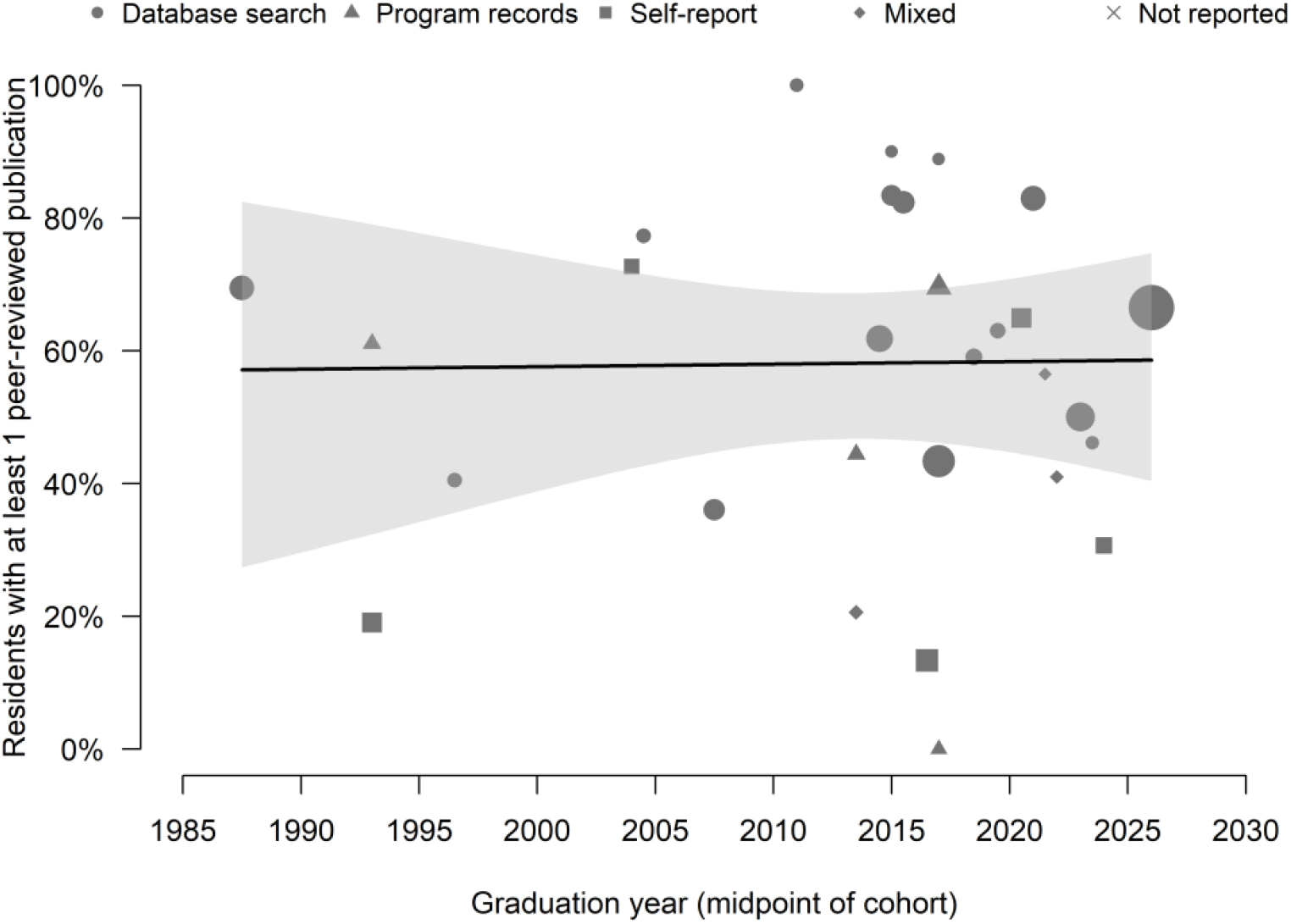
Proportion of residents with at least 1 peer-reviewed publication by the graduation-year midpoint of each cohort, 29 US studies. Marker size increases with the number of residents, and marker shape shows the data source. The line shows the fitted random-effects meta-regression, and the shaded band its 95% CI: slope 0.002 logit per year (95% CI, −0.044 to 0.047; *p* = 0.95).

### 3.6. Training Length and Data Source Mattered, but the Specialty Gap Remained

We examined three moderators: training length, data source, and training window. Each was associated with the proportion, but none explained the difference between specialties: proportions still differed widely among specialties of the same length, medical specialties remained lower after adjustment for data source, and medical specialties remained lower among studies with a strict training window.

In 26 US studies without pre-intervention cohorts and with an assigned length, proportions rose from 45.0% in 3-year specialties (6 studies) and 58.5% in 4-year specialties (7 studies) to 77.8% in 5-year specialties (12 studies); the single 7-year neurological surgery study had 69.5% (Supplementary Material 1, Figure S3 and Table S8). A linear model gave an OR of 1.68 per year (95% CI, 1.10–2.57).

Ophthalmology (60.8%; 4 studies) and physical medicine and rehabilitation (19.1%; 1 study), with the same 4 years, differed more than threefold. This contrast is itself between a surgical and a medical specialty. The other two 4-year studies were in dermatology (grouped with the other specialties), with 83.4%, and anesthesiology, with 59.1%. Length and specialty are highly collinear: 5 of the 6 three-year studies were medical, and all studies of 5 years or more were surgical or radiation oncology. When both entered the model, medical specialties remained lower (medical vs. all other specialties, OR, 0.29; 95% CI, 0.09–0.97; *p* = 0.045), and length was no longer significant (OR per year, 1.16; 95% CI, 0.69–1.97; *p* = 0.58).

Database-search studies had a pooled proportion of 71.5% (19 studies), compared with 39.5% for mixed sources (4 studies), 38.8% for program records (4 studies), and 37.3% for self-report (5 studies). Adjusted for specialty group, program records (OR, 0.34; 95% CI, 0.12–0.96) and self-report (OR, 0.32; 95% CI, 0.12–0.81) were associated with lower proportions than database search. Two of the 4 program-records studies were pre-intervention cohorts. After pre-intervention status was added, the omnibus test for data source was of borderline significance (*p* = 0.059). In the parsimonious model, database search vs. other sources had an OR of 2.38 (95% CI, 1.17–4.85; *p* = 0.017).

Estimates varied by window class (Supplementary Material 1, Table S4). The 22 studies with a strict training window had 63.7% (95% CI, 48.6%–76.5%), and the 7 with an extended window had 44.1%. In strict-window studies without pre-intervention cohorts, surgical cohorts had 78.7% (10 studies) and medical cohorts 46.1% (4 studies; OR, 0.23; 95% CI, 0.06–0.85). Studies of residents still in training had 51.5% (9 studies; 95% CI, 38.1%–64.6%); 5 of them were surgical. The 8 studies that covered only part of the training window had 14.4%, and 1 study had an undefined window. These classes differed in specialty and source, and we did not test them formally.

Protected research time was reported as present or absent in only 8 of 32 studies (6 yes, 2 no; a further study reported mixed exposure), too few for an estimate, so we report it descriptively. The 13 single-institution studies had a numerically lower pooled proportion than the 18 multi-institution studies (54.7% vs. 60.6%); 1 study did not report the number of institutions.

### 3.7. Sensitivity Analyses

Sensitivity analyses gave pooled proportions from 46.2% (all countries, 103 studies) to 73.8% (low risk of bias only, 12 studies) (Table 2). Without the Mayo cohort, the primary estimate was 58.6% (31 studies; 95% CI, 47.5%–68.9%). Six studies (5,110 residents, 38.0%) also counted abstracts or other items; the largest counted conference papers, book chapters, and editorials in Scopus [6]. Restricting to journal articles gave 57.6% (26 studies; 95% CI, 44.6%–69.6%); surgical and medical estimates were 66.8% and 27.7%. Adding the 4 first-author-only studies as a lower bound gave 60.1% (36 studies). Including the 7 extended-window studies gave 56.5% (39 studies), including the 8 partial-window studies gave 51.0% (40 studies), and including the survey with an unrestricted counting period gave 61.1% (33 studies).

In unadjusted comparisons, the direction of the specialty gap was the same in every study selection (Supplementary Material 1, Figure S4 and Table S9). Across 9 nested study selections, the OR for medical vs. surgical specialties ranged from 0.12 to 0.47; it was 0.20 in all US studies, including pre-intervention cohorts. The other 8 selections excluded these cohorts and restricted by data source, number of institutions, sampling frame, or window. The 95% CI excluded 1 in 6 of the 9 selections, including the strict-window selection (OR, 0.23; 95% CI, 0.06–0.85). The other 3 were database-search studies only (57.1% in 3 medical studies with 991 residents vs. 73.9% in 12 surgical studies with 10,199 residents; OR, 0.47; 95% CI, 0.12–1.88), database-search studies from more than 1 institution (1 medical study; OR, 0.23; 95% CI, 0.05–1.19), and the selection without self-report surveys (OR, 0.43; 95% CI, 0.14–1.31). The national sampling frame selection contained only 1 medical study. Leave-one-out ORs without pre-intervention cohorts ranged from 0.16 to 0.32, including removal of the Mayo cohort.

### 3.8. Distribution of Study Estimates

Supplementary Material 1, Figure S5 and Table S10, describe the distribution of US study estimates without pre-intervention cohorts; they give a rough reference, not a ranking of programs. Surgical estimates were narrower than medical estimates: median 71.9% (10th–90th percentile, 40.1%–90.7%; 15 studies). Medical specialties were very wide: median 38.1% (9.1%–79.1%; 6 studies). The interquartile range of the predictive distribution for medical specialties (20.4% to 59.7%) lay mostly below that for surgical specialties (56.2% to 83.6%). Other specialties had a median of 67.1% (31.3%– 90.1%; 6 studies), and all specialties combined 64.2% (27.6%–89.4%; 27 studies). Estimates for each study by specialty are shown in Supplementary Material 1, Figure S6.

## 4. Discussion

### 4.1. Main Findings and Implications

In a typical published US cohort, about 6 in 10 residents published a peer-reviewed article during training (59.6%; 95% CI, 48.7%–69.6%). Overall, most residents published during training, but specialties were not alike. Proportions were lowest in medical specialties: 29.6% in a typical medical cohort, compared with 68.7% in a typical surgical cohort. After adjustment for data source and pre-intervention status, the odds of publishing in medical cohorts were less than one-third of those in surgical cohorts (OR, 0.28; 95% CI, 0.12–0.63). Because surgical training is generally longer, we also counted publications per resident-year; medical specialties still published at less than one-third of the surgical rate (rate ratio, 0.28; 95% CI, 0.12–0.65). Within medical specialties, pediatrics (13.4%) and physical medicine and rehabilitation (19.1%) had the lowest cohorts, and in the only family medicine cohort (14 residents), no resident published during training.

The main implication of these findings is that they identify the specialties most in need of support. High proportions are attainable in medical specialties: the Mayo Clinic internal medicine cohort, with a mandatory research curriculum, reached 82.4% [32]. This suggests that medical-specialty programs that make research a required part of training may reach levels similar to those of surgical specialties. Resources such as protected research time, mentors, and research curricula may have the most room to improve output when they go first to the specialties with the lowest proportions. Residents in medical specialties often cite time, statistics, and research methods as their main barriers [33], and resident projects with statistical and methodological support were about twice as likely to be published [34]. Where a program’s own resources are limited, research-methods and writing support from outside the program may be one way to close the gap.

### 4.2. Significance of the Findings Compared With Existing Literature

Earlier systematic reviews asked how research curricula and interventions change resident output [9–12]; the proportion of residents who publish was outside their question. This review adds that proportion across specialties, with the resident as the unit, and so shows which specialties most need such interventions. The intervention literature also shows that the proportion can be raised: in a meta-analysis of 19 studies, the publication rate after a scholarly activity initiative was 2.6 times the rate before it (95% CI, 1.6–4.3) [11], although an earlier review found mixed effects of interventions on publications [10]. The research output that medical students bring to the residency Match has risen year by year [35], but the proportion who published during residency was largely flat across graduation years (Figure 3), which suggests that progress here has been limited. Residents arrive with more research experience, and residency is a time when that experience can be built on.

### 4.3. Possible Explanations for the Observed Results

Several mechanisms may contribute to the difference between specialties, and our data cannot separate them. One is selection: several specialties with high proportions, such as radiation oncology and orthopaedic surgery, select applicants with strong research records [35]. Another is work carried over from medical school: most studies counted every article published during residency, whenever the work began. The largest orthopaedic cohort counted publications from September 1 of the first training year [6], and an ophthalmology study noted that some articles completed before residency were counted [7]. The outcome therefore reflects publications that appear during residency or in the year after graduation, whenever the work began. Training structure may also contribute: most surgical specialties train for 5 years or more, and many residents take dedicated research years, built into accredited training in some specialties and taken outside it in others, such as general surgery [36,37]. Internal medicine, pediatrics, and family medicine train for 3 years, and protected time for research during residency is limited [38,39].

Training length alone, however, does not explain the gap. In the United States, most surgical specialties, such as orthopaedic surgery, urology, neurological surgery, and otolaryngology, are entered directly after medical school, so their residencies, like internal medicine residency, are complete programs that lead to board certification [19]. Among specialties of the same length, publication proportions still differed widely, and when length and specialty entered the same model, medical specialties remained lower while length was no longer significant (Supplementary Material 1, Table S8). The ACGME requires scholarly activity from residents in every specialty [1], whether or not they plan a fellowship.

Within medical specialties, the wide range more plausibly reflects program type and career plans. The highest internal medicine estimate came from an academic program with a mandatory research curriculum [32], and the lowest estimates came from clinically focused programs, such as a community hospital program [40]. In 3-year specialties, residents often apply for fellowship early in their final year, so publications meant for the application usually need to appear by the second year; in pediatrics, scholarly work is required of subspecialty fellows for board eligibility [41], so residents bound for fellowship may defer research to fellowship. Support in 3-year specialties therefore needs to start early: for research to appear before the fellowship application, it needs to begin in the first year.

### 4.4. Limitations

This review has three main limitations. First, studies counted publications in different ways, by database search, program records, or resident self-report, which may affect how individual estimates are read. We therefore adjusted for data source in the meta-regression and repeated the specialty comparison under 9 study selections restricted by data source, number of institutions, sampling frame, and training window, and with leave-one-out analysis. Medical specialties had lower proportions than surgical specialties in every selection, and the specialties with higher and lower proportions were largely the same.

Second, the estimates for each specialty come from different studies, and the studies are heterogeneous: training rules differ by specialty (some specialties begin counting at PGY-2, and surgical training often includes dedicated research years), and studies counted publications in different ways. Specialty, training length, and data source also tend to go together, so we cannot tell which of them produces the difference. However, each of the 32 US studies in the primary analysis covered a single specialty, and none compared specialties within one study, so a comparison across studies is the only way to answer this question with the available evidence. We therefore made the comparison as consistent as we could: counting windows and groups followed each specialty’s accredited residency, the unit by which programs and accreditors review resident output; we adjusted for data source and pre-intervention status; and we counted publications per resident-year to offset differences in training length. Under each of these approaches, medical specialties remained lower.

Third, we did not search Embase or Scopus. However, this review concerns US residents, and studies on this topic are mostly published in medical education and specialty journals indexed in PubMed, which is maintained by the US National Library of Medicine; at least 14 of the 19 database-search studies in the primary analysis themselves used PubMed or MEDLINE to count residents’ publications; meta-research also shows that searching databases beyond PubMed has a limited effect on the results of systematic reviews [42,43]. We also searched forward and backward citations of the studies included at each of two rounds and of 4 prior reviews through OpenAlex to find studies that the database searches might have missed.

## 5. Conclusions

About 60% of US residents publish during residency, but specialties differ widely, and medical specialties have the most room for improvement. We suggest that medical specialties explore other ways of teaching research, so that residents can complete a scholarly publication within training that is shorter than surgical training and learn the full process of producing knowledge.

### Supplementary Materials

Supporting information accompanies this manuscript. Supplementary Material 1: Figure S1: PRISMA 2020 flow diagram; Figure S2: Risk of bias summary (JBI checklist for prevalence studies); Figure S3: Proportion of residents with at least 1 publication by standard length of residency training; Figure S4: Medical vs. surgical odds ratio under study selections; Figure S5: Distribution of published US cohort estimates; Figure S6: Proportion of residents with at least 1 publication by specialty, US studies; Table S1: PRISMA 2020 checklist and PRISMA 2020 for Abstracts checklist; Table S2: Full search strategies for all sources; Table S3: Evidence table of all included studies; Table S4: Training-window classes, pooled proportions, and study composition; Table S5: Data extraction details for each included study; Table S6: Studies included with data provided by their corresponding authors; Table S7: Risk of bias ratings for each study; Table S8: Pooled proportions by standard training length, with a linear model; Table S9: Medical vs. surgical gap under study selections and leave-one-out analysis; Table S10: Distribution of published US cohort estimates by specialty group; Table S11: Reports excluded at full-text review, with reasons. Supplementary Material 2: Extraction dataset for the 172 included studies (.xlsx).

References 44–201 are cited in Table 1, Figures 1 and 2, and the Supplementary Materials.

### Author Contributions

Conceptualization, P.-L.C., W.-L.T., W.-C.L., and I.-C.T.; methodology, P.-L.C., W.-L.T., W.-C.L., and I.-C.T.; software, I.-C.T.; validation, W.-L.T., W.-C.L., and I.-C.T.; formal analysis, I.-C.T.; investigation, W.-C.L. and I.-C.T.; resources, P.-L.C., W.-L.T., W.-C.L., and I.-C.T.; data curation, P.-L.C., W.-L.T., and I.-C.T.; writing – original draft, I.-C.T.; writing – review and editing, P.-L.C., W.-L.T., W.-C.L., and I.-C.T.; visualization, P.-L.C., W.-L.T., W.-C.L., and I.-C.T.; supervision, I.-C.T.; project administration, P.-L.C., W.-L.T., W.-C.L., and I.-C.T. All authors have read and agreed to the published version of the manuscript.

### Funding

This research received no external funding.

### Institutional Review Board Statement

Not applicable. This study is a systematic review and meta-analysis of previously published data and did not involve human participants.

### Informed Consent Statement

Not applicable. This systematic review used only data from previously published studies.

### Data Availability Statement

The original contributions presented in this study are included in the article and the Supplementary Materials. The extraction methods are described in Section 2.7. The extracted data for every included study, with all variables used in the analyses and the location in the source article (page, table, or section) of each extracted count, are given in Supplementary Material 2 and, in part, in Supplementary Material 1, Tables S3 and S5. The search strategies are given in Supplementary Material 1, Table S2. Available screenshots and source-location records are available from the corresponding author on request. Further inquiries can be directed to the corresponding author.

## Acknowledgments

During the preparation of this study and manuscript, I.-C.T. used Claude Opus 5.5 and Claude Sonnet 5.5 (Anthropic), operated through Claude Code (Anthropic), and GPT-6 Astra (OpenAI), operated through Codex (OpenAI), between October 21 (protocol drafting) and October 29, 2026. Claude was used for protocol drafting, title and abstract screening, full-text eligibility assessment, risk-of-bias rating, data extraction (Claude Opus 5.5), proposing values for disagreements (Claude Sonnet 5.5), generating the R code for the statistical analyses, and English editing of the manuscript text. GPT-6 Astra was used for independent data extraction, blinded re-screening of sampled exclusions, and review of the manuscript. The authors checked random samples of 10–20% of these outputs for accuracy, supervised and reviewed these outputs, and are responsible for the content of the submitted publication.

## Supplementary Material 1

**Who Publishes During Residency? A Systematic Review and Meta-Analysis of Peer-Reviewed Publication Among Residents in the United States**

**Table S1.**
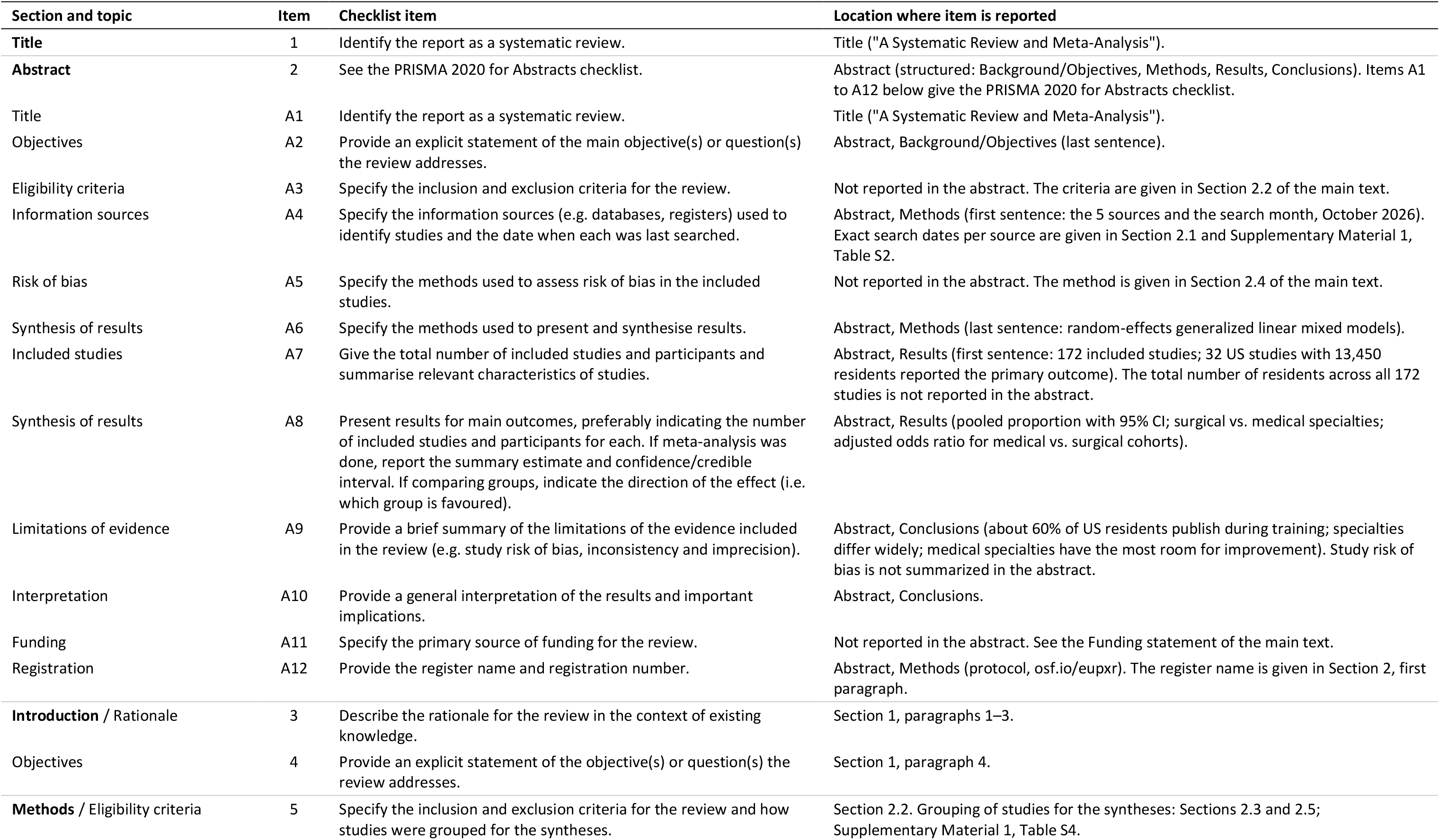
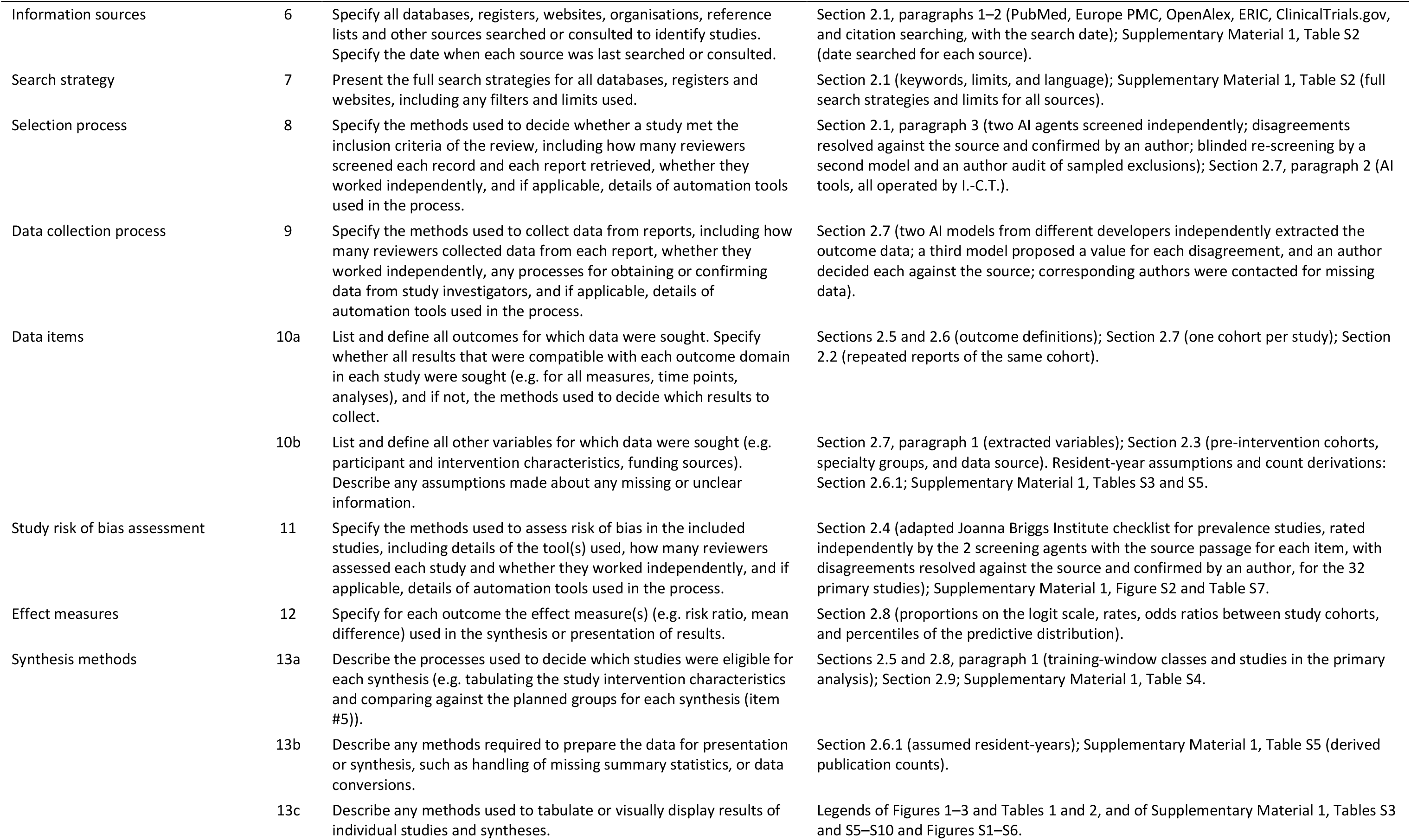
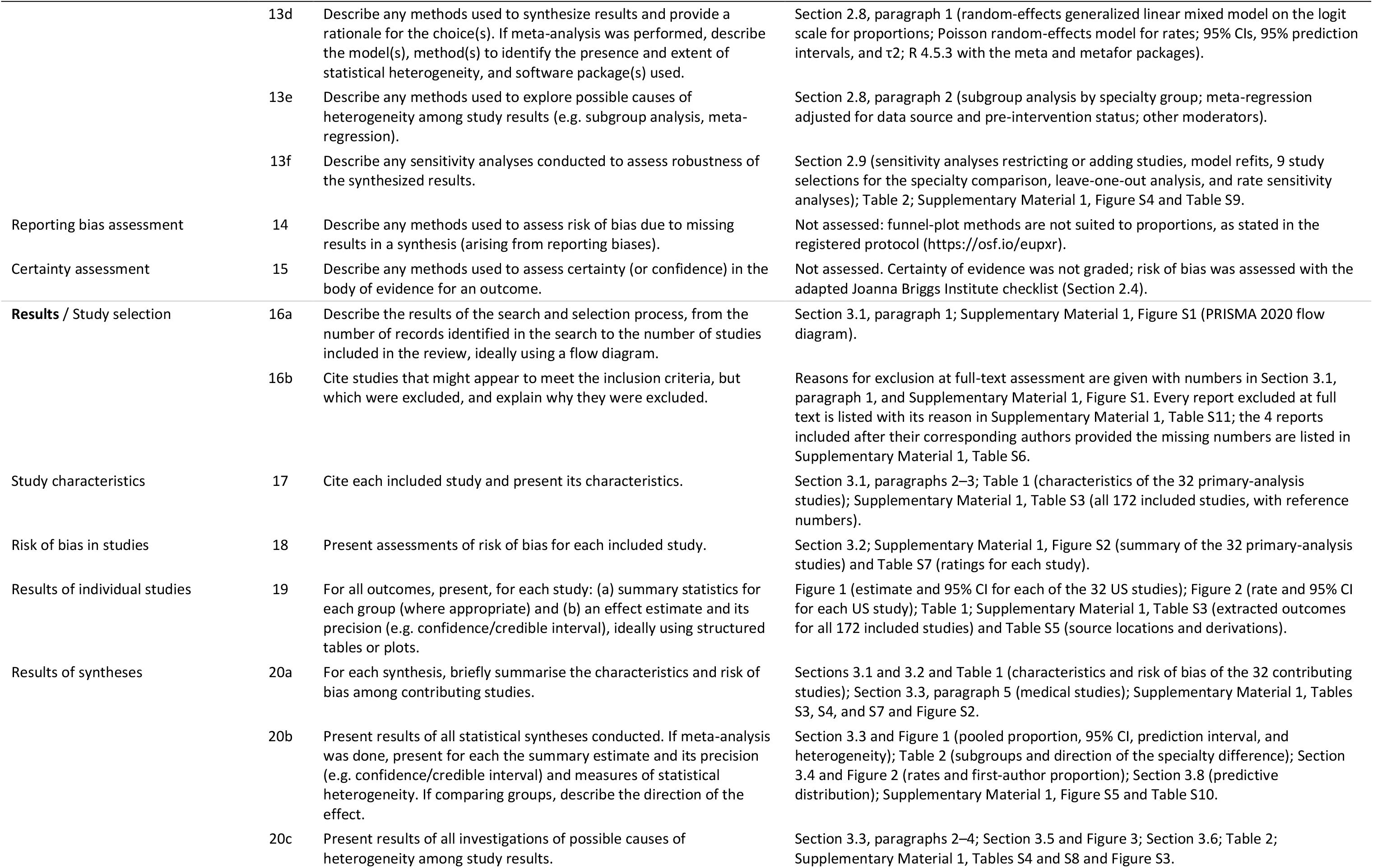
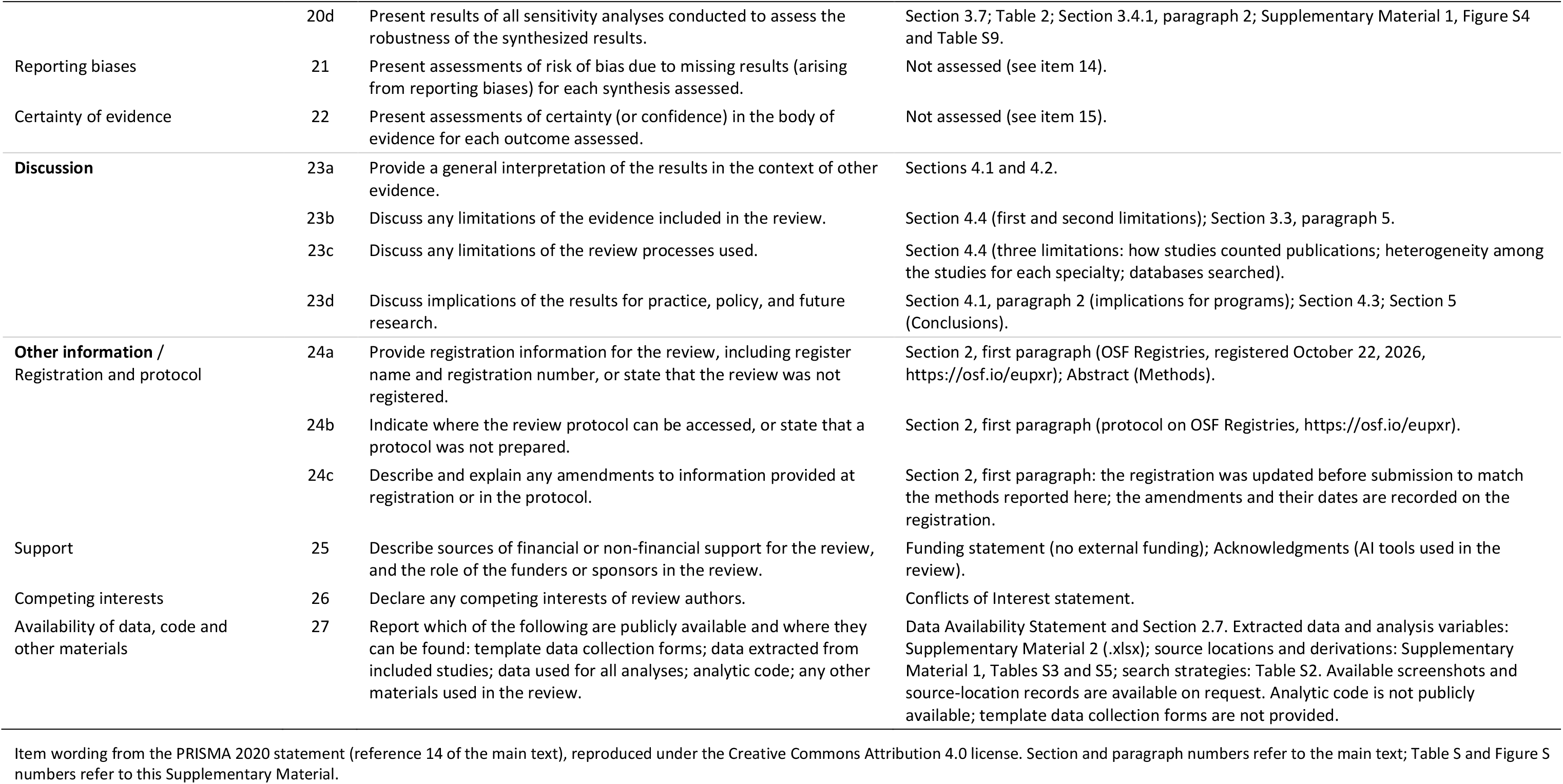
PRISMA 2020 checklist and PRISMA 2020 for Abstracts checklist. Location in this report of each item of the PRISMA 2020 checklist (27 items) and of the PRISMA 2020 for Abstracts checklist (12 items). PRISMA, Preferred Reporting Items for Systematic Reviews and Meta-Analyses.

**Table S2.** Full search strategies for all sources. Dates, exact search strings and limits, and numbers of records retrieved.

| Source | Date searched | Search string and limits | Records retrieved, n |
|---|---|---|---|
| **Main searches, publications from 2000 onward** |  |  |  |
| PubMed | 2026-10-22 | ("Internship and Residency"[Mesh] OR resident[tiab] OR residents[tiab] OR residency[tiab] OR "house staff"[tiab] OR housestaff[tiab] OR trainee*[tiab]) AND (publication*[tiab] OR publish*[tiab] OR "scholarly activit*"[tiab] OR "scholarly output"[tiab] OR "research productivity"[tiab] OR "research output"[tiab] OR "peer-reviewed"[tiab] OR bibliometric*[tiab] OR "h-index"[tiab]) AND (rate*[tiab] OR proportion*[tiab] OR percent*[tiab] OR prevalence[tiab] OR number[tiab] OR "per resident"[tiab]) NOT (Editorial[pt] OR Letter[pt] OR Comment[pt]) AND 2000:2026[dp] AND english[la] | 5,744 |
| Europe PMC | 2026-10-22 | (ABSTRACT:(resident OR residents OR residency OR "house staff" OR trainee*) OR TITLE:(resident OR residents OR residency OR "house staff" OR trainee*)) AND (ABSTRACT:(publication* OR publish* OR "scholarly activity" OR "scholarly output" OR "research productivity" OR "research output" OR "peer-reviewed" OR bibliometric* OR "h-index") OR TITLE:(publication* OR publish* OR "scholarly activity" OR "research productivity" OR bibliometric*)) AND ABSTRACT:(rate* OR proportion* OR percent* OR prevalence OR number OR "per resident") AND PUB_YEAR:[2000 TO 2026] AND LANG:eng<br>Records indexed as editorials, letters, or comments were removed after retrieval. | 5,663 |
| OpenAlex | 2026-10-22 | (resident OR residents OR residency) AND (publication OR publications OR "scholarly activity" OR "research productivity") AND (rate OR proportion OR percentage)<br>Filter: title_and_abstract.search; language English; type article; publication date from January 1, 2000. | 3,728 |
| ERIC | 2026-10-22 | (resident OR residents OR residency) AND (publication OR publications OR "scholarly activity" OR "research productivity" OR "research output") AND (medical OR graduate medical education)<br>No date limit in the query; records were split by publication year after retrieval. | 82 |
| ClinicalTrials.gov | 2026-10-22 | (residents OR residency OR resident physicians) AND (publication OR scholarly activity OR research productivity OR research curriculum)<br>Searched as the query.term parameter; no date limit. | 200 |
| PubMed, supplementary title search<sup>a</sup> | 2026-10-22 | (resident[ti] OR residents[ti] OR residency[ti] OR trainee*[ti] OR "house staff"[ti]) AND (publication*[ti] OR publish*[ti] OR scholar*[ti] OR "research productivity"[ti] OR "research output"[ti] OR "academic productivity"[ti] OR bibliometric*[ti]) AND 2000:2026[dp] AND english[la] NOT (Editorial[pt] OR Letter[pt] OR Comment[pt] OR News[pt]) | 610 |
| **Searches for publications before 2000<sup>b</sup>** |  |  |  |
| PubMed | 2026-10-22 | ("Internship and Residency"[Mesh] OR resident[tiab] OR residents[tiab] OR residency[tiab] OR "house staff"[tiab] OR housestaff[tiab] OR trainee*[tiab]) AND (publication*[tiab] OR publish*[tiab] OR "scholarly activit*"[tiab] OR "scholarly output"[tiab] OR "research productivity"[tiab] OR "research output"[tiab] OR "peer-reviewed"[tiab] OR bibliometric*[tiab] OR "h-index"[tiab]) AND (rate*[tiab] OR proportion*[tiab] OR percent*[tiab] OR prevalence[tiab] OR number[tiab] OR "per resident"[tiab]) NOT (Editorial[pt] OR Letter[pt] OR Comment[pt]) AND 1000:1999[dp] AND english[la] | 335 |
| PubMed, supplementary title search | 2026-10-22 | (resident[ti] OR residents[ti] OR residency[ti] OR trainee*[ti] OR "house staff"[ti]) AND (publication*[ti] OR publish*[ti] OR scholar*[ti] OR "research productivity"[ti] OR "research output"[ti] OR "academic productivity"[ti] OR bibliometric*[ti]) AND 1000:1999[dp] AND english[la] NOT (Editorial[pt] OR Letter[pt] OR Comment[pt] OR News[pt]) | 17 |
| Europe PMC | 2026-10-22 | (ABSTRACT:(resident OR residents OR residency OR "house staff" OR trainee*) OR TITLE:(resident OR residents OR residency OR "house staff" OR trainee*)) AND (ABSTRACT:(publication* OR publish* OR "scholarly activity" OR "scholarly output" OR "research productivity" OR "research output" OR "peer-reviewed" OR bibliometric* OR "h-index") OR TITLE:(publication* OR publish* OR "scholarly activity" OR "research productivity" OR bibliometric*)) AND ABSTRACT:(rate* OR proportion* OR percent* OR prevalence OR number OR "per resident") AND PUB_YEAR:[1000 TO 1999] AND LANG:eng | 312 |
| OpenAlex | 2026-10-22 | (resident OR residents OR residency) AND (publication OR publications OR "scholarly activity" OR "research productivity") AND (rate OR proportion OR percentage)<br>Filter: title_and_abstract.search; language English; type article; publication date to December 31, 1999. | 221 |
| ERIC | 2026-10-22 | Same query as the main search; records published before 2000. | 200 |
| ClinicalTrials.gov | 2026-10-22 | Not repeated; the main search had no date limit. | 0 |
| **Registry supplementary search** |  |  |  |
| ClinicalTrials.gov | 2026-10-27 | (residents OR residency OR resident physicians) AND (publication OR scholarly activity OR research productivity OR research curriculum)<br>Same query, all result pages retrieved; no date limit. Of the records retrieved, 200 had previously retrieved registry identifiers and 53 were new to the local collection. These cannot all be assumed to have been present at the original search date. | 253 |
| **Citation searching (other methods)<sup>c</sup>** |  |  |  |
| OpenAlex, included studies, round 1 | 2026-10-23 | Backward: references of each seed; forward: works citing each seed (OpenAlex cites filter). Seeds: 143 studies included at that date. English-language records only. Of 3,125 candidates, 728 were already retrieved, 87 were not in English, and 21 had no title. | 2,289 |
| OpenAlex, included studies, round 2 | 2026-10-24 | As round 1, with all 161 studies included at that date as seeds. Of 3,397 candidates, 2,996 were already retrieved, 95 were not in English, and 56 had no title. | 250 |
| OpenAlex, prior reviews, forward | 2026-10-23 | Works citing 4 prior systematic reviews of resident research (Hebert 2003; Stevenson 2017; Wood 2018; Zimmerman 2019), after removal of records already retrieved. | 56 |
| Prior reviews, backward | 2026-10-23 | Reference lists of the same 4 reviews, checked against records already retrieved. | 13 |

Records identified: 16,912 from databases and 453 from ClinicalTrials.gov; 7,533 duplicates and 27 other records were removed, leaving 9,805 records screened. The records removed for other reasons (n=27) represent a historical PubMed identification/parser discrepancy whose individual identities could not be completely reconciled. ERIC, Education Resources Information Center.
<sup>a</sup>Title-level search added because short reports without abstracts may lack the rate terms; new records are those not retrieved by the other searches.
<sup>b</sup>Search of publication years before 2000. The rows sum to 1,085 records. Of these, 4 title-search records already returned by the PubMed search were not stored again; of the remaining 1,081, 355 were duplicates and 726 were new.
<sup>c</sup>OpenAlex was the only citation source. Citation searching identified 2,608 records in total.

**Table S3.**
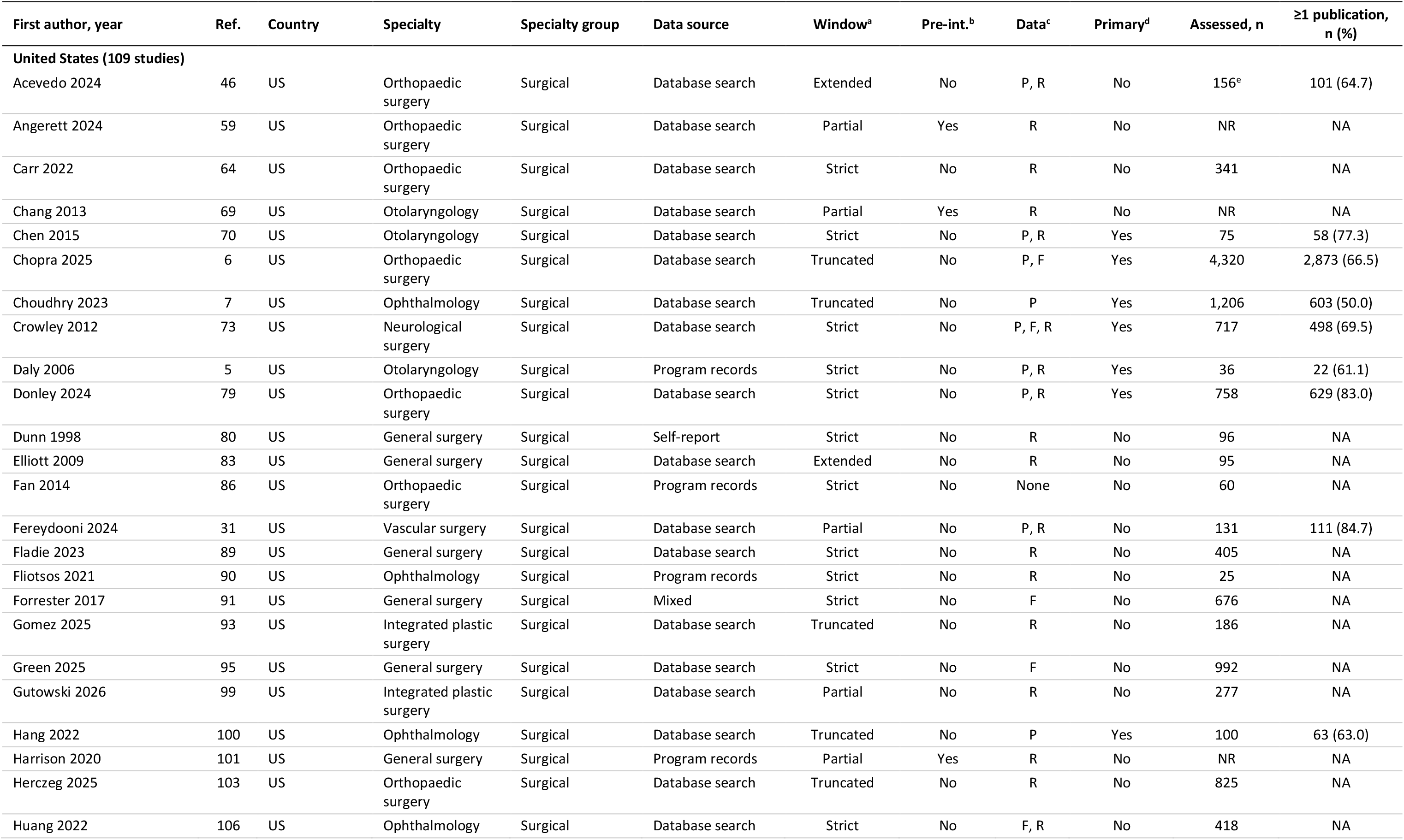
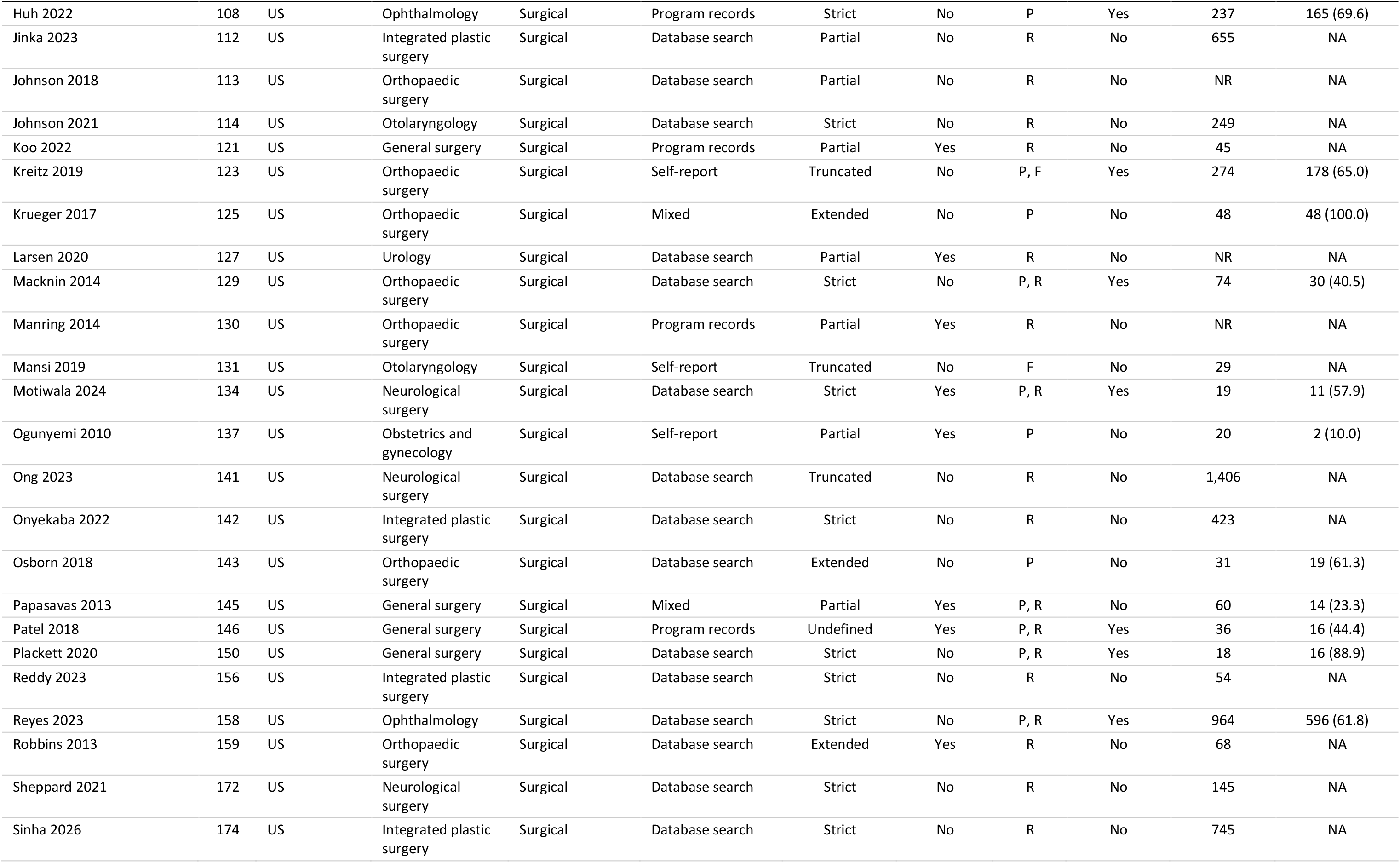
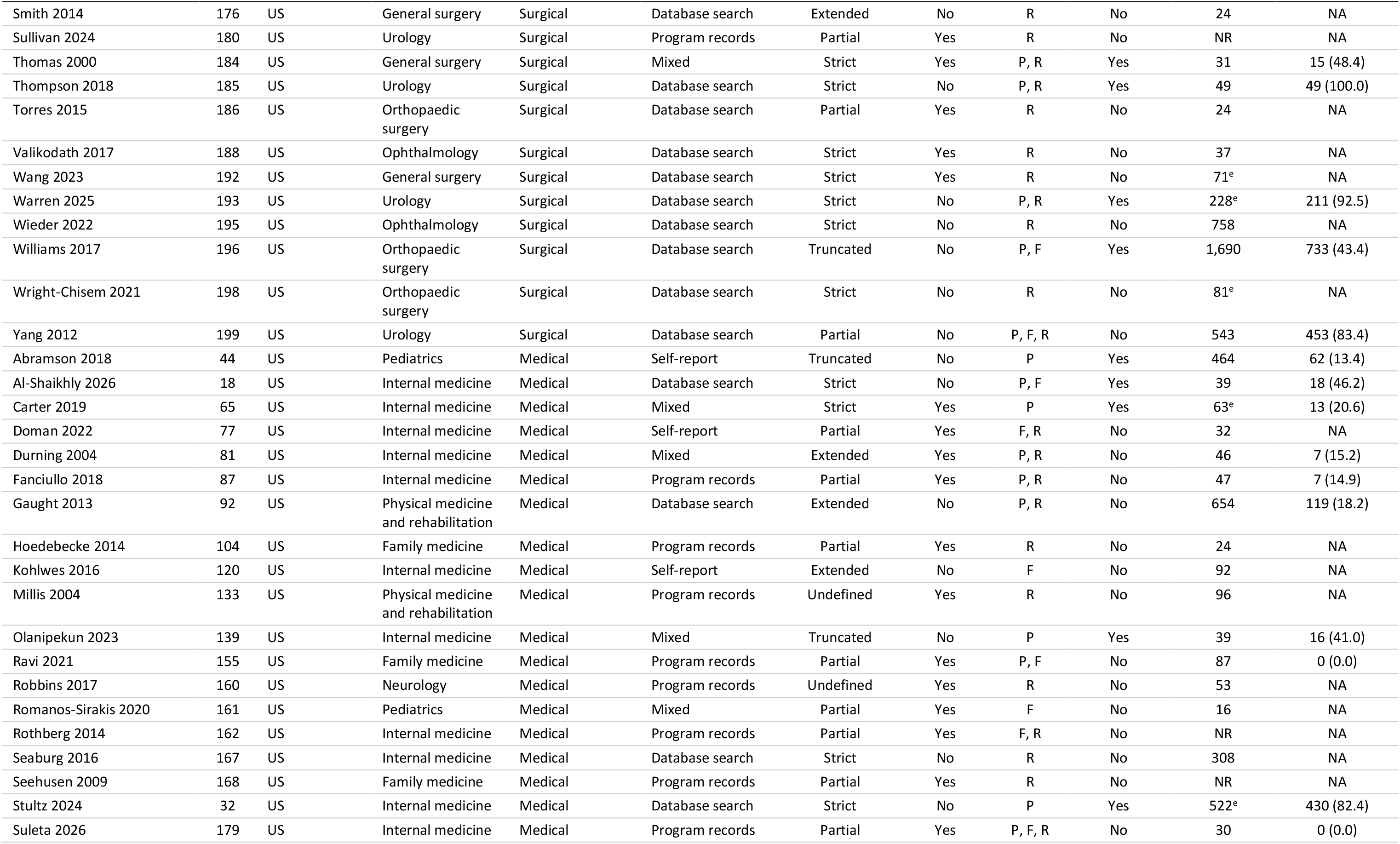
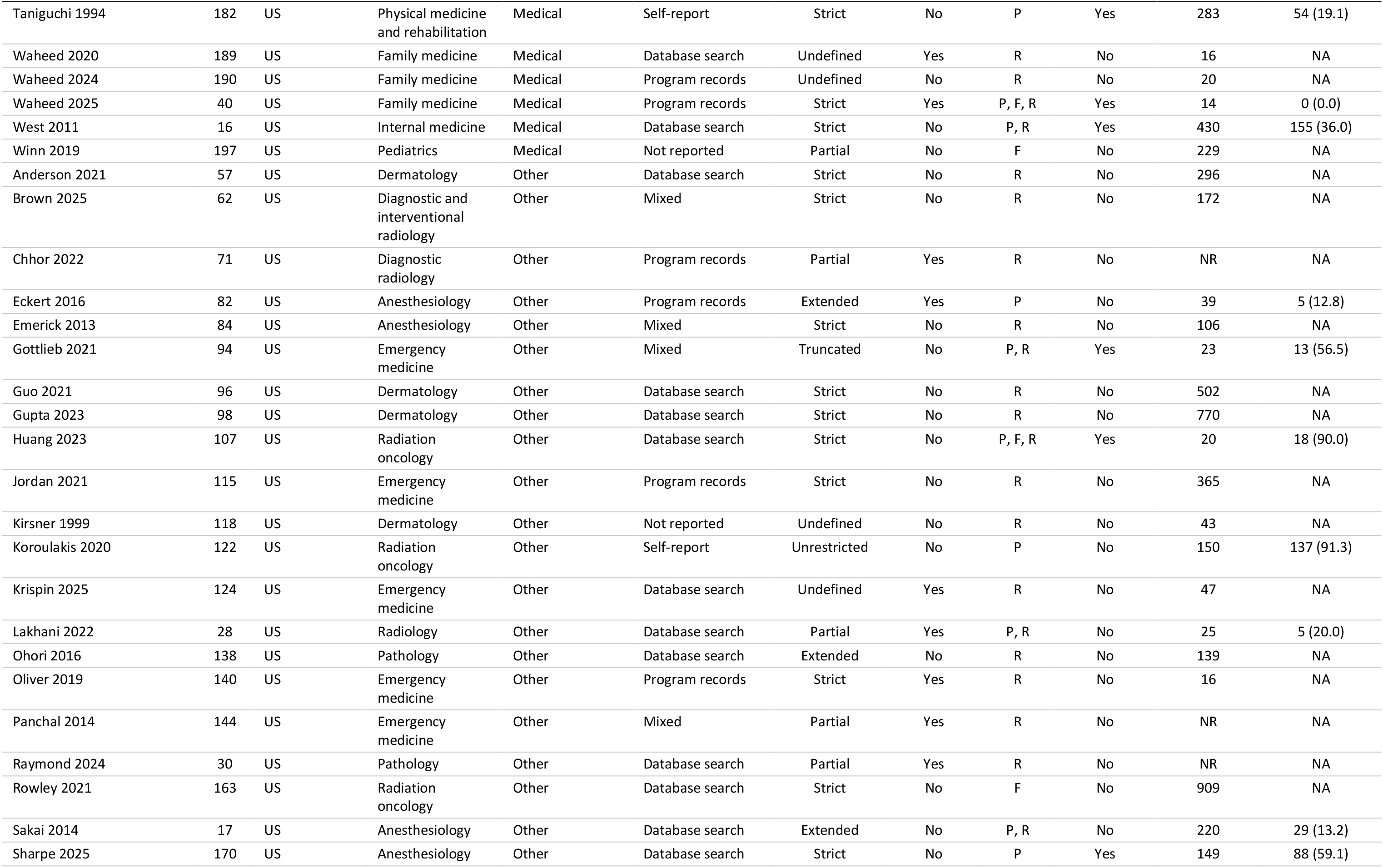
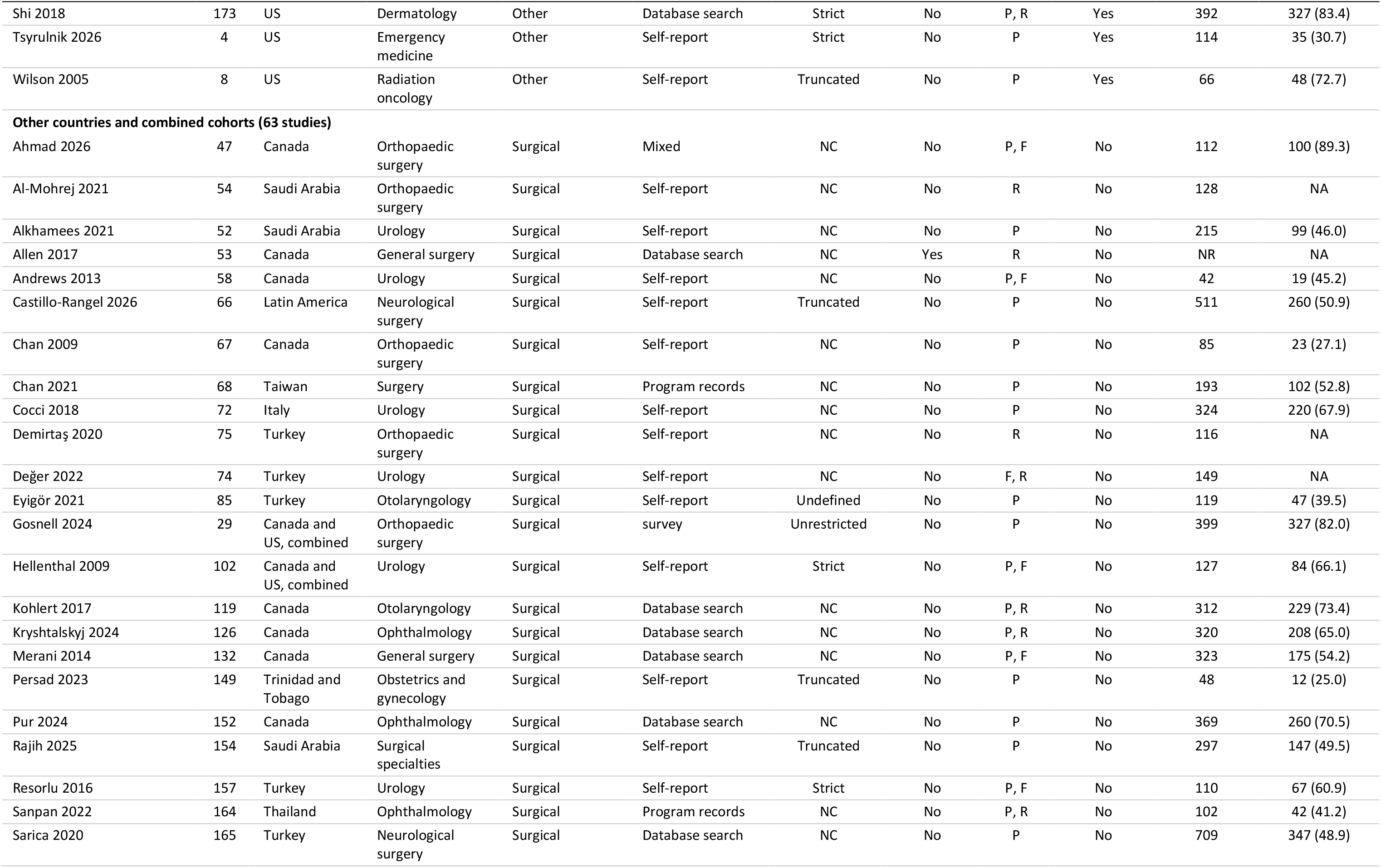
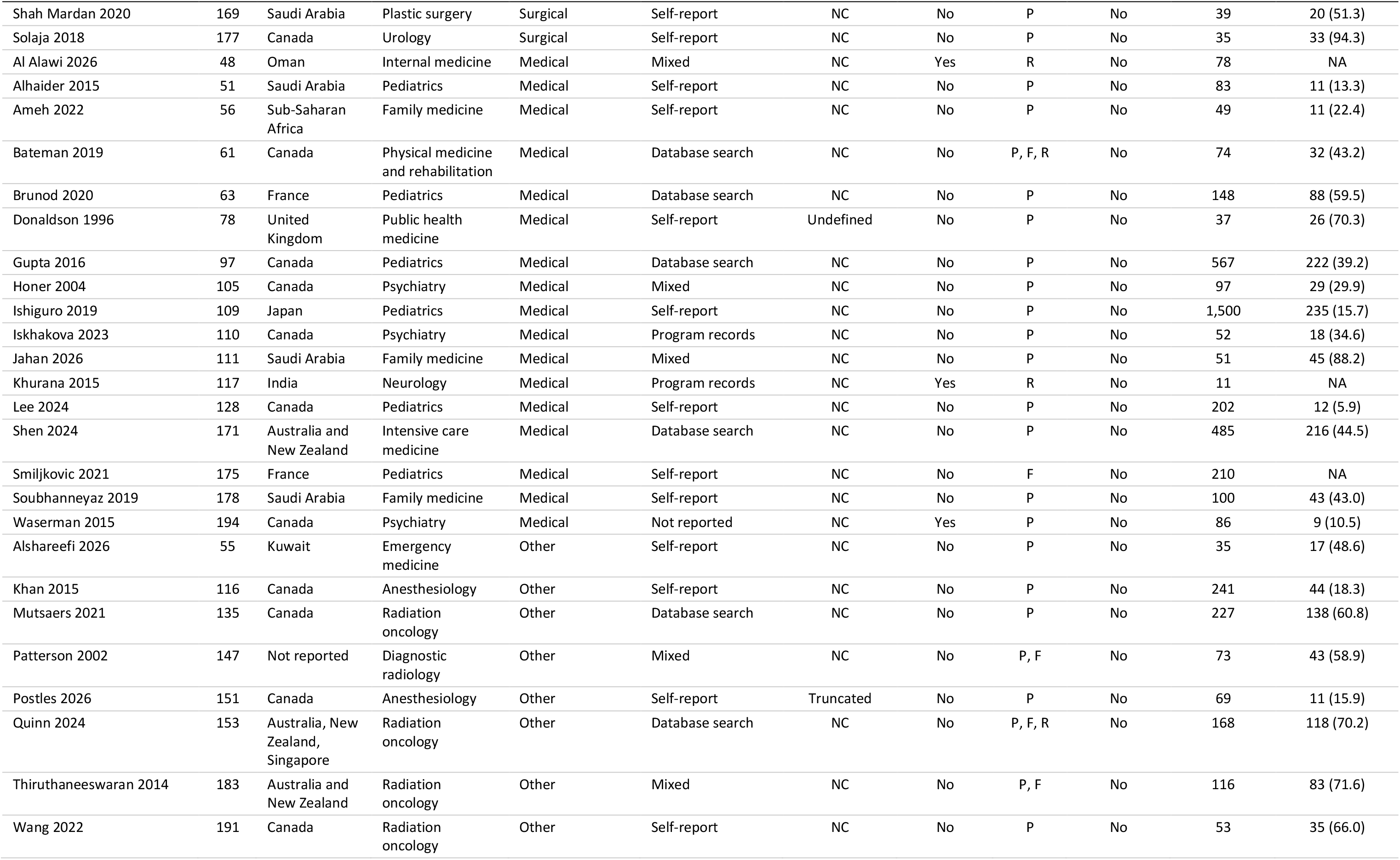
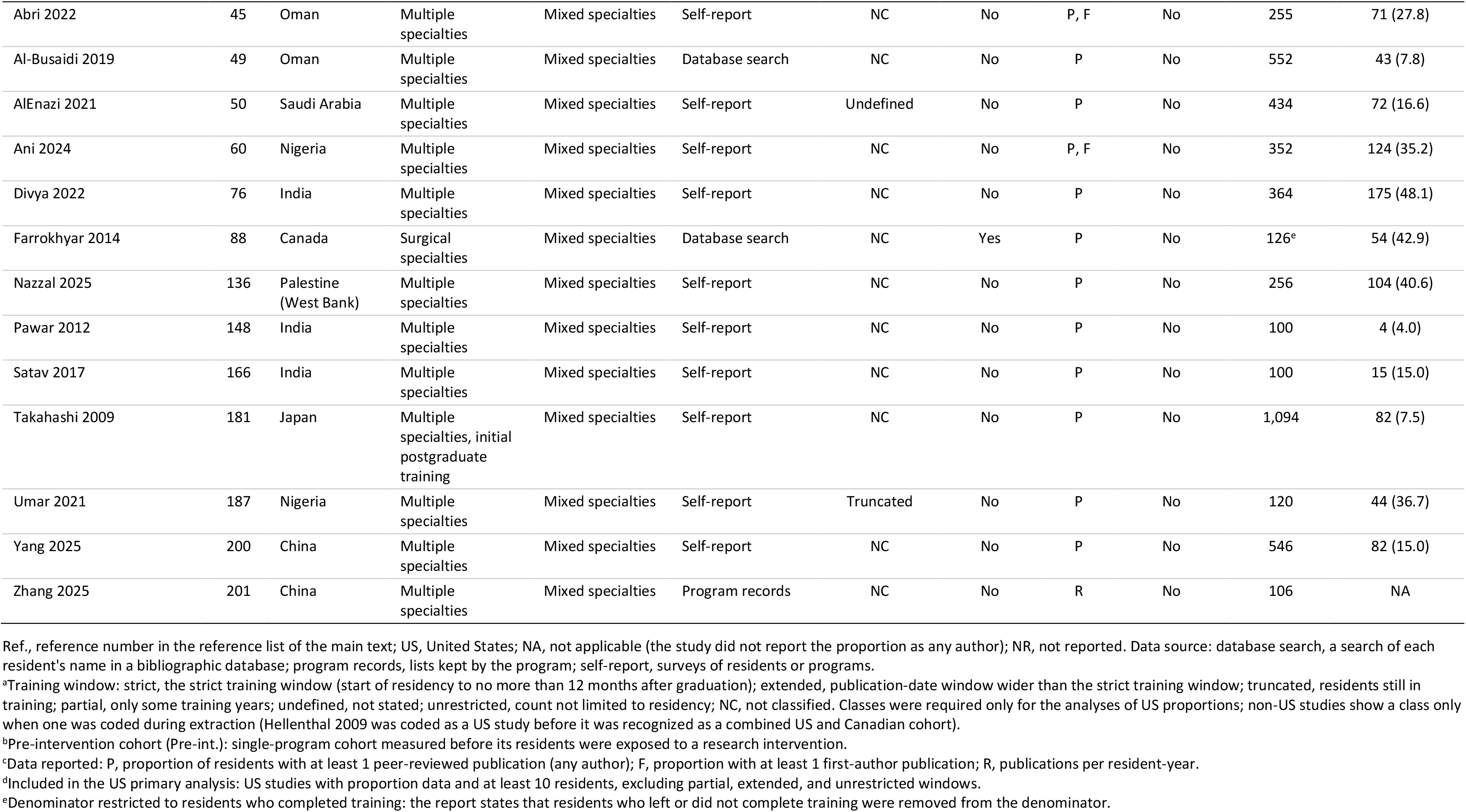
Evidence table of all included studies. All 172 included studies, by country and specialty group; the US primary analysis comprised 32 studies (13,450 residents).

| First author, year | Ref. | Country | Specialty | Specialty group | Data source | Window<sup>a</sup> | Pre-int.<sup>b</sup> | Data<sup>c</sup> | Primary<sup>d</sup> | Assessed, n | ≥1 publication, n (%) |
|---|---|---|---|---|---|---|---|---|---|---|---|
| **United States (109 studies)** |  |  |  |  |  |  |  |  |  |  |  |
| Acevedo 2024 | 46 | US | Orthopaedic surgery | Surgical | Database search | Extended | No | P, R | No | 156<sup>e</sup> | 101 (64.7) |
| Angerett 2024 | 59 | US | Orthopaedic surgery | Surgical | Database search | Partial | Yes | R | No | NR | NA |
| Carr 2022 | 64 | US | Orthopaedic surgery | Surgical | Database search | Strict | No | R | No | 341 | NA |
| Chang 2013 | 69 | US | Otolaryngology | Surgical | Database search | Partial | Yes | R | No | NR | NA |
| Chen 2015 | 70 | US | Otolaryngology | Surgical | Database search | Strict | No | P, R | Yes | 75 | 58 (77.3) |
| Chopra 2025 | 6 | US | Orthopaedic surgery | Surgical | Database search | Truncated | No | P, F | Yes | 4,320 | 2,873 (66.5) |
| Choudhry 2023 | 7 | US | Ophthalmology | Surgical | Database search | Truncated | No | P | Yes | 1,206 | 603 (50.0) |
| Crowley 2012 | 73 | US | Neurological surgery | Surgical | Database search | Strict | No | P, F, R | Yes | 717 | 498 (69.5) |
| Daly 2006 | 5 | US | Otolaryngology | Surgical | Program records | Strict | No | P, R | Yes | 36 | 22 (61.1) |
| Donley 2024 | 79 | US | Orthopaedic surgery | Surgical | Database search | Strict | No | P, R | Yes | 758 | 629 (83.0) |
| Dunn 1998 | 80 | US | General surgery | Surgical | Self-report | Strict | No | R | No | 96 | NA |
| Elliott 2009 | 83 | US | General surgery | Surgical | Database search | Extended | No | R | No | 95 | NA |
| Fan 2014 | 86 | US | Orthopaedic surgery | Surgical | Program records | Strict | No | None | No | 60 | NA |
| Fereydooni 2024 | 31 | US | Vascular surgery | Surgical | Database search | Partial | No | P, R | No | 131 | 111 (84.7) |
| Fladie 2023 | 89 | US | General surgery | Surgical | Database search | Strict | No | R | No | 405 | NA |
| Fliotsos 2021 | 90 | US | Ophthalmology | Surgical | Program records | Strict | No | R | No | 25 | NA |
| Forrester 2017 | 91 | US | General surgery | Surgical | Mixed | Strict | No | F | No | 676 | NA |
| Gomez 2025 | 93 | US | Integrated plastic surgery | Surgical | Database search | Truncated | No | R | No | 186 | NA |
| Green 2025 | 95 | US | General surgery | Surgical | Database search | Strict | No | F | No | 992 | NA |
| Gutowski 2026 | 99 | US | Integrated plastic surgery | Surgical | Database search | Partial | No | R | No | 277 | NA |
| Hang 2022 | 100 | US | Ophthalmology | Surgical | Database search | Truncated | No | P | Yes | 100 | 63 (63.0) |
| Harrison 2020 | 101 | US | General surgery | Surgical | Program records | Partial | Yes | R | No | NR | NA |
| Herczeg 2025 | 103 | US | Orthopaedic surgery | Surgical | Database search | Truncated | No | R | No | 825 | NA |
| Huang 2022 | 106 | US | Ophthalmology | Surgical | Database search | Strict | No | F, R | No | 418 | NA |
| Huh 2022 | 108 | US | Ophthalmology | Surgical | Program records | Strict | No | P | Yes | 237 | 165 (69.6) |
| Jinka 2023 | 112 | US | Integrated plastic surgery | Surgical | Database search | Partial | No | R | No | 655 | NA |
| Johnson 2018 | 113 | US | Orthopaedic surgery | Surgical | Database search | Partial | No | R | No | NR | NA |
| Johnson 2021 | 114 | US | Otolaryngology | Surgical | Database search | Strict | No | R | No | 249 | NA |
| Koo 2022 | 121 | US | General surgery | Surgical | Program records | Partial | Yes | R | No | 45 | NA |
| Kreitz 2019 | 123 | US | Orthopaedic surgery | Surgical | Self-report | Truncated | No | P, F | Yes | 274 | 178 (65.0) |
| Krueger 2017 | 125 | US | Orthopaedic surgery | Surgical | Mixed | Extended | No | P | No | 48 | 48 (100.0) |
| Larsen 2020 | 127 | US | Urology | Surgical | Database search | Partial | Yes | R | No | NR | NA |
| Macknin 2014 | 129 | US | Orthopaedic surgery | Surgical | Database search | Strict | No | P, R | Yes | 74 | 30 (40.5) |
| Manring 2014 | 130 | US | Orthopaedic surgery | Surgical | Program records | Partial | Yes | R | No | NR | NA |
| Mansi 2019 | 131 | US | Otolaryngology | Surgical | Self-report | Truncated | No | F | No | 29 | NA |
| Motiwala 2024 | 134 | US | Neurological surgery | Surgical | Database search | Strict | Yes | P, R | Yes | 19 | 11 (57.9) |
| Ogunyemi 2010 | 137 | US | Obstetrics and gynecology | Surgical | Self-report | Partial | Yes | P | No | 20 | 2 (10.0) |
| Ong 2023 | 141 | US | Neurological surgery | Surgical | Database search | Truncated | No | R | No | 1,406 | NA |
| Onyekaba 2022 | 142 | US | Integrated plastic surgery | Surgical | Database search | Strict | No | R | No | 423 | NA |
| Osborn 2018 | 143 | US | Orthopaedic surgery | Surgical | Database search | Extended | No | P | No | 31 | 19 (61.3) |
| Papasavas 2013 | 145 | US | General surgery | Surgical | Mixed | Partial | Yes | P, R | No | 60 | 14 (23.3) |
| Patel 2018 | 146 | US | General surgery | Surgical | Program records | Undefined | Yes | P, R | Yes | 36 | 16 (44.4) |
| Plackett 2020 | 150 | US | General surgery | Surgical | Database search | Strict | No | P, R | Yes | 18 | 16 (88.9) |
| Reddy 2023 | 156 | US | Integrated plastic surgery | Surgical | Database search | Strict | No | R | No | 54 | NA |
| Reyes 2023 | 158 | US | Ophthalmology | Surgical | Database search | Strict | No | P, R | Yes | 964 | 596 (61.8) |
| Robbins 2013 | 159 | US | Orthopaedic surgery | Surgical | Database search | Extended | Yes | R | No | 68 | NA |
| Sheppard 2021 | 172 | US | Neurological surgery | Surgical | Database search | Strict | No | R | No | 145 | NA |
| Sinha 2026 | 174 | US | Integrated plastic surgery | Surgical | Database search | Strict | No | R | No | 745 | NA |
| Smith 2014 | 176 | US | General surgery | Surgical | Database search | Extended | No | R | No | 24 | NA |
| Sullivan 2024 | 180 | US | Urology | Surgical | Program records | Partial | Yes | R | No | NR | NA |
| Thomas 2000 | 184 | US | General surgery | Surgical | Mixed | Strict | Yes | P, R | Yes | 31 | 15 (48.4) |
| Thompson 2018 | 185 | US | Urology | Surgical | Database search | Strict | No | P, R | Yes | 49 | 49 (100.0) |
| Torres 2015 | 186 | US | Orthopaedic surgery | Surgical | Database search | Partial | Yes | R | No | 24 | NA |
| Valikodath 2017 | 188 | US | Ophthalmology | Surgical | Database search | Strict | Yes | R | No | 37 | NA |
| Wang 2023 | 192 | US | General surgery | Surgical | Database search | Strict | Yes | R | No | 71<sup>e</sup> | NA |
| Warren 2025 | 193 | US | Urology | Surgical | Database search | Strict | No | P, R | Yes | 228<sup>e</sup> | 211 (92.5) |
| Wieder 2022 | 195 | US | Ophthalmology | Surgical | Database search | Strict | No | R | No | 758 | NA |
| Williams 2017 | 196 | US | Orthopaedic surgery | Surgical | Database search | Truncated | No | P, F | Yes | 1,690 | 733 (43.4) |
| Wright-Chisem 2021 | 198 | US | Orthopaedic surgery | Surgical | Database search | Strict | No | R | No | 81<sup>e</sup> | NA |
| Yang 2012 | 199 | US | Urology | Surgical | Database search | Partial | No | P, F, R | No | 543 | 453 (83.4) |
| Abramson 2018 | 44 | US | Pediatrics | Medical | Self-report | Truncated | No | P | Yes | 464 | 62 (13.4) |
| Al-Shaikhly 2026 | 18 | US | Internal medicine | Medical | Database search | Strict | No | P, F | Yes | 39 | 18 (46.2) |
| Carter 2019 | 65 | US | Internal medicine | Medical | Mixed | Strict | Yes | P | Yes | 63<sup>e</sup> | 13 (20.6) |
| Doman 2022 | 77 | US | Internal medicine | Medical | Self-report | Partial | Yes | F, R | No | 32 | NA |
| Durning 2004 | 81 | US | Internal medicine | Medical | Mixed | Extended | Yes | P, R | No | 46 | 7 (15.2) |
| Fanciullo 2018 | 87 | US | Internal medicine | Medical | Program records | Partial | Yes | P, R | No | 47 | 7 (14.9) |
| Gaught 2013 | 92 | US | Physical medicine and rehabilitation | Medical | Database search | Extended | No | P, R | No | 654 | 119 (18.2) |
| Hoedebecke 2014 | 104 | US | Family medicine | Medical | Program records | Partial | Yes | R | No | 24 | NA |
| Kohlwes 2016 | 120 | US | Internal medicine | Medical | Self-report | Extended | No | F | No | 92 | NA |
| Millis 2004 | 133 | US | Physical medicine and rehabilitation | Medical | Program records | Undefined | Yes | R | No | 96 | NA |
| Olanipekun 2023 | 139 | US | Internal medicine | Medical | Mixed | Truncated | No | P | Yes | 39 | 16 (41.0) |
| Ravi 2021 | 155 | US | Family medicine | Medical | Program records | Partial | Yes | P, F | No | 87 | 0 (0.0) |
| Robbins 2017 | 160 | US | Neurology | Medical | Program records | Undefined | Yes | R | No | 53 | NA |
| Romanos-Sirakis 2020 | 161 | US | Pediatrics | Medical | Mixed | Partial | Yes | F | No | 16 | NA |
| Rothberg 2014 | 162 | US | Internal medicine | Medical | Program records | Partial | Yes | F, R | No | NR | NA |
| Seaburg 2016 | 167 | US | Internal medicine | Medical | Database search | Strict | No | R | No | 308 | NA |
| Seehusen 2009 | 168 | US | Family medicine | Medical | Program records | Partial | Yes | R | No | NR | NA |
| Stultz 2024 | 32 | US | Internal medicine | Medical | Database search | Strict | No | P | Yes | 522<sup>e</sup> | 430 (82.4) |
| Suleta 2026 | 179 | US | Internal medicine | Medical | Program records | Partial | Yes | P, F, R | No | 30 | 0 (0.0) |
| Taniguchi 1994 | 182 | US | Physical medicine and rehabilitation | Medical | Self-report | Strict | No | P | Yes | 283 | 54 (19.1) |
| Waheed 2020 | 189 | US | Family medicine | Medical | Database search | Undefined | Yes | R | No | 16 | NA |
| Waheed 2024 | 190 | US | Family medicine | Medical | Program records | Undefined | No | R | No | 20 | NA |
| Waheed 2025 | 40 | US | Family medicine | Medical | Program records | Strict | Yes | P, F, R | Yes | 14 | 0 (0.0) |
| West 2011 | 16 | US | Internal medicine | Medical | Database search | Strict | No | P, R | Yes | 430 | 155 (36.0) |
| Winn 2019 | 197 | US | Pediatrics | Medical | Not reported | Partial | No | F | No | 229 | NA |
| Anderson 2021 | 57 | US | Dermatology | Other | Database search | Strict | No | R | No | 296 | NA |
| Brown 2025 | 62 | US | Diagnostic and interventional radiology | Other | Mixed | Strict | No | R | No | 172 | NA |
| Chhor 2022 | 71 | US | Diagnostic radiology | Other | Program records | Partial | Yes | R | No | NR | NA |
| Eckert 2016 | 82 | US | Anesthesiology | Other | Program records | Extended | Yes | P | No | 39 | 5 (12.8) |
| Emerick 2013 | 84 | US | Anesthesiology | Other | Mixed | Strict | No | R | No | 106 | NA |
| Gottlieb 2021 | 94 | US | Emergency medicine | Other | Mixed | Truncated | No | P, R | Yes | 23 | 13 (56.5) |
| Guo 2021 | 96 | US | Dermatology | Other | Database search | Strict | No | R | No | 502 | NA |
| Gupta 2023 | 98 | US | Dermatology | Other | Database search | Strict | No | R | No | 770 | NA |
| Huang 2023 | 107 | US | Radiation oncology | Other | Database search | Strict | No | P, F, R | Yes | 20 | 18 (90.0) |
| Jordan 2021 | 115 | US | Emergency medicine | Other | Program records | Strict | No | R | No | 365 | NA |
| Kirsner 1999 | 118 | US | Dermatology | Other | Not reported | Undefined | No | R | No | 43 | NA |
| Koroulakis 2020 | 122 | US | Radiation oncology | Other | Self-report | Unrestricted | No | P | No | 150 | 137 (91.3) |
| Krispin 2025 | 124 | US | Emergency medicine | Other | Database search | Undefined | Yes | R | No | 47 | NA |
| Lakhani 2022 | 28 | US | Radiology | Other | Database search | Partial | Yes | P, R | No | 25 | 5 (20.0) |
| Ohori 2016 | 138 | US | Pathology | Other | Database search | Extended | No | R | No | 139 | NA |
| Oliver 2019 | 140 | US | Emergency medicine | Other | Program records | Strict | Yes | R | No | 16 | NA |
| Panchal 2014 | 144 | US | Emergency medicine | Other | Mixed | Partial | Yes | R | No | NR | NA |
| Raymond 2024 | 30 | US | Pathology | Other | Database search | Partial | Yes | R | No | NR | NA |
| Rowley 2021 | 163 | US | Radiation oncology | Other | Database search | Strict | No | F | No | 909 | NA |
| Sakai 2014 | 17 | US | Anesthesiology | Other | Database search | Extended | No | P, R | No | 220 | 29 (13.2) |
| Sharpe 2025 | 170 | US | Anesthesiology | Other | Database search | Strict | No | P | Yes | 149 | 88 (59.1) |
| Shi 2018 | 173 | US | Dermatology | Other | Database search | Strict | No | P, R | Yes | 392 | 327 (83.4) |
| Tsyrulnik 2026 | 4 | US | Emergency medicine | Other | Self-report | Strict | No | P | Yes | 114 | 35 (30.7) |
| Wilson 2005 | 8 | US | Radiation oncology | Other | Self-report | Truncated | No | P | Yes | 66 | 48 (72.7) |
| **Other countries and combined cohorts (63 studies)** |  |  |  |  |  |  |  |  |  |  |  |
| Ahmad 2026 | 47 | Canada | Orthopaedic surgery | Surgical | Mixed | NC | No | P, F | No | 112 | 100 (89.3) |
| Al-Mohrej 2021 | 54 | Saudi Arabia | Orthopaedic surgery | Surgical | Self-report | NC | No | R | No | 128 | NA |
| Alkhamees 2021 | 52 | Saudi Arabia | Urology | Surgical | Self-report | NC | No | P | No | 215 | 99 (46.0) |
| Allen 2017 | 53 | Canada | General surgery | Surgical | Database search | NC | Yes | R | No | NR | NA |
| Andrews 2013 | 58 | Canada | Urology | Surgical | Self-report | NC | No | P, F | No | 42 | 19 (45.2) |
| Castillo-Rangel 2026 | 66 | Latin America | Neurological surgery | Surgical | Self-report | Truncated | No | P | No | 511 | 260 (50.9) |
| Chan 2009 | 67 | Canada | Orthopaedic surgery | Surgical | Self-report | NC | No | P | No | 85 | 23 (27.1) |
| Chan 2021 | 68 | Taiwan | Surgery | Surgical | Program records | NC | No | P | No | 193 | 102 (52.8) |
| Cocci 2018 | 72 | Italy | Urology | Surgical | Self-report | NC | No | P | No | 324 | 220 (67.9) |
| Demirtaş 2020 | 75 | Turkey | Orthopaedic surgery | Surgical | Self-report | NC | No | R | No | 116 | NA |
| Değer 2022 | 74 | Turkey | Urology | Surgical | Self-report | NC | No | F, R | No | 149 | NA |
| Eyigör 2021 | 85 | Turkey | Otolaryngology | Surgical | Self-report | Undefined | No | P | No | 119 | 47 (39.5) |
| Gosnell 2024 | 29 | Canada and US, combined | Orthopaedic surgery | Surgical | survey | Unrestricted | No | P | No | 399 | 327 (82.0) |
| Hellenthal 2009 | 102 | Canada and US, combined | Urology | Surgical | Self-report | Strict | No | P, F | No | 127 | 84 (66.1) |
| Kohlert 2017 | 119 | Canada | Otolaryngology | Surgical | Database search | NC | No | P, R | No | 312 | 229 (73.4) |
| Kryshtalskyj 2024 | 126 | Canada | Ophthalmology | Surgical | Database search | NC | No | P, R | No | 320 | 208 (65.0) |
| Merani 2014 | 132 | Canada | General surgery | Surgical | Database search | NC | No | P, F | No | 323 | 175 (54.2) |
| Persad 2023 | 149 | Trinidad and Tobago | Obstetrics and gynecology | Surgical | Self-report | Truncated | No | P | No | 48 | 12 (25.0) |
| Pur 2024 | 152 | Canada | Ophthalmology | Surgical | Database search | NC | No | P | No | 369 | 260 (70.5) |
| Rajih 2025 | 154 | Saudi Arabia | Surgical specialties | Surgical | Self-report | Truncated | No | P | No | 297 | 147 (49.5) |
| Resorlu 2016 | 157 | Turkey | Urology | Surgical | Self-report | Strict | No | P, F | No | 110 | 67 (60.9) |
| Sanpan 2022 | 164 | Thailand | Ophthalmology | Surgical | Program records | NC | No | P, R | No | 102 | 42 (41.2) |
| Sarica 2020 | 165 | Turkey | Neurological surgery | Surgical | Database search | NC | No | P | No | 709 | 347 (48.9) |
| Shah Mardan 2020 | 169 | Saudi Arabia | Plastic surgery | Surgical | Self-report | NC | No | P | No | 39 | 20 (51.3) |
| Solaja 2018 | 177 | Canada | Urology | Surgical | Self-report | NC | No | P | No | 35 | 33 (94.3) |
| Al Alawi 2026 | 48 | Oman | Internal medicine | Medical | Mixed | NC | Yes | R | No | 78 | NA |
| Alhaider 2015 | 51 | Saudi Arabia | Pediatrics | Medical | Self-report | NC | No | P | No | 83 | 11 (13.3) |
| Ameh 2022 | 56 | Sub-Saharan Africa | Family medicine | Medical | Self-report | NC | No | P | No | 49 | 11 (22.4) |
| Bateman 2019 | 61 | Canada | Physical medicine and rehabilitation | Medical | Database search | NC | No | P, F, R | No | 74 | 32 (43.2) |
| Brunod 2020 | 63 | France | Pediatrics | Medical | Database search | NC | No | P | No | 148 | 88 (59.5) |
| Donaldson 1996 | 78 | United Kingdom | Public health medicine | Medical | Self-report | Undefined | No | P | No | 37 | 26 (70.3) |
| Gupta 2016 | 97 | Canada | Pediatrics | Medical | Database search | NC | No | P | No | 567 | 222 (39.2) |
| Honer 2004 | 105 | Canada | Psychiatry | Medical | Mixed | NC | No | P | No | 97 | 29 (29.9) |
| Ishiguro 2019 | 109 | Japan | Pediatrics | Medical | Self-report | NC | No | P | No | 1,500 | 235 (15.7) |
| Iskhakova 2023 | 110 | Canada | Psychiatry | Medical | Program records | NC | No | P | No | 52 | 18 (34.6) |
| Jahan 2026 | 111 | Saudi Arabia | Family medicine | Medical | Mixed | NC | No | P | No | 51 | 45 (88.2) |
| Khurana 2015 | 117 | India | Neurology | Medical | Program records | NC | Yes | R | No | 11 | NA |
| Lee 2024 | 128 | Canada | Pediatrics | Medical | Self-report | NC | No | P | No | 202 | 12 (5.9) |
| Shen 2024 | 171 | Australia and New Zealand | Intensive care medicine | Medical | Database search | NC | No | P | No | 485 | 216 (44.5) |
| Smiljkovic 2021 | 175 | France | Pediatrics | Medical | Self-report | NC | No | F | No | 210 | NA |
| Soubhanneyaz 2019 | 178 | Saudi Arabia | Family medicine | Medical | Self-report | NC | No | P | No | 100 | 43 (43.0) |
| Waserman 2015 | 194 | Canada | Psychiatry | Medical | Not reported | NC | Yes | P | No | 86 | 9 (10.5) |
| Alshareefi 2026 | 55 | Kuwait | Emergency medicine | Other | Self-report | NC | No | P | No | 35 | 17 (48.6) |
| Khan 2015 | 116 | Canada | Anesthesiology | Other | Self-report | NC | No | P | No | 241 | 44 (18.3) |
| Mutsaers 2021 | 135 | Canada | Radiation oncology | Other | Database search | NC | No | P | No | 227 | 138 (60.8) |
| Patterson 2002 | 147 | Not reported | Diagnostic radiology | Other | Mixed | NC | No | P, F | No | 73 | 43 (58.9) |
| Postles 2026 | 151 | Canada | Anesthesiology | Other | Self-report | Truncated | No | P | No | 69 | 11 (15.9) |
| Quinn 2024 | 153 | Australia, New Zealand, Singapore | Radiation oncology | Other | Database search | NC | No | P, F, R | No | 168 | 118 (70.2) |
| Thiruthaneeswaran 2014 | 183 | Australia and New Zealand | Radiation oncology | Other | Mixed | NC | No | P, F | No | 116 | 83 (71.6) |
| Wang 2022 | 191 | Canada | Radiation oncology | Other | Self-report | NC | No | P | No | 53 | 35 (66.0) |
| Abri 2022 | 45 | Oman | Multiple specialties | Mixed specialties | Self-report | NC | No | P, F | No | 255 | 71 (27.8) |
| Al-Busaidi 2019 | 49 | Oman | Multiple specialties | Mixed specialties | Database search | NC | No | P | No | 552 | 43 (7.8) |
| AlEnazi 2021 | 50 | Saudi Arabia | Multiple specialties | Mixed specialties | Self-report | Undefined | No | P | No | 434 | 72 (16.6) |
| Ani 2024 | 60 | Nigeria | Multiple specialties | Mixed specialties | Self-report | NC | No | P, F | No | 352 | 124 (35.2) |
| Divya 2022 | 76 | India | Multiple specialties | Mixed specialties | Self-report | NC | No | P | No | 364 | 175 (48.1) |
| Farrokhyar 2014 | 88 | Canada | Surgical specialties | Mixed specialties | Database search | NC | Yes | P | No | 126<sup>e</sup> | 54 (42.9) |
| Nazzal 2025 | 136 | Palestine (West Bank) | Multiple specialties | Mixed specialties | Self-report | NC | No | P | No | 256 | 104 (40.6) |
| Pawar 2012 | 148 | India | Multiple specialties | Mixed specialties | Self-report | NC | No | P | No | 100 | 4 (4.0) |
| Satav 2017 | 166 | India | Multiple specialties | Mixed specialties | Self-report | NC | No | P | No | 100 | 15 (15.0) |
| Takahashi 2009 | 181 | Japan | Multiple specialties, initial postgraduate training | Mixed specialties | Self-report | NC | No | P | No | 1,094 | 82 (7.5) |
| Umar 2021 | 187 | Nigeria | Multiple specialties | Mixed specialties | Self-report | Truncated | No | P | No | 120 | 44 (36.7) |
| Yang 2025 | 200 | China | Multiple specialties | Mixed specialties | Self-report | NC | No | P | No | 546 | 82 (15.0) |
| Zhang 2025 | 201 | China | Multiple specialties | Mixed specialties | Program records | NC | No | R | No | 106 | NA |

Ref., reference number in the reference list of the main text; US, United States; NA, not applicable (the study did not report the proportion as any author); NR, not reported. Data source: database search, a search of each resident's name in a bibliographic database; program records, lists kept by the program; self-report, surveys of residents or programs.
<sup>a</sup>Training window: strict, the strict training window (start of residency to no more than 12 months after graduation); extended, publication-date window wider than the strict training window; truncated, residents still in training; partial, only some training years; undefined, not stated; unrestricted, count not limited to residency; NC, not classified. Classes were required only for the analyses of US proportions; non-US studies show a class only when one was coded during extraction (Hellenthal 2009 was coded as a US study before it was recognized as a combined US and Canadian cohort).
<sup>b</sup>Pre-intervention cohort (Pre-int.): single-program cohort measured before its residents were exposed to a research intervention.
<sup>c</sup>Data reported: P, proportion of residents with at least 1 peer-reviewed publication (any author); F, proportion with at least 1 first-author publication; R, publications per resident-year.
<sup>d</sup>Included in the US primary analysis: US studies with proportion data and at least 10 residents, excluding partial, extended, and unrestricted windows.
<sup>e</sup>Denominator restricted to residents who completed training: the report states that residents who left or did not complete training were removed from the denominator.

**Table S4.**
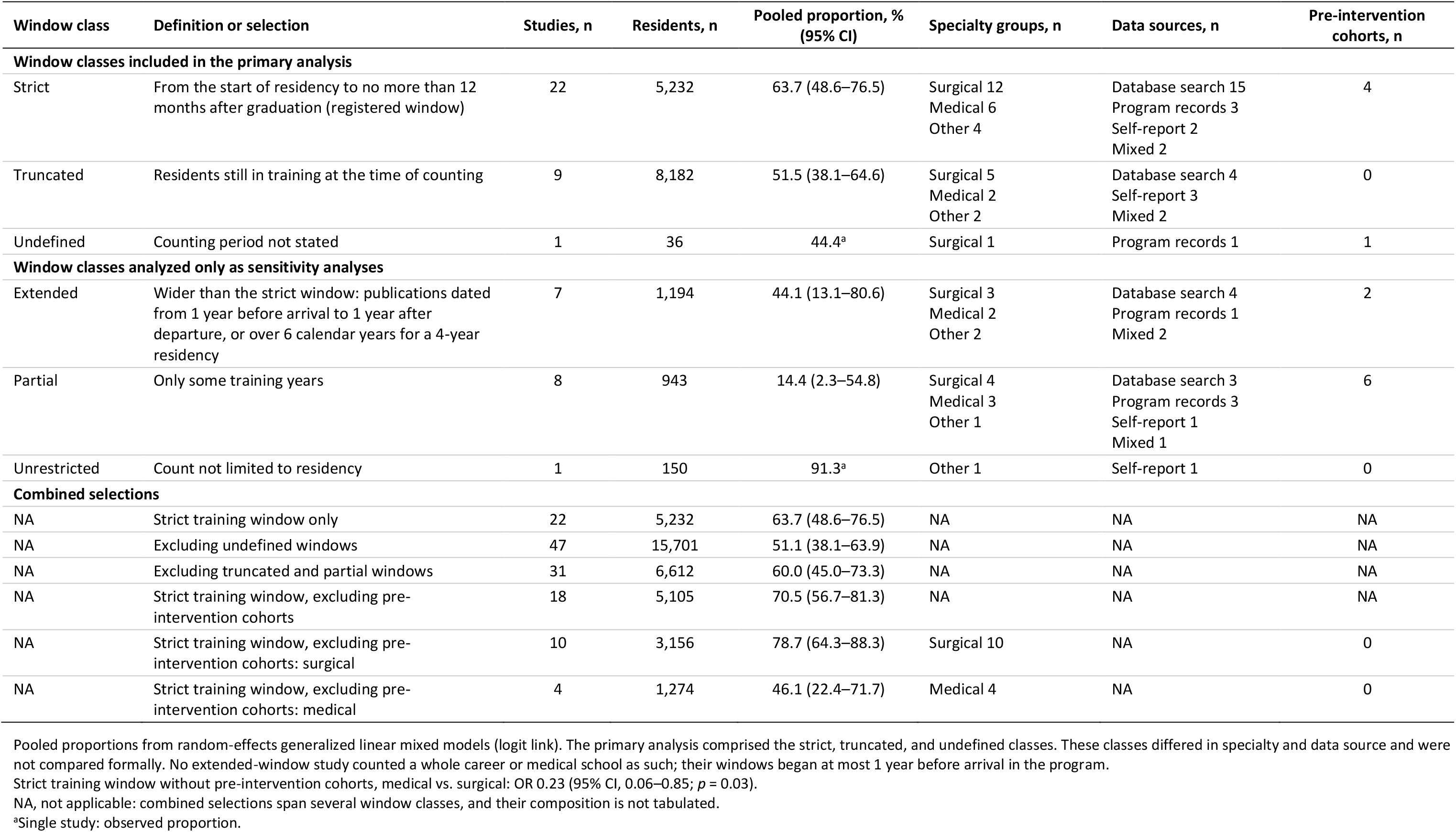
Training-window classes, pooled proportions, and study composition. US studies with proportion data: pooled proportions of residents with at least 1 peer-reviewed publication, and composition by specialty group, data source, and pre-intervention status.

| Window class | Definition or selection | Studies, n | Residents, n | Pooled proportion, % (95% CI) | Specialty groups, n | Data sources, n | Pre-intervention cohorts, n |
|---|---|---|---|---|---|---|---|
| **Window classes included in the primary analysis** |  |  |  |  |  |  |  |
| Strict | From the start of residency to no more than 12 months after graduation (registered window) | 22 | 5,232 | 63.7 (48.6–76.5) | Surgical 12<br>Medical 6<br>Other 4 | Database search 15<br>Program records 3<br>Self-report 2<br>Mixed 2 | 4 |
| Truncated | Residents still in training at the time of counting | 9 | 8,182 | 51.5 (38.1–64.6) | Surgical 5<br>Medical 2<br>Other 2 | Database search 4<br>Self-report 3<br>Mixed 2 | 0 |
| Undefined | Counting period not stated | 1 | 36 | 44.4<sup>a</sup> | Surgical 1 | Program records 1 | 1 |
| **Window classes analyzed only as sensitivity analyses** |  |  |  |  |  |  |  |
| Extended | Wider than the strict window: publications dated from 1 year before arrival to 1 year after departure, or over 6 calendar years for a 4-year residency | 7 | 1,194 | 44.1 (13.1–80.6) | Surgical 3<br>Medical 2<br>Other 2 | Database search 4<br>Program records 1<br>Mixed 2 | 2 |
| Partial | Only some training years | 8 | 943 | 14.4 (2.3–54.8) | Surgical 4<br>Medical 3<br>Other 1 | Database search 3<br>Program records 3<br>Self-report 1<br>Mixed 1 | 6 |
| Unrestricted | Count not limited to residency | 1 | 150 | 91.3<sup>a</sup> | Other 1 | Self-report 1 | 0 |
| **Combined selections** |  |  |  |  |  |  |  |
| NA | Strict training window only | 22 | 5,232 | 63.7 (48.6–76.5) | NA | NA | NA |
| NA | Excluding undefined windows | 47 | 15,701 | 51.1 (38.1–63.9) | NA | NA | NA |
| NA | Excluding truncated and partial windows | 31 | 6,612 | 60.0 (45.0–73.3) | NA | NA | NA |
| NA | Strict training window, excluding pre-intervention cohorts | 18 | 5,105 | 70.5 (56.7–81.3) | NA | NA | NA |
| NA | Strict training window, excluding pre-intervention cohorts: surgical | 10 | 3,156 | 78.7 (64.3–88.3) | Surgical 10 | NA | 0 |
| NA | Strict training window, excluding pre-intervention cohorts: medical | 4 | 1,274 | 46.1 (22.4–71.7) | Medical 4 | NA | 0 |

Pooled proportions from random-effects generalized linear mixed models (logit link). The primary analysis comprised the strict, truncated, and undefined classes. These classes differed in specialty and data source and were not compared formally. No extended-window study counted a whole career or medical school as such; their windows began at most 1 year before arrival in the program.
Strict training window without pre-intervention cohorts, medical vs. surgical: OR 0.23 (95% CI, 0.06–0.85; *p* = 0.03).
NA, not applicable: combined selections span several window classes, and their composition is not tabulated.
<sup>a</sup>Single study: observed proportion.

**Table S5.** Data extraction details for each included study. Where each value used in the analyses was found in the report of each of the 172 included studies, and what was calculated.

| Study | Details of data extraction | Adjudicated<sup>a</sup> |
|---|---|---|
| Tsyrulnik 2026 [4] | Residents assessed from Abstract and Results, paragraph 1. Residents with at least 1 publication from Results, paragraph 3. | No |
| Daly 2006 [5]<sup>b</sup> | Residents assessed from Methods (41 − 5 = 36). Residents with at least 1 publication from table 1 (0.60 × 36 = 21.6, rounded to 22). Total publications from table 1, peer-reviewed publications (1.7 × 36 = 61.2, rounded to 61). Resident-years from Results and table 1 (36 × 4 years = 144). | Values |
| Chopra 2025 [6] | Residents assessed from Abstract and Results, paragraph 1. Residents with at least 1 publication, residents with a first-author publication, and total publications from Results, during residency. | No |
| Choudhry 2023 [7]<sup>b</sup> | Residents assessed from Abstract and Results, paragraph 1. Residents with at least 1 publication from Methods, paragraph 3 (0.50 × 1,206 = 603). Total publications from table 2, all 1,206 residents (no gap year 1,036, gap year 170) (1,036 × 1.07 + 170 × 2.11 = 1,467.22, rounded to 1,467). | No |
| Wilson 2005 [8]<sup>c</sup> | Residents assessed from Results, paragraph 1 (67 − 1 = 66). Residents with at least 1 publication from Results, paragraph 1, table 1, and Abstract. Resident-years from Results (66 × 5 years = 330). | Values |
| West 2011 [16] | Residents assessed from Results (437 − 7 = 430). Residents with at least 1 publication and total publications from table 2. Resident-years from Methods (430 × 3 years = 1,290). | No |
| Sakai 2014 [17]<sup>c</sup> | Residents assessed from Results and Abstract. Residents with at least 1 publication from Results, paragraph 2, and Abstract. Total publications from Results, paragraph 2. Resident-years from not reported in the study; 220 residents x 4 years (standard length for anesthesiology) = 880. | Values |
| Al-Shaikhly 2026 [18]<sup>c</sup> | Residents assessed from Results. Residents with at least 1 publication from Results (0.4615 × 39 = 18). Residents with a first-author publication from Results. Resident-years from length not stated in the study; 39 residents x 3 years (standard length for internal medicine) = 117. | Values |
| Lakhani 2022 [28] | Residents assessed from corresponding author's reply (Dhairya A. Lakhani, October 27, 2026). Residents with at least 1 publication and total publications from corresponding author's reply. Resident-years from corresponding author's reply (18 + 20 residents enrolled in the 2 pre-intervention years = 38 resident-years). | No |
| Gosnell 2024 [29] | Residents assessed from corresponding author's reply (Griffith G. Gosnell, October 27, 2026). Residents with at least 1 publication from corresponding author's reply. | No |
| Raymond 2024 [30]<sup>b</sup> | Total publications from Results (5.3 × 7 = 37.1, rounded to 37). Resident-years from corresponding author's reply (Caitlin Raymond, October 28, 2026) (19 + 19 + 20 + 21/2 = 68.5). | No |
| Fereydooni 2024 [31]<sup>c</sup> | Residents assessed from corresponding author's reply (Arash Fereydooni, October 27, 2026). Residents with at least 1 publication and total publications from corresponding author's reply. Resident-years from assumed (131 × 4 years = 524). | No |
| Stultz 2024 [32] | Residents assessed from Results, paragraph 1, and table 1. Residents with at least 1 publication from table 2 (204 + 103 + 123 = 430). Resident-years from Methods (522 × 3 years = 1,566). | Values |
| Waheed 2025 [40] | Residents assessed from Results, paragraph 1. Residents with at least 1 publication from table 3. Residents with a first-author publication from table 3, all manuscripts. Total publications from table 3. Resident-years from table 1 (14 × 3 years = 42). | No |
| Abramson 2018 [44] | Residents assessed from Results and table 3. Residents with at least 1 publication from Results (13.4% (n = 62) had a manuscript accepted). | No |
| Abri 2022 [45] | Residents assessed, residents with at least 1 publication, and residents with a first-author publication from table 2. | No |
| Acevedo 2024 [46]<sup>c</sup> | Residents assessed from Methods, paragraph 3, and Results. Residents with at least 1 publication from Results, paragraph 1 (156 − 55 = 101). Total publications from figure 1 (bar chart, read from the figure). Resident-years from Methods, paragraph 3 (156 × 6 years = 936). | No |
| Ahmad 2026 [47]<sup>b</sup> | Residents assessed from Methods, paragraph 1. Residents with at least 1 publication and residents with a first-author publication from Results, paragraph 1. Total publications from Results, paragraph 1 (4.95 × 112 = 554.4, rounded to 554). | Values |
| Al Alawi 2026 [48] | Residents assessed from Methods and table 1. Total publications from table 1 (9 publications in 2020/21 + 12 in 2021/22 = 21). Resident-years from Methods (78 × 2 = 156). | No |
| Al-Busaidi 2019 [49] | Residents assessed from Abstract, Results, and table 1. Residents with at least 1 publication from Abstract and table 1. Total publications from Abstract and Results. | No |
| AlEnazi 2021 [50] | Residents assessed from Results, paragraph 1. Residents with at least 1 publication from Results. | No |
| Alhaider 2015 [51] | Residents assessed from Results, paragraph 1, and table 1. Residents with at least 1 publication from Results and Abstract (83 × 0.867 = 72 never, 83 − 72 = 11). | No |
| Alkhamees 2021 [52] | Residents assessed from Results, paragraph 1, and table 1. Residents with at least 1 publication from table 4 (60 + 17 + 22 = 99). | No |
| Allen 2017 [53]<sup>b</sup> | Total publications from Results, paragraph 2 (0.30 × 99 = 29.7, rounded to 30). Resident-years from table 1 (18 + 18 + 21 + 20 + 22 = 99). | No |
| Al-Mohrej 2021 [54]<sup>b</sup> | Residents assessed from Results, paragraph 1, and table 1. Total publications from Results (1.14 × 128 = 145.9, rounded to 146). Resident-years from table 1, training level (22 × 1 + 36 × 2 + 30 × 3 + 26 × 4 + 14 × 5 = 358). | Eligibility |
| Alshareefi 2026 [55] | Residents assessed from table 2 (13 men + 22 women = 35 who answered the publication item (40 completers)). Residents with at least 1 publication from table 2 (published a project, 3 of 13 men + 14 of 22 women = 17). | Eligibility |
| Ameh 2022 [56] | Residents assessed from tables 1 and 2. Residents with at least 1 publication from table 2. | No |
| Anderson 2021 [57] | Residents assessed from Results and table 1. Total publications from table 2. Resident-years from Methods, data analysis (296 × 4 years = 1,184). | Values |
| Andrews 2013 [58] | Residents assessed from Results, paragraph 1, and table 1. Residents with at least 1 publication from Results and Abstract. Residents with a first-author publication from Results (0.38 × 42 = 15.96, rounded to 16). | No |
| Angerett 2024 [59] | Total publications from Results and table 1. Resident-years from Methods (25 × 5 = 125). | No |
| Ani 2024 [60] | Residents assessed from Results, paragraph 1, and table 1. Residents with at least 1 publication and residents with a first-author publication from table 3. | No |
| Bateman 2019 [61] | Residents assessed from Abstract and Results, paragraph 1. Residents with at least 1 publication from Results, paragraph 1 (43.2% (n = 32) had at least 1 publication). Residents with a first-author publication from Results, paragraph 1. Total publications from Results, paragraph 1, and Abstract. Resident-years from Methods (74 × 5 years = 370). | No |
| Brown 2025 [62]<sup>c</sup> | Residents assessed from Abstract and Results (185 − 13 = 172). Total publications from table 2. Resident-years from table 1 (172 × 4 years = 688). | No |
| Brunod 2020 [63] | Residents assessed from Results, paragraph 1, and Abstract. Residents with at least 1 publication from Results, paragraph 2, and Abstract. Total publications from Results, paragraph 2, and table 1. | Values |
| Carr 2022 [64] | Residents assessed from Abstract and Results, paragraph 1. Total publications from table 2, during residency training. Resident-years from Methods, data analysis (341 × 5 years = 1,705). | Values |
| Carter 2019 [65] | Residents assessed from Abstract, Results, and table 2. Residents with at least 1 publication from table 2 and Results (0.206 × 63 = 12.98, rounded to 13). Resident-years from Methods (63 × 3 years = 189). | Values |
| Castillo-Rangel 2026 [66] | Residents assessed from Results, paragraph 1. Residents with at least 1 publication from Results (0.491 × 511 = 251 without, 511 − 251 = 260). | No |
| Chan 2009 [67] | Residents assessed from Abstract and Results. Residents with at least 1 publication from Discussion (85 − 62 = 23). Total publications from Results. | No |
| Chan 2021 [68] | Residents assessed from Abstract and table 2. Residents with at least 1 publication from Abstract and Results, paragraph 2. | No |
| Chang 2013 [69]<sup>b</sup> | Total publications from Results (0.13 × 70 resident-years (10 residents × 7 academic years, 1998-2004) = 9.1, rounded to 9). Resident-years from Methods, paragraph 4 (10 × 7 = 70). | No |
| Chen 2015 [70] | Residents assessed and residents with at least 1 publication from Results and table 1. Total publications from Results and Abstract. Resident-years from Methods (64 × 5 + 11 × 7 = 397). | No |
| Chhor 2022 [71] | Total publications from Results, paragraph 1, and table 1. Resident-years from Results, paragraph 1. | Values |
| Cocci 2018 [72] | Residents assessed from Abstract and Results. Residents with at least 1 publication from Results and Abstract (324 − 104 = 220). | No |
| Crowley 2012 [73]<sup>bc</sup> | Residents assessed from Results and table 1. Residents with at least 1 publication from Results (717 − 219 = 498). Residents with a first-author publication from Results (717 − 377 = 340). Total publications from Results, Publications (717 × 2.5 = 1,792.5, rounded to 1,792 (half rounded down)). Resident-years from not reported in the study; 717 residents x 7 years (standard length for neurological surgery) = 5,019. | Values |
| Değer 2022 [74]<sup>bc</sup> | Residents assessed from Results and table 1. Residents with a first-author publication from table 2 (at least 1 first-authored publication, 21 + 16 = 37). Total publications from table 2 (3.21 × 100 + 6.41 × 49 = 635.1, rounded to 635). Resident-years from length not stated in the study; 149 residents x 5 years (national length of urology residency in Turkey) = 745. | Eligibility |
| Demirtaş 2020 [75]<sup>bc</sup> | Residents assessed from Results, paragraph 1. Total publications from table 7 (94 × 1.77 + 22 × 3.77 = 249.3, rounded to 249). Resident-years from length not stated in the study; 116 residents x 5 years (national length of orthopaedic residency in Turkey) = 580. | Eligibility |
| Divya 2022 [76] | Residents assessed from Abstract and Results, paragraph 1. Residents with at least 1 publication from Results and Discussion (48% (n = 175) had publications). | No |
| Doman 2022 [77] | Residents assessed and residents with a first-author publication from table 1. Total publications from table 1 and Results, paragraph 1. Resident-years from table 1 (32 × 1 = 32). | No |
| Donaldson 1996 [78] | Residents assessed from Results and tables 1 and 2 (38 − 1 = 37). Residents with at least 1 publication from table 1. | Eligibility |
| Donley 2024 [79]<sup>bc</sup> | Residents assessed from Results, paragraph 1, and table 1. Residents with at least 1 publication from table 1 and Results, paragraph 2 (758 − 129 = 629). Total publications from table I, publications per resident during residency (5.8 × 758 = 4,396.4, rounded to 4,396). Resident-years from Methods (758 × 5 = 3,790). | No |
| Dunn 1998 [80]<sup>c</sup> | Residents assessed from Methods. Total publications from table 1, by prior research (49 residents × 9.5 + 46 residents × 7.2 = 796.7, rounded to 797 (95 of the 96 respondents had data)). Resident-years from Results (96 × (5 years standard for general surgery + 1 laboratory year) = 576). | Both |
| Durning 2004 [81]<sup>c</sup> | Residents assessed from table 1 and Results. Residents with at least 1 publication from Results (table 2 resident authors 1 + 1 + 1 + 1 + 2 + 1 = 7). Total publications from tables 1 and 2. Resident-years from individual exposure not reported in the study; 46 residents x 3 years (standard length for internal medicine) = 138. This remains an assumed denominator, not verified resident-level exposure. | Values |
| Eckert 2016 [82] | Residents assessed from table 3 (class sizes before the program, 14 + 14 + 11 = 39). Residents with at least 1 publication from table 3 (residents who published, 2 + 1 + 2 = 5). Total publications from table 3 (peer-reviewed publications, 4 + 6 + 2 = 12). | No |
| Elliott 2009 [83]<sup>c</sup> | Residents assessed from Methods and Abstract. Total publications from Results and Abstract. Resident-years from Methods (95 × 5 years = 475). | No |
| Emerick 2013 [84] | Residents assessed from Results and table 4 (10 + 11 + 19 + 14 + 9 + 15 + 15 + 13 = 106). Total publications from table 4 (manuscripts, 2 + 3 + 11 + 6 + 1 + 5 + 4 + 18 = 50). Resident-years from Methods (106 × 4 = 424). | Values |
| Eyigör 2021 [85] | Residents assessed from Results, paragraph 1. Residents with at least 1 publication from Discussion and table 2 (0.395 × 119 = 47). | Eligibility |
| Fan 2014 [86]<sup>dbc</sup> | Residents assessed from Methods and Results. Total publications from table 1 and Results (1.18 × 18 + 0.34 × 42 = 35.52). Resident-years from not reported in the study; 60 residents x 5 years (standard length for orthopaedic surgery) = 300. | Values |
| Fanciullo 2018 [87]<sup>c</sup> | Residents assessed from Results, paragraph 1, and table 2. Residents with at least 1 publication and total publications from Abstract, Results, and table 2. Resident-years from Methods (47 residents × 3 years = 141. This calculation assumes all 47 residents contributed the full period). | Values |
| Farrokhyar 2014 [88] | Residents assessed from Results (133 − 7 = 126). Residents with at least 1 publication from Results. | No |
| Fladie 2023 [89]<sup>c</sup> | Residents assessed from Abstract, Results, and table 1. Total publications from Results and table 3, by fellowship (327 graduates × 4.7 + 78 graduates × 1.5 = 1,653.9, rounded to 1,654). Resident-years from length not stated in the study; 405 x 5 years (standard length for general surgery) = 2,025. | Values |
| Fliotsos 2021 [90]<sup>bc</sup> | Residents assessed from Abstract, Results, and table 1. Total publications from table 1, publications during residency (4 residents with parental leave × 4.8 + 21 without leave × 3.7 = 96.9, rounded to 97). Resident-years from Discussion (25 × 3 years = 75). | No |
| Forrester 2017 [91] | Residents assessed from Abstract and Results, paragraph 2. Residents with a first-author publication from Results. | Values |
| Gaught 2013 [92]<sup>c</sup> | Residents assessed from Abstract and Methods. Residents with at least 1 publication from Results, paragraph 1 (publication rate 18.2% (n = 119)). Total publications from figure 1 legend. Resident-years from not reported in the study; 654 x 5 years (standard length of 4 years for physical medicine and rehabilitation + 1 year for the extended window) = 3,270. | No |
| Gomez 2025 [93]<sup>bc</sup> | Residents assessed from Results, paragraph 1. Total publications from Results, paragraph 2, and figure 3 (7.1 × 186 = 1,320.6, rounded to 1,321). Resident-years from Methods (186 × 6 = 1,116). | Eligibility |
| Gottlieb 2021 [94] | Residents assessed from Results, paragraph 1, and table 1. Residents with at least 1 publication from Results, paragraph 1. Total publications from Results, paragraph 1, and table 2. Resident-years from Methods (11 × 1 + 12 × 2 = 35). | Values |
| Green 2025 [95] | Residents assessed from Results, paragraph 1, and table 1 (826 nonacademic + 166 academic = 992 graduates). Residents with a first-author publication from Results, paragraph 1. | Both |
| Guo 2021 [96]<sup>c</sup> | Residents assessed from Results, paragraph 1. Total publications from Results, paragraph 1, and table 1. Resident-years from Methods (502 × 3.5 = 1,757). | Values |
| Gupta 2016 [97] | Residents assessed from Abstract and Results. Residents with at least 1 publication from Abstract, Results, and table 2 (published during residency, 93 + 129 = 222). Resident-years from Methods (567 × 4 years = 2,268). | No |
| Gupta 2023 [98] | Residents assessed from Results, paragraph 1. Total publications from table 1, publications during PGY-2 to PGY-4 by study type; 89 + 40 + 1,092 + 697 + 275 + 436 = 2,629. Resident-years from Methods and table 1 footnote (770 × 3 years = 2,310). | Values |
| Gutowski 2026 [99]<sup>bc</sup> | Residents assessed from Abstract and Results, paragraph 1. Total publications from Results (8.7 × 277 = 2,409.9, rounded to 2,410). Resident-years from Methods (277 × 6 years = 1,662). | Both |
| Hang 2022 [100] | Residents assessed from Abstract and Results, paragraph 1. Residents with at least 1 publication from Abstract and Results (100 − 37 = 63). Total publications from Results, paragraph 1 (429 − 237 = 192). | Values |
| Harrison 2020 [101] | Total publications from table 2 and Results, paragraph 2. Resident-years from table 2. | No |
| Hellenthal 2009 [102] | Residents assessed from Results and Abstract. Residents with at least 1 publication from Results. Residents with a first-author publication from Introduction and Results. Total publications from Results and figure 1. | No |
| Herczeg 2025 [103]<sup>c</sup> | Residents assessed from table 1. Total publications from table 2. Resident-years from table 1, residents by PGY level (844 × 1 + 829 × 2 + 838 × 3 + 825 × 4 + 9 × 5 = 8,361). | Values |
| Hoedebecke 2014 [104] | Residents assessed from Introduction. Total publications from Introduction and Abstract. Resident-years from Introduction (24 × 1 = 24). | No |
| Honer 2004 [105] | Residents assessed and residents with at least 1 publication from Results. | No |
| Huang 2022 [106]<sup>b</sup> | Residents assessed from Abstract, Results, and table 1. Residents with a first-author publication from figure 1A (bar chart, read from the figure) (418 − 212 = 206). Total publications from table 1 (2.68 × 418 = 1,120.24, rounded to 1,120). Resident-years from Methods (418 × 3 years = 1,254). | Values |
| Huang 2023 [107]<sup>c</sup> | Residents assessed from Abstract, Methods, and tables 1 and 2. Residents with at least 1 publication from table 2 (20 − 2 = 18). Residents with a first-author publication from table 1 (20 − 5 = 15). Total publications from Results and Abstract. Resident-years from not reported in the study; 20 residents x 5 years (standard length for radiation oncology) = 100. | No |
| Huh 2022 [108] | Residents assessed from table 2 (residents without leave with publication data, yes 165 + no 72 = 237). Residents with at least 1 publication from table 2. Resident-years from Methods (residents who did not take parental leave, table 2: 165 + 72 = 237; 237 × 3 = 711). | Values |
| Ishiguro 2019 [109] | Residents assessed from Abstract and Results, paragraph 1. Residents with at least 1 publication from Results, paragraph 3, and table 1. Resident-years from Results, paragraph 1 (1,500 × 3.9 = 5,850). | No |
| Iskhakova 2023 [110] | Residents assessed from Outcomes and Abstract. Residents with at least 1 publication from Outcomes. Resident-years from Outcomes (52 × 5 years = 260). | Eligibility |
| Jahan 2026 [111] | Residents assessed from Results (cohorts of 12 + 17 + 22 = 51 residents). Residents with at least 1 publication from Results and Abstract. Total publications from Results. | Eligibility |
| Jinka 2023 [112]<sup>b</sup> | Residents assessed from Results, paragraph 1, and Abstract. Total publications from Results, paragraph 1 (3.1 × 655 = 2,030.5, rounded to 2,031). Resident-years from table 4, residency length including research years (147 × 5 + 458 × 6 + 41 × 7 + 9 × 8 = 3,842). | Values |
| Johnson 2018 [113]<sup>c</sup> | Total publications from Results (286 + 512 = 798). Resident-years from Results (286 / 0.290 = 986). | Values |
| Johnson 2021 [114]<sup>b</sup> | Residents assessed from Abstract and Results, paragraph 1. Total publications from Results (3.5 × 249 = 871.5, rounded to 872). Resident-years from Methods (249 × 5 years = 1,245). | Values |
| Jordan 2021 [115]<sup>b</sup> | Residents assessed from Results, paragraph 1, and Abstract. Total publications from Results (93 chief residents × mean 1.60 + 272 other graduates × mean 0.83 = 374.56, rounded to 375). Resident-years from Results, paragraph 1 (167 × 3 + 198 × 4 = 1,293). | Values |
| Khan 2015 [116] | Residents assessed from Results (244 − 3 = 241). Residents with at least 1 publication from Results and table 2. | No |
| Khurana 2015 [117] | Residents assessed from Methods. Total publications from tables 1 and 2. Resident-years from Methods (11 × 24 / 12 = 22). | No |
| Kirsner 1999 [118]<sup>c</sup> | Residents assessed from text, page 401, and table 2. Total publications from table *I* (articles published by the 43 dermatology residents graduating 1988-1995, total N = 148). Resident-years from page 400 (43 × 3 years = 129). | Both |
| Kohlert 2017 [119]<sup>b</sup> | Residents assessed from Abstract and Results. Residents with at least 1 publication from Results (312 − 83 = 229). Total publications from table 3 and Results (3.35 × 312 = 1,045.2, rounded to 1,045). Resident-years from Methods (312 × 5 years = 1,560). | Values |
| Kohlwes 2016 [120] | Residents assessed from table 2 footnote. Residents with a first-author publication from Methods. | Values |
| Koo 2022 [121]<sup>b</sup> | Residents assessed from Results and table 1. Total publications from table 1 (0.56 × 45 = 25.2, rounded to 25). Resident-years from Methods (45 residents × 1 year = 45). | No |
| Koroulakis 2020 [122] | Residents assessed from table 1 and Results, paragraph 1. Residents with at least 1 publication from table 1 (150 − 13 = 137). | Eligibility |
| Kreitz 2019 [123] | Residents assessed from Abstract and Results, paragraph 1. Residents with at least 1 publication from Methods and figure 1 legend (0.65 × 274 = 178.1, rounded to 178). Residents with a first-author publication from Methods (0.49 × 274 = 134.3, rounded to 134). | No |
| Krispin 2025 [124] | Residents assessed from Results, paragraph 1 (90 − 43 = 47). Total publications from Results, paragraph 2. Resident-years from Methods (47 × 3 years = 141). | No |
| Krueger 2017 [125] | Residents assessed from table 2, third program (no protected research time) (0 + 5 + 3 + 40 + 0 = 48 residents). Residents with at least 1 publication from table 2, third program (no protected research time) (48 − 0 = 48). | No |
| Kryshtalskyj 2024 [126] | Residents assessed and residents with at least 1 publication from Abstract, Results, and table 1. Total publications from Results. Resident-years from Introduction (320 × 5 years = 1,600). | No |
| Larsen 2020 [127] | Total publications from Results, paragraph 1. Resident-years from Methods (10 × 9 = 90). | Values |
| Lee 2024 [128] | Residents assessed and residents with at least 1 publication from Results. | No |
| Macknin 2014 [129]<sup>b</sup> | Residents assessed from Methods. Residents with at least 1 publication from Results (0.41 × 74 = 30.3, rounded to 30). Total publications from figure 4 (bar label, read from the figure) (0.9 × 74 = 66.6, rounded to 67). Resident-years from Methods (74 × 5 = 370). | No |
| Manring 2014 [130]<sup>c</sup> | Residents with at least 1 publication and total publications from Results. Resident-years from Methods, paragraph 1 (30 × 1 year = 30). | Eligibility |
| Mansi 2019 [131] | Residents assessed from table 3. Residents with a first-author publication from Results (0.6897 × 29 = 20.0, rounded to 20). | Values |
| Merani 2014 [132] | Residents assessed from table, total row, in-training column, and Results. Residents with at least 1 publication and residents with a first-author publication from table, total row, in training. | No |
| Millis 2004 [133]<sup>bc</sup> | Residents assessed from Methods, page 429. Total publications from Results, page 430 (0.71 × 288 = 204.5, rounded to 204). Resident-years from page 430 (96 × 3 years = 288). | Eligibility |
| Motiwala 2024 [134]<sup>c</sup> | Residents assessed from Results and table 1. Residents with at least 1 publication from figure 1A (dot plot, read from the figure) (19 − 8 = 11). Total publications from Results. Resident-years from not reported in the study; 19 residents x 7 years (standard length for neurological surgery) = 133. | Values |
| Mutsaers 2021 [135] | Residents assessed from Abstract and Results. Residents with at least 1 publication from Results (0.61 × 227 = 138.5, rounded to 138). Total publications from Abstract and Results. | No |
| Nazzal 2025 [136] | Residents assessed from Results, paragraph 1, and table 1. Residents with at least 1 publication from table 3. | No |
| Ogunyemi 2010 [137] | Residents assessed from Discussion, page 96. Residents with at least 1 publication from Discussion, page 96 (0.10 × 20 = 2). Resident-years from Discussion (20 × 1 = 20). | Eligibility |
| Ohori 2016 [138]<sup>bc</sup> | Residents assessed from Results, paragraph 1, and table 1 (30 + 34 + 36 + 39 = 139). Total publications from table 1, publications per resident (30 × 1.43 + 34 × 2.11 + 36 × 3.64 + 39 × 3.92 = 398.56, rounded to 399). Resident-years from table 1 training types and Discussion lengths (anatomic or clinical pathology 3 or 4 years, combined training 4 or 5 years, by period); 148 + 164 + 136 + 143 = 591. | Values |
| Olanipekun 2023 [139] | Residents assessed from Results, paragraph 1, and table 2. Residents with at least 1 publication from table 2. | No |
| Oliver 2019 [140] | Residents assessed from Abstract and Methods. Total publications from Abstract and Results. Resident-years from Methods (16 × 3 years = 48). | No |
| Ong 2023 [141]<sup>c</sup> | Residents assessed from Abstract, Results, and table 1 (303 women + 1,103 men = 1,406). Total publications from table 1, publications during residency (1,686 (women) + 11,365 (men) = 13,051). Resident-years from table 1, residents by PGY level (1 woman in an additional year not counted) (1,050 (women) + 4,484 (men) = 5,534). | No |
| Onyekaba 2022 [142]<sup>b</sup> | Residents assessed from table 1 and Results. Total publications from Abstract (15.9 × 423 = 6,725.7, rounded to 6,726). Resident-years from table 1, 6-year category (423 × 6 = 2,538). | Values |
| Osborn 2018 [143]<sup>b</sup> | Residents assessed from table 2b, the program with no dedicated research time (0 + 1 + 0 + 18 + 12 = 31 residents). Residents with at least 1 publication from table 2b, the program with no dedicated research time (31 − 12 = 19). Total publications from table 2b, the program with no dedicated research time (2.10 × 31 = 65.1, rounded to 65). | No |
| Panchal 2014 [144] | Total publications from table 1, peer-reviewed articles before the program (2 + 1 + 3 = 6). Resident-years from Results, paragraph 1. | Values |
| Papasavas 2013 [145] | Residents assessed from table 1, before the mandatory research requirement (July 2008-July 2010) (annual categorical resident counts 31 + 29 = 60 (unique residents not reported)). Residents with at least 1 publication from Results. Total publications from table 1 and Results, paragraph 3. Resident-years from table 1, categorical residents (31 in 2008-2009 + 29 in 2009-2010 = 60). | Values |
| Patel 2018 [146]<sup>c</sup> | Residents assessed from Methods and Results. Residents with at least 1 publication from Results (0.44 × 36 = 15.8, rounded to 16). Total publications from Results. Resident-years from length not stated in the study; 36 residents x 5 years (standard length for general surgery) = 180. | Values |
| Patterson 2002 [147]<sup>b</sup> | Residents assessed from Methods and Abstract. Residents with at least 1 publication from Results. Residents with a first-author publication from Results and table 3. Total publications from Results (43 × 2.00 = 86). | No |
| Pawar 2012 [148] | Residents assessed from Abstract and Methods. Residents with at least 1 publication from Abstract and Results (0.04 × 100 = 4). | No |
| Persad 2023 [149] | Residents assessed from Abstract and Results, paragraph 1. Residents with at least 1 publication from Results. | No |
| Plackett 2020 [150]<sup>b</sup> | Residents assessed from Results, paragraph 5. Residents with at least 1 publication from Results, paragraph 5, and Abstract. Total publications from Results, paragraph 5, and figure 1 (18 × 4.00 = 72). Resident-years from Methods (18 × 6 = 108). | Values |
| Postles 2026 [151] | Residents assessed from Abstract and Results, paragraph 1. Residents with at least 1 publication from table 1 (first or coauthor of a publication, 2 + 0 + 4 + 5 + 0 = 11 (16%)). | No |
| Pur 2024 [152]<sup>b</sup> | Residents assessed from Results, paragraph 1, and table 1. Residents with at least 1 publication from table 1. Total publications from Results (187 × 5.17 + 182 × 1.60 = 1,258). | No |
| Quinn 2024 [153] | Residents assessed from Abstract and Results, paragraph 1 (191 − 23 = 168). Residents with at least 1 publication from Abstract and table 5. Residents with a first-author publication from Abstract and table 4. Total publications from Results, paragraph 2, and tables 2 and 3. Resident-years from Methods (168 × 5 years = 840). | Values |
| Rajih 2025 [154] | Residents assessed from Results, paragraph 1, and table 1 (306 − 9 = 297). Residents with at least 1 publication from table 2. | No |
| Ravi 2021 [155] | Residents assessed from Results, demographics. Residents with at least 1 publication from Results. Residents with a first-author publication from Results, Impact. Total publications from Results. | Values |
| Reddy 2023 [156]<sup>bc</sup> | Residents assessed from Methods, page 104. Total publications from Results, page 104 (12.3 × 54 = 664.2, rounded to 664). Resident-years from length not stated in the study; 54 residents x 6 years (standard length for integrated plastic surgery) = 324. | No |
| Resorlu 2016 [157] | Residents assessed from Abstract, Methods, and Results. Residents with at least 1 publication and residents with a first-author publication from Results, page 32. | No |
| Reyes 2023 [158]<sup>b</sup> | Residents assessed from Results, paragraph 1 (201 + 222 + 244 + 297 = 964). Residents with at least 1 publication from Results, paragraph 2. Total publications from table 1, intraresidency publications (2.03 × 964 = 1,956.92, rounded to 1,957). Resident-years from Methods, Classification (964 × 3 years = 2,892). | No |
| Robbins 2013 [159] | Residents assessed from table 3, page 650, residents in the classes of 1998-2005 (8 + 8 + 8 + 7 + 10 + 7 + 8 + 12 = 68). Total publications from table 3, page 650, publications per class 1998-2005 (16 + 9 + 15 + 7 + 5 + 15 + 27 + 16 = 110). Resident-years from Methods (68 × 5 years = 340). | Values |
| Robbins 2017 [160]<sup>b</sup> | Residents assessed from Results. Total publications from table 2, peer-reviewed publications (0.75 × 53 = 39.75, rounded to 40). Resident-years from Methods (53 × 3 years = 159). | No |
| Romanos-Sirakis 2020 [161] | Residents assessed from Setting and participants (8 + 8 = 16). Residents with a first-author publication and total publications from Abstract. Resident-years from Outcome measures (8 + 16 = 24). | Values |
| Rothberg 2014 [162]<sup>c</sup> | Residents with a first-author publication from table 2, resident first author. Total publications from table 2 and Abstract. Resident-years from length not stated in the study; 85 PGY-3 residents x 3 years (standard length for internal medicine) = 255. | Both |
| Rowley 2021 [163] | Residents assessed from Results. Residents with a first-author publication from Discussion (909 × (1 − 0.233) = 697.2, rounded to 697). Total publications from Results. Resident-years from Methods (909 × 4 = 3,636). | Values |
| Sanpan 2022 [164] | Residents assessed from Results. Residents with at least 1 publication from Results and table 1. Total publications from Results. Resident-years from Introduction (102 × 3 years = 306). | No |
| Sarica 2020 [165] | Residents assessed from Abstract and Results, page 674. Residents with at least 1 publication from Abstract and figure 1B (254 + 53 + 16 + 22 + 2 = 347). | No |
| Satav 2017 [166] | Residents assessed from Abstract and Results (102 − 2 = 100). Residents with at least 1 publication from table 3 and Abstract (0.15 × 100 = 15). | No |
| Seaburg 2016 [167] | Residents assessed from Abstract, Methods, and table 1. Total publications from Abstract, Results, and table 1. Resident-years from Methods (308 × 3 years = 924). | Values |
| Seehusen 2009 [168] | Total publications from Evaluation, page 468. Resident-years from Evaluation, page 468 (18 × 4 = 72). | Values |
| Shah Mardan 2020 [169] | Residents assessed from Methods (40 − 1 = 39). Residents with at least 1 publication from table 2, publications during residency (14 + 4 + 2 = 20). | No |
| Sharpe 2025 [170] | Residents assessed and residents with at least 1 publication from Results, page 2. | Values |
| Shen 2024 [171] | Residents assessed and residents with at least 1 publication from Abstract and Results. Total publications from Results. | Values |
| Sheppard 2021 [172]<sup>bc</sup> | Residents assessed from tables 1 and 2. Total publications from table 2, 2011 match (14.7 × 145 = 2,131.5, rounded to 2,132). Resident-years from table 1 (145 × 7 = 1,015). | Both |
| Shi 2018 [173] | Residents assessed from Results and table 1. Residents with at least 1 publication from table 2 (at least 1 publication during residency, 112 in academic + 215 in private practice = 327). Total publications from Results, paragraph 1. Resident-years from Methods (392 graduates × 3 years = 1,176). | Values |
| Sinha 2026 [174]<sup>bc</sup> | Residents assessed from Results, paragraph 1, and tables 1 and 2. Total publications from table 2, integrated graduates 2020-2024, during residency (12.41 × 745 = 9,245.45, rounded to 9,245). Resident-years from length not stated in the study; 745 x 6 years (standard length for integrated plastic surgery) = 4,470. | Eligibility |
| Smiljkovic 2021 [175] | Residents assessed from Results, paragraph 1. Residents with a first-author publication from Results, paragraph 2 (0.11 × 210 = 23.1, rounded to 23). | Values |
| Smith 2014 [176]<sup>c</sup> | Residents assessed from table 1. Total publications from table 3, no laboratory time (1.29 × 24 = 30.96, rounded to 31). Resident-years from length of clinical training not stated; 68 x 5 years (standard length for general surgery) + 90 laboratory years from table 1 = 430. | Both |
| Solaja 2018 [177]<sup>c</sup> | Residents assessed from Methods and Results. Residents with at least 1 publication from Abstract and Results, paragraph 2 (0.94 × 35 = 32.9, rounded to 33). Resident-years from length not stated in the study; 35 x 5 years (standard length for urology in Canada) = 175. | Values |
| Soubhanneyaz 2019 [178] | Residents assessed from Results and table 1. Residents with at least 1 publication from table 3 and Results. | Eligibility |
| Suleta 2026 [179] | Residents assessed from Abstract and Methods. Residents with at least 1 publication from Results, paragraph 3, and table 1. Residents with a first-author publication from Results. Total publications from table 1. Resident-years from Methods (30 × 1 = 30). | No |
| Sullivan 2024 [180] | Total publications from Results, page 902 (8 + 8 + 9 = 25). Resident-years from Introduction (20 × 3 = 60). | Values |
| Takahashi 2009 [181] | Residents assessed from Results. Residents with at least 1 publication from Results, paragraph 2. Resident-years from Results (1,094 × 2 years = 2,188). | Values |
| Taniguchi 1994 [182] | Residents assessed from Abstract and Results, page 241. Residents with at least 1 publication from Abstract, page 240, and figure 1. | No |
| Thiruthaneeswaran 2014 [183] | Residents assessed from Methods and Results, paragraph 1. Residents with at least 1 publication from Results. Residents with a first-author publication from Results and figure 1 (7 + 9 + 14 + 12 + 8 + 12 = 62). | No |
| Thomas 2000 [184]<sup>b</sup> | Residents assessed from Methods, page 136, and table 1. Residents with at least 1 publication from figure 1 (bar chart, read from the figure) (control group (traditional 5-year residency) nonzero bars 9 + 1 + 2 + 3 + 0 = 15 residents with at least 1 publication). Total publications from table 1 and Results, page 136 (0.94 × 31 = 29.14, rounded to 29). Resident-years from Methods, page 136, and Abstract (31 × 5 years = 155). | Values |
| Thompson 2018 [185]<sup>b</sup> | Residents assessed from Abstract, Results, and table 1. Residents with at least 1 publication from Abstract and Results. Total publications from Abstract and Results (9.0 × 49 = 441). Resident-years from Methods and table 1 (34 × 5 + 15 × 6 = 260). | No |
| Torres 2015 [186]<sup>c</sup> | Residents assessed from Abstract and Methods. Total publications from Results and table 2. Resident-years from not reported in the study; 24 residents x 5 years (standard length for orthopaedic surgery) = 120. | No |
| Umar 2021 [187] | Residents assessed from Abstract and Methods. Residents with at least 1 publication from Abstract and Results. Total publications from Results. | Values |
| Valikodath 2017 [188]<sup>b</sup> | Residents assessed from Methods and tables 1 and 2. Total publications from table 2 (2.00 × 37 = 74). Resident-years from Methods (37 × 3 years = 111). | No |
| Waheed 2020 [189] | Residents assessed from Results, paragraph 1, and table 3. Total publications from table 3 and Discussion. Resident-years from table 1 (16 × 3 years = 48). | No |
| Waheed 2024 [190] | Residents assessed from Results, paragraph 1, and table 1. Total publications from table 2. Resident-years from Results (20 × 3 years = 60). | Values |
| Wang 2022 [191] | Residents assessed from Abstract and Results. Residents with at least 1 publication from Abstract (0.66 × 53 = 34.98, rounded to 35). | No |
| Wang 2023 [192]<sup>bc</sup> | Residents assessed from Results and table 2. Total publications from table 2 (5.4 × 71 = 383.4, rounded to 383). Resident-years from Methods (71 × 5 years = 355). | Both |
| Warren 2025 [193]<sup>bc</sup> | Residents assessed from Results and tables 1a and 1b. Residents with at least 1 publication from figure 2B (228 × 0.0746 = 17.0, rounded to 17; 228 − 17 = 211). Total publications from table 1b, total residency publications (6.9 × 228 = 1,573.2, rounded to 1,573). Resident-years from not reported in the study; 228 residents x 5 years (standard length for urology) = 1,140. | Values |
| Waserman 2015 [194] | Residents assessed from table, page 493, 2008-2011 (19 men + 67 women = 86). Residents with at least 1 publication from table, page 493, 2008-2011, residents who published (2 men + 7 women = 9). Total publications from table, page 493, 2008-2011, publications (3 + 8 = 11). | No |
| Wieder 2022 [195]<sup>bc</sup> | Residents assessed from Abstract and Results, paragraph 1. Total publications from table 1, residency publications (1.3 × 758 = 985.4, rounded to 985). Resident-years from length not stated in the study; 758 x 3 years (standard length of ophthalmology residency after the intern year) = 2,274. | Values |
| Williams 2017 [196] | Residents assessed from Abstract and Results, paragraph 1. Residents with at least 1 publication and residents with a first-author publication from Results. Total publications from Results, paragraph 1. | No |
| Winn 2019 [197] | Residents assessed from table, non-recipients of the resident research grant (N = 229 (all tracks), the group with the reported publication outcome). Residents with a first-author publication from table, published manuscript. | Values |
| Wright-Chisem 2021 [198]<sup>bc</sup> | Residents assessed from Methods, Results, paragraph 1, table 1, and Abstract. Total publications from table 3 (22 × 22.0 + 59 × 16.5 = 1,457.5, rounded to 1,458). Resident-years from Methods (81 × 5 years = 405). | Values |
| Yang 2012 [199]<sup>bc</sup> | Residents assessed from Results, paragraph 1, and Abstract (459 men + 84 women = 543). Residents with at least 1 publication from Results, paragraph 2 (0.84 × 459 = 385.6 and 0.80 × 84 = 67.2; 386 + 67 = 453). Residents with a first-author publication from Results, paragraph 2 (0.70 × 459 = 321.3 and 0.65 × 84 = 54.6; 321 + 55 = 376). Total publications from table 1, mean publications during residency (459 men × 4.8 + 84 women × 3.0 = 2,455.2, rounded to 2,455). Resident-years from Methods (543 × 3 = 1,629). | No |
| Yang 2025 [200] | Residents assessed from Results and table 1. Residents with at least 1 publication from Abstract and Results. | Eligibility |
| Zhang 2025 [201]<sup>b</sup> | Residents assessed from tables 2 and 3. Total publications from table 3 (1.11 × 106 = 117.66, rounded to 118). Resident-years from table 2, training years (16 × 2 + 90 × 3 = 302). | No |

Numbers in square brackets are reference numbers in the reference list of the main text. Tables and figures of the study report are written in lower case. The extracted values are given in Table S3 and Supplementary Material 2.
<sup>a</sup>Eligibility: the 2 screening agents disagreed about eligibility; resolved against the source and confirmed by I.-C.T. Values: I.-C.T. decided a disagreement between the 2 extraction models (US studies) or between the 2 screening agents (other studies). Both: both kinds of disagreement. No: no disagreement.
<sup>b</sup>Approximate publication total derived from a reported mean or rate; a sensitivity analysis excludes all approximate totals.
<sup>c</sup>Resident-years calculated with the standard training length because the report did not state the length or counting period; excluded in a sensitivity analysis.
<sup>d</sup>No usable integer publication count was available; the fractional estimate is excluded from all Poisson rate analyses.

**Table S6.** Studies included with data provided by their corresponding authors. Reports that lacked the number of residents who published or the number assessed, included after their corresponding authors provided the missing numbers on request.

| First author, year | Ref. | Provided by | Data provided on request | Analyses |
|---|---|---|---|---|
| Lakhani, 2022 | 28 | Dhairya A. Lakhani | Publications for each resident in each academic year, anonymized | Partial-window sensitivity analysis; publications per resident-year |
| Gosnell, 2024 | 29 | Griffith G. Gosnell | Number of survey respondents and number with at least 1 publication | All-countries sensitivity analysis |
| Raymond, 2024 | 30 | Caitlin Raymond | Number of residents in each academic year (denominator) | Publications per resident-year |
| Fereydooni, 2024 | 31 | Arash Fereydooni | Number of residents, number with at least 1 publication, and total publications | Partial-window sensitivity analysis; publications per resident-year |

Ref., reference number in the reference list of the main text. Data were requested once from the corresponding author of each study that reported publication outcomes without the number of residents who published or the number assessed. None of these studies met the criteria for the US primary analysis.

**Table S7.**
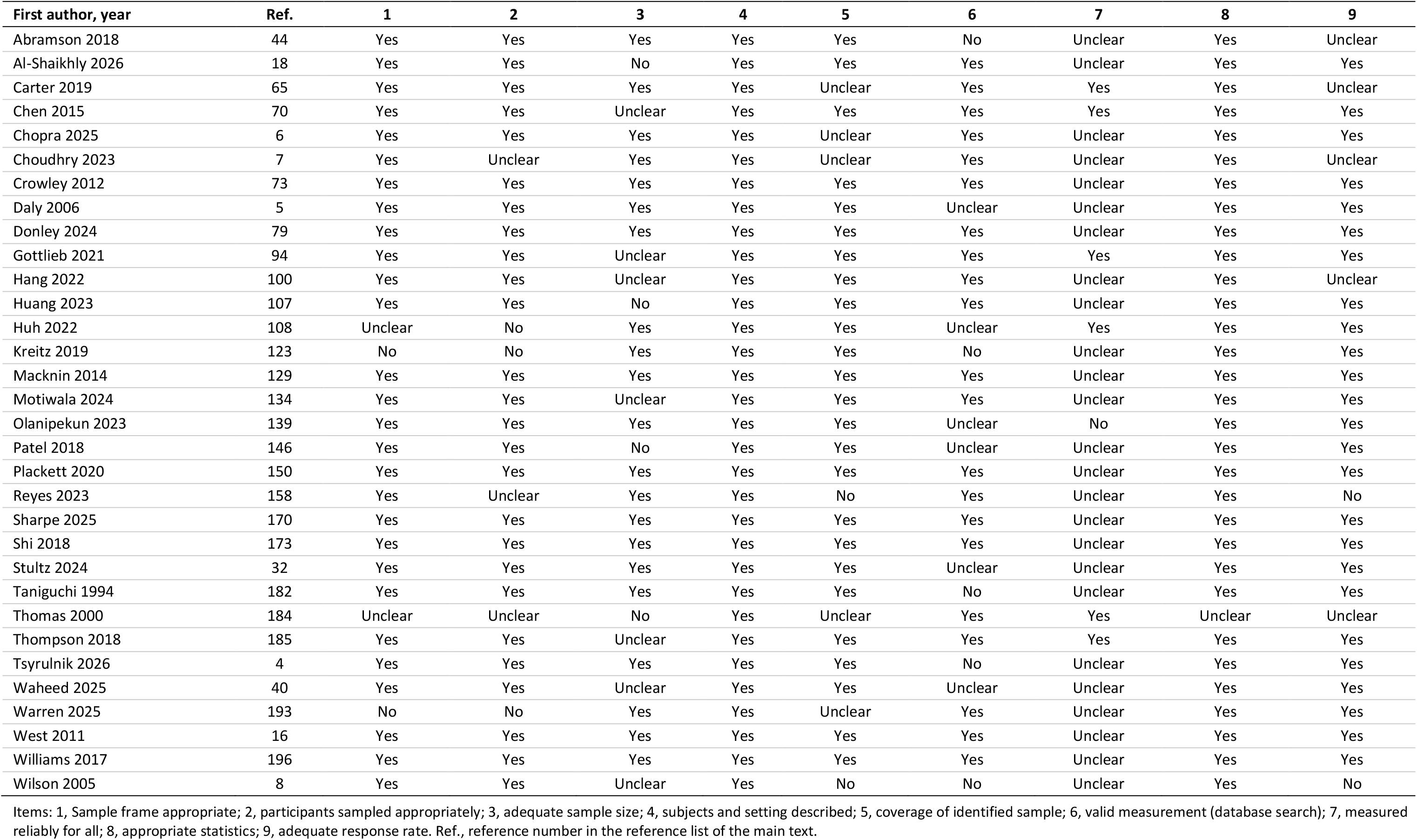
Risk of bias ratings for each study. The 32 US studies of the primary analysis, by item of the adapted Joanna Briggs Institute (JBI) checklist for prevalence studies.

| First author, year | Ref. | 1 | 2 | 3 | 4 | 5 | 6 | 7 | 8 | 9 |
|---|---|---|---|---|---|---|---|---|---|---|
| Abramson 2018 | 44 | Yes | Yes | Yes | Yes | Yes | No | Unclear | Yes | Unclear |
| Al-Shaikhly 2026 | 18 | Yes | Yes | No | Yes | Yes | Yes | Unclear | Yes | Yes |
| Carter 2019 | 65 | Yes | Yes | Yes | Yes | Unclear | Yes | Yes | Yes | Unclear |
| Chen 2015 | 70 | Yes | Yes | Unclear | Yes | Yes | Yes | Yes | Yes | Yes |
| Chopra 2025 | 6 | Yes | Yes | Yes | Yes | Unclear | Yes | Unclear | Yes | Yes |
| Choudhry 2023 | 7 | Yes | Unclear | Yes | Yes | Unclear | Yes | Unclear | Yes | Unclear |
| Crowley 2012 | 73 | Yes | Yes | Yes | Yes | Yes | Yes | Unclear | Yes | Yes |
| Daly 2006 | 5 | Yes | Yes | Yes | Yes | Yes | Unclear | Unclear | Yes | Yes |
| Donley 2024 | 79 | Yes | Yes | Yes | Yes | Yes | Yes | Unclear | Yes | Yes |
| Gottlieb 2021 | 94 | Yes | Yes | Unclear | Yes | Yes | Yes | Yes | Yes | Yes |
| Hang 2022 | 100 | Yes | Yes | Unclear | Yes | Yes | Yes | Unclear | Yes | Unclear |
| Huang 2023 | 107 | Yes | Yes | No | Yes | Yes | Yes | Unclear | Yes | Yes |
| Huh 2022 | 108 | Unclear | No | Yes | Yes | Yes | Unclear | Yes | Yes | Yes |
| Kreitz 2019 | 123 | No | No | Yes | Yes | Yes | No | Unclear | Yes | Yes |
| Macknin 2014 | 129 | Yes | Yes | Yes | Yes | Yes | Yes | Unclear | Yes | Yes |
| Motiwala 2024 | 134 | Yes | Yes | Unclear | Yes | Yes | Yes | Unclear | Yes | Yes |
| Olanipekun 2023 | 139 | Yes | Yes | Yes | Yes | Yes | Unclear | No | Yes | Yes |
| Patel 2018 | 146 | Yes | Yes | No | Yes | Yes | Unclear | Unclear | Yes | Yes |
| Plackett 2020 | 150 | Yes | Yes | Yes | Yes | Yes | Yes | Unclear | Yes | Yes |
| Reyes 2023 | 158 | Yes | Unclear | Yes | Yes | No | Yes | Unclear | Yes | No |
| Sharpe 2025 | 170 | Yes | Yes | Yes | Yes | Yes | Yes | Unclear | Yes | Yes |
| Shi 2018 | 173 | Yes | Yes | Yes | Yes | Yes | Yes | Unclear | Yes | Yes |
| Stultz 2024 | 32 | Yes | Yes | Yes | Yes | Yes | Unclear | Unclear | Yes | Yes |
| Taniguchi 1994 | 182 | Yes | Yes | Yes | Yes | Yes | No | Unclear | Yes | Yes |
| Thomas 2000 | 184 | Unclear | Unclear | No | Yes | Unclear | Yes | Yes | Unclear | Unclear |
| Thompson 2018 | 185 | Yes | Yes | Unclear | Yes | Yes | Yes | Yes | Yes | Yes |
| Tsyrulnik 2026 | 4 | Yes | Yes | Yes | Yes | Yes | No | Unclear | Yes | Yes |
| Waheed 2025 | 40 | Yes | Yes | Unclear | Yes | Yes | Unclear | Unclear | Yes | Yes |
| Warren 2025 | 193 | No | No | Yes | Yes | Unclear | Yes | Unclear | Yes | Yes |
| West 2011 | 16 | Yes | Yes | Yes | Yes | Yes | Yes | Unclear | Yes | Yes |
| Williams 2017 | 196 | Yes | Yes | Yes | Yes | Yes | Yes | Unclear | Yes | Yes |
| Wilson 2005 | 8 | Yes | Yes | Unclear | Yes | No | No | Unclear | Yes | No |

Items: 1, Sample frame appropriate; 2, participants sampled appropriately; 3, adequate sample size; 4, subjects and setting described; 5, coverage of identified sample; 6, valid measurement (database search); 7, measured reliably for all; 8, appropriate statistics; 9, adequate response rate. Ref., reference number in the reference list of the main text.

**Table S8.** Pooled proportions by standard training length, with a linear model. Proportions of residents with at least 1 peer-reviewed publication in 26 US studies without pre-intervention cohorts.

| Standard length | Specialties, n of studies | Studies, n | Residents, n | Pooled proportion, % (95% CI) |
|---|---|---|---|---|
| 3 years | Internal medicine 4; emergency medicine 1; pediatrics 1 | 6 | 1,517 | 45.0 (25.8–65.7) |
| 4 years | Ophthalmology 4; anesthesiology 1; dermatology 1; physical medicine and rehabilitation 1 | 7 | 3,331 | 58.5 (42.5–72.8) |
| 5 years | Orthopaedic surgery 5; otolaryngology 2; radiation oncology 2; urology 2; general surgery 1 | 12 | 7,608 | 77.8 (63.9–87.4) |
| 7 years | Neurological surgery 1 | 1 | 717 | 69.5<sup>a</sup> |
| **Meta-regression models by training length** |  |  |  |  |
| **Model** | **Term** | **OR (95% CI)** | ***p*** | **Residual τ<sup>2</sup>** |
| Linear | Per additional year of standard training | 1.68 (1.10–2.57) | 0.02 | 1.06 |
| Length and specialty | Per additional year | 1.16 (0.69–1.97) | 0.58 | NA |
|  | Medical vs. all other specialties | 0.29 (0.09–0.97) | 0.045 | NA |

Mixed-effects logistic meta-regression; OR, odds ratio between study cohorts; CI, confidence interval; τ<sup>2</sup>, residual between-study variance on the logit scale. Training length is fixed by specialty, so length and specialty cannot be separated in these data. Standard length counts from the first postgraduate year and includes any required preliminary year (4 years for ophthalmology); the resident-years in Table S5 instead use the residency years each study counted or, when assumed, the length of the residency after the intern year (3 years for ophthalmology). NA, not applicable: residual τ<sup>2</sup> was extracted for the single-term models only.
Same 4-year length: ophthalmology 60.8% (4 studies; 95% CI, 53.2–67.9) vs. physical medicine and rehabilitation 19.1% (1 study).
<sup>a</sup>Single study: observed proportion.

**Table S9.**
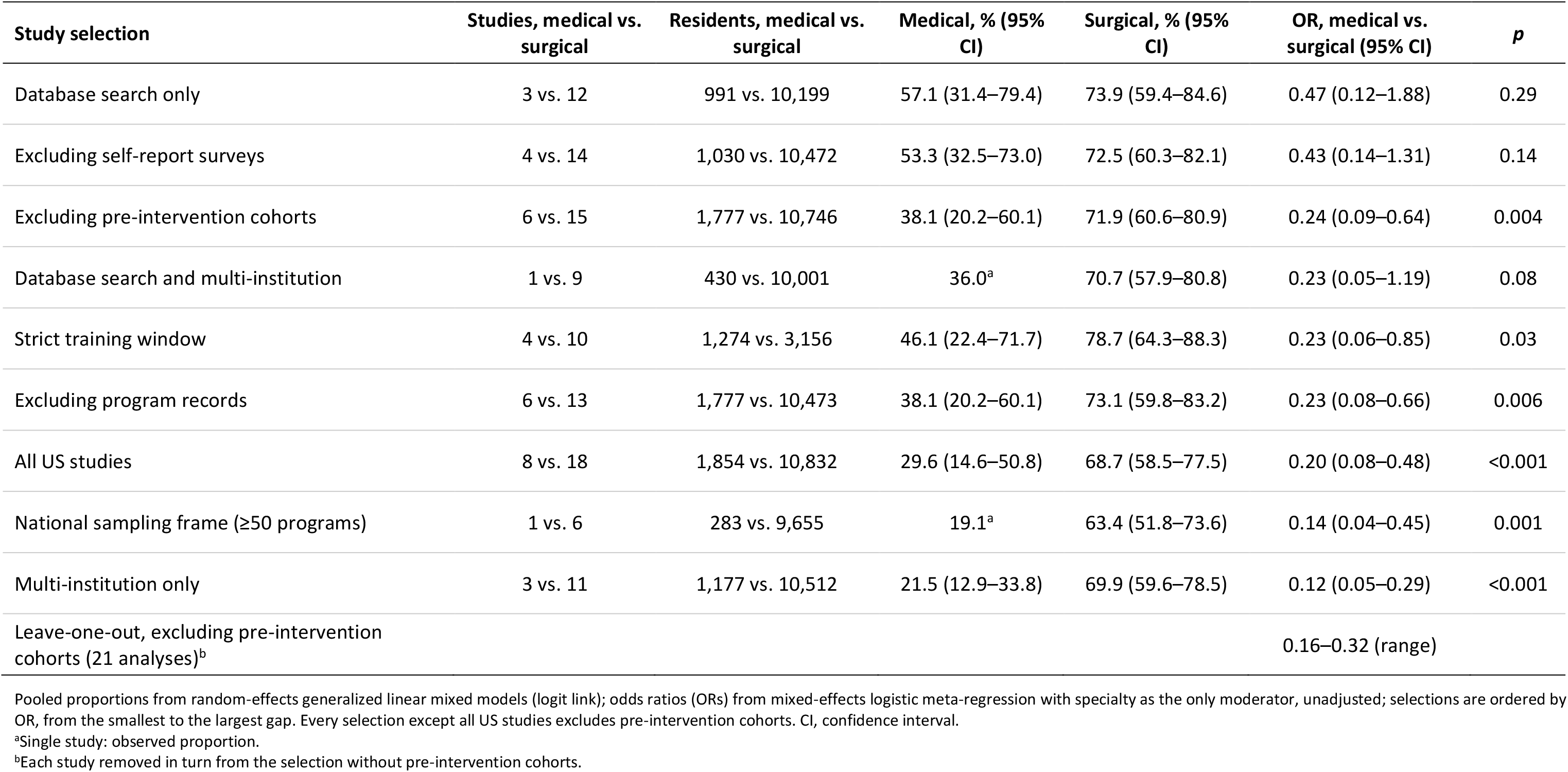
Medical vs. surgical gap under study selections and leave-one-out analysis. Pooled proportions in medical and surgical specialties and the medical vs. surgical odds ratio under 9 nested study selections of US studies.

**Table S10.** Distribution of published US cohort estimates by specialty group. Percentiles of the predictive distribution for proportions and rates, excluding pre-intervention cohorts.

| **Specialty group** | **Studies, n** | **10th** | **25th** | **50th** | **75th** | **90th** |
|---|---|---|---|---|---|---|
| **Residents with at least 1 publication, %** |  |  |  |  |  |  |
| Surgical specialties | 15 | 40.1 | 56.2 | 71.9 | 83.6 | 90.7 |
| Other specialties<sup>a</sup> | 6 | 31.3 | 49.8 | 67.1 | 80.8 | 90.1 |
| All specialties | 27 | 27.6 | 44.5 | 64.2 | 80.1 | 89.4 |
| Medical specialties | 6 | 9.1 | 20.4 | 38.1 | 59.7 | 79.1 |
| **Publications per resident-year** |  |  |  |  |  |  |
| Surgical specialties | 29 | 0.39 | 0.60 | 0.98 | 1.59 | 2.49 |
| All specialties | 42 | 0.28 | 0.46 | 0.79 | 1.37 | 2.25 |
| Other specialties<sup>a</sup> | 10 | 0.20 | 0.34 | 0.59 | 1.01 | 1.72 |
| Medical specialties | 3 | 0.02 | 0.12 | 0.27 | 0.62 | 3.56 |

Percentiles of the predictive distribution from random-effects generalized linear mixed models, using a t distribution with k − 2 degrees of freedom (k, number of studies; the prediction intervals in the main text use the package default of k − 1 degrees of freedom). Pre-intervention cohorts are excluded from every row. The percentiles describe the spread of published study cohorts, many of which pooled several programs. Percentiles were estimated for groups with at least 3 studies.
<sup>a</sup>Radiology, radiation oncology, pathology, anesthesiology, emergency medicine, and dermatology; in the rate analysis, anesthesiology, dermatology, diagnostic radiology, emergency medicine, and radiation oncology.

**Table S11.**
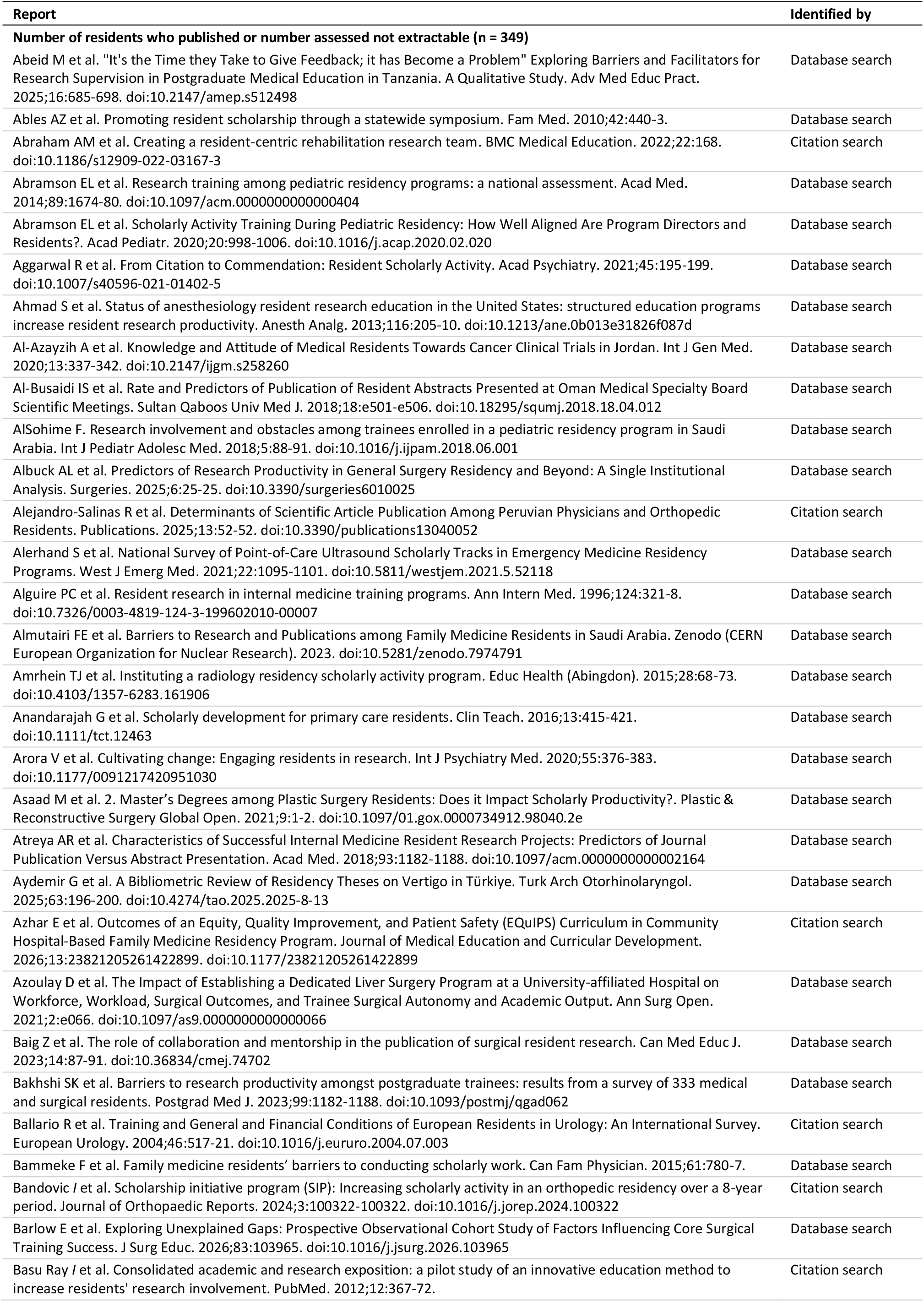
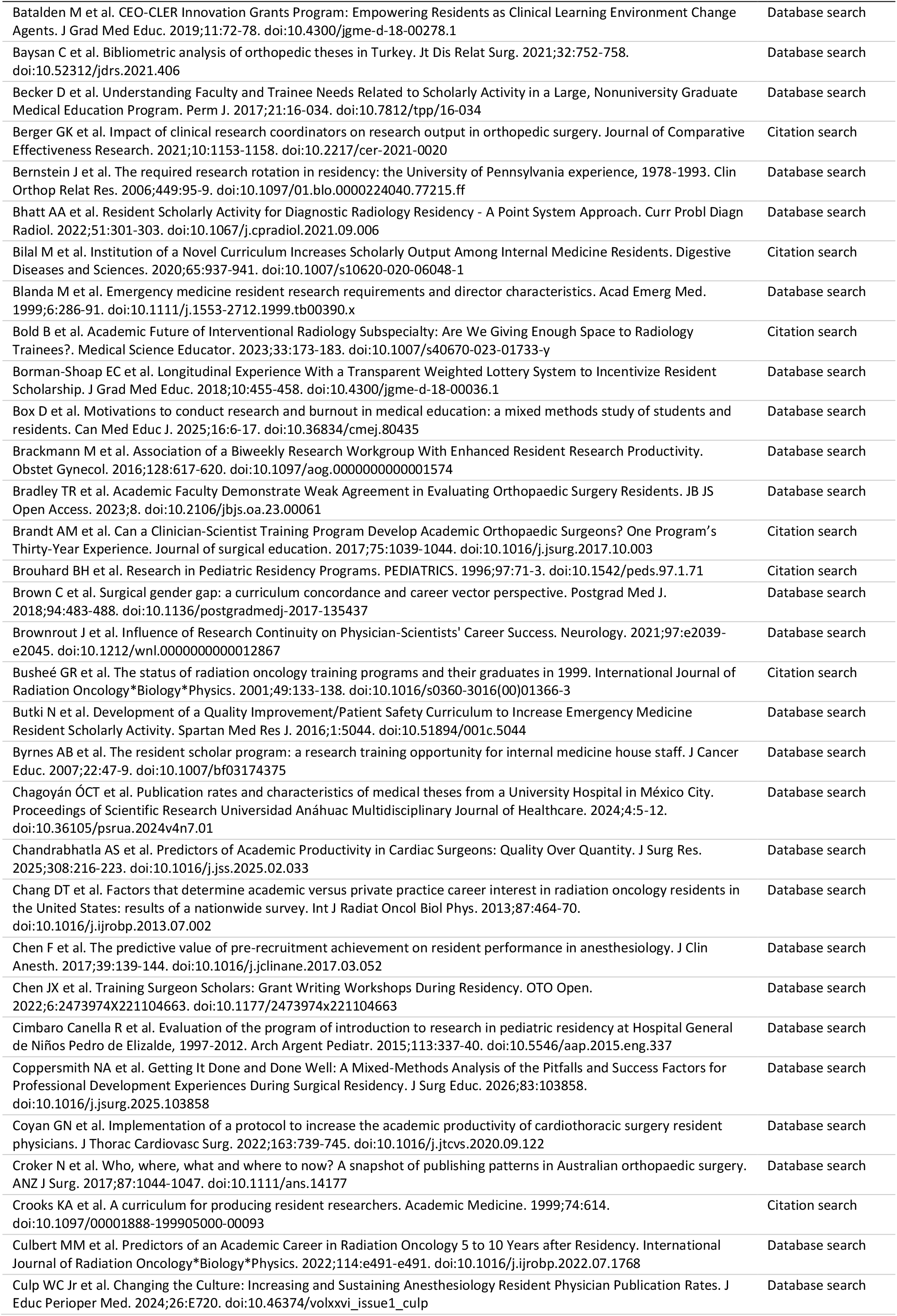
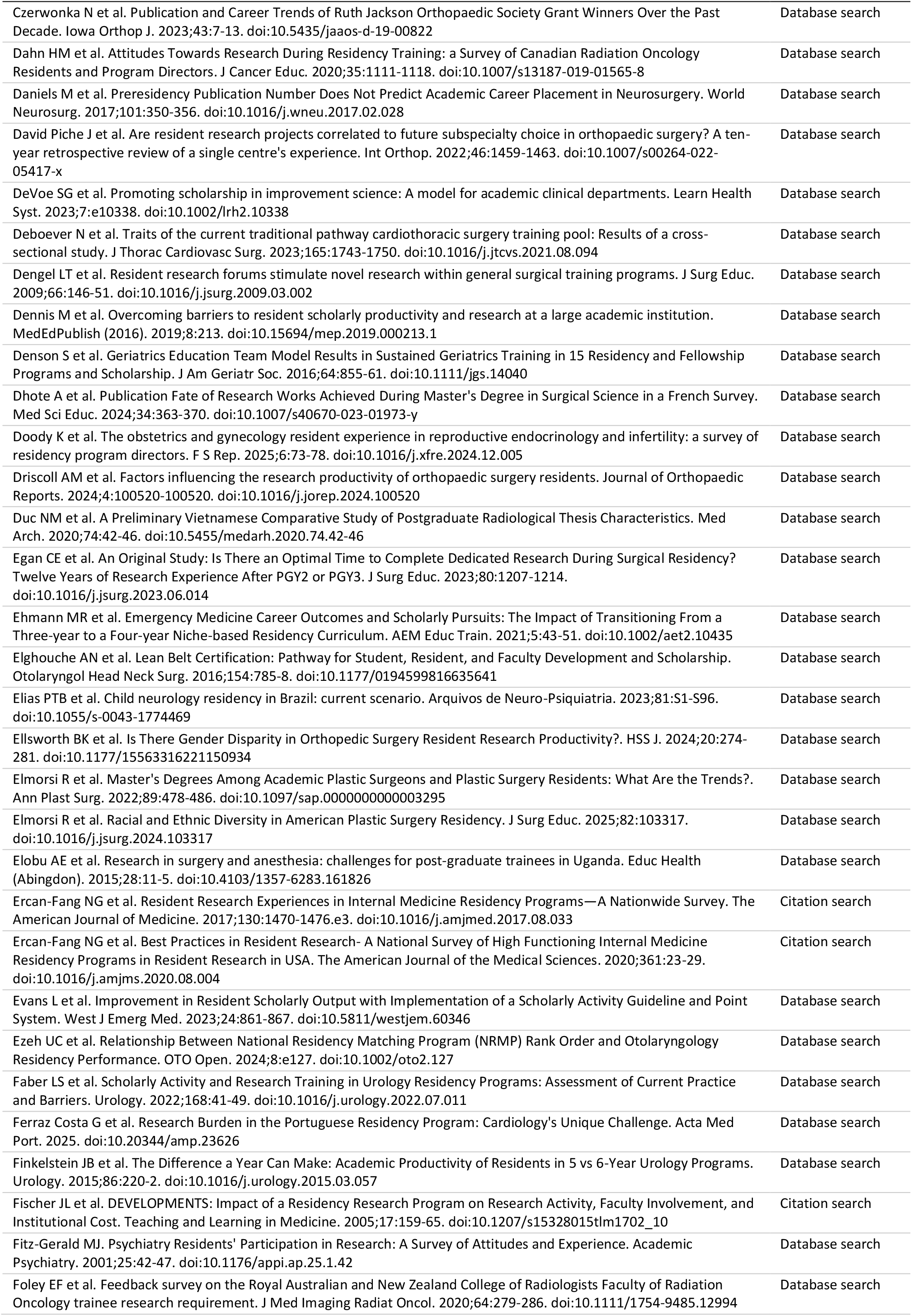
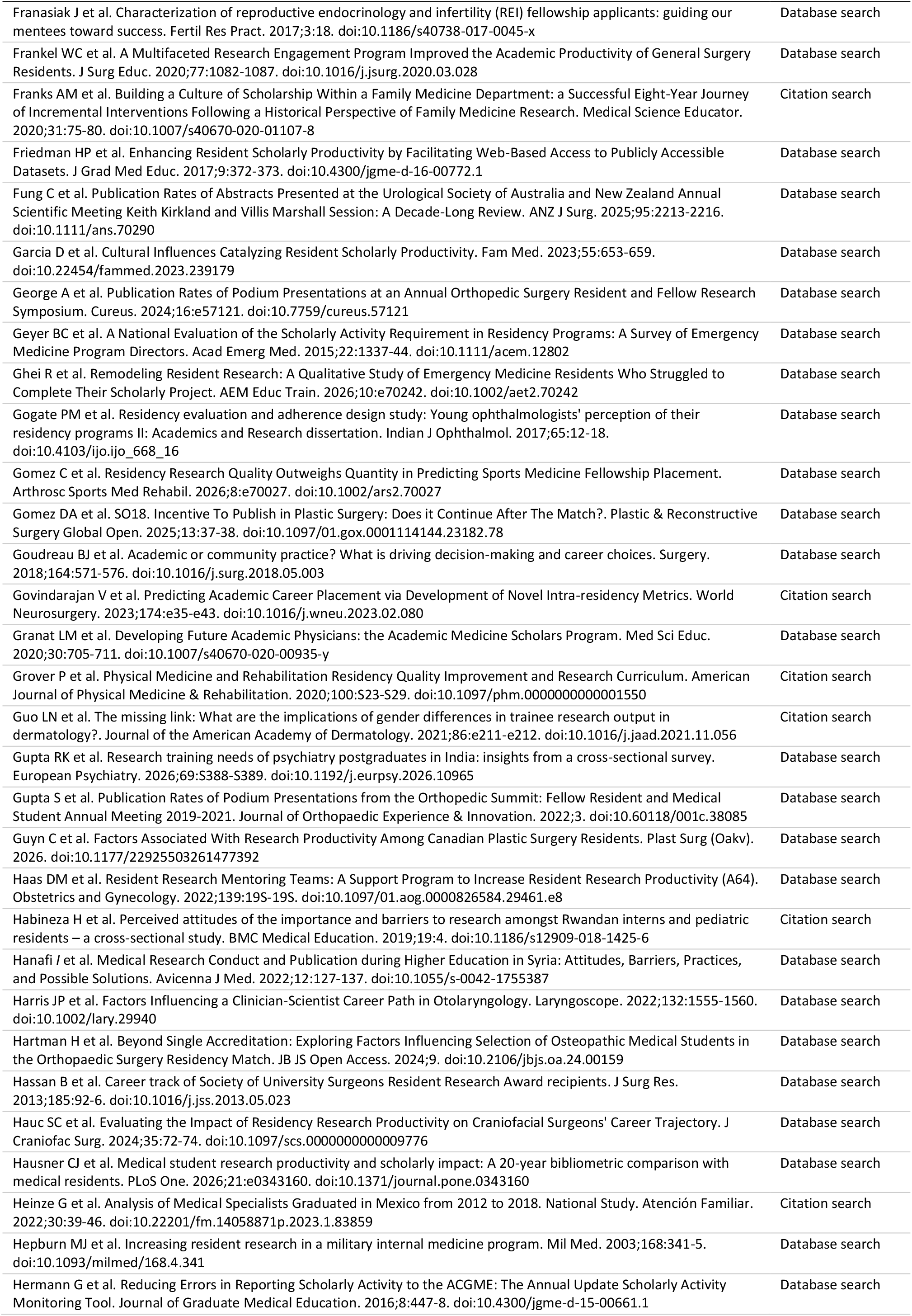
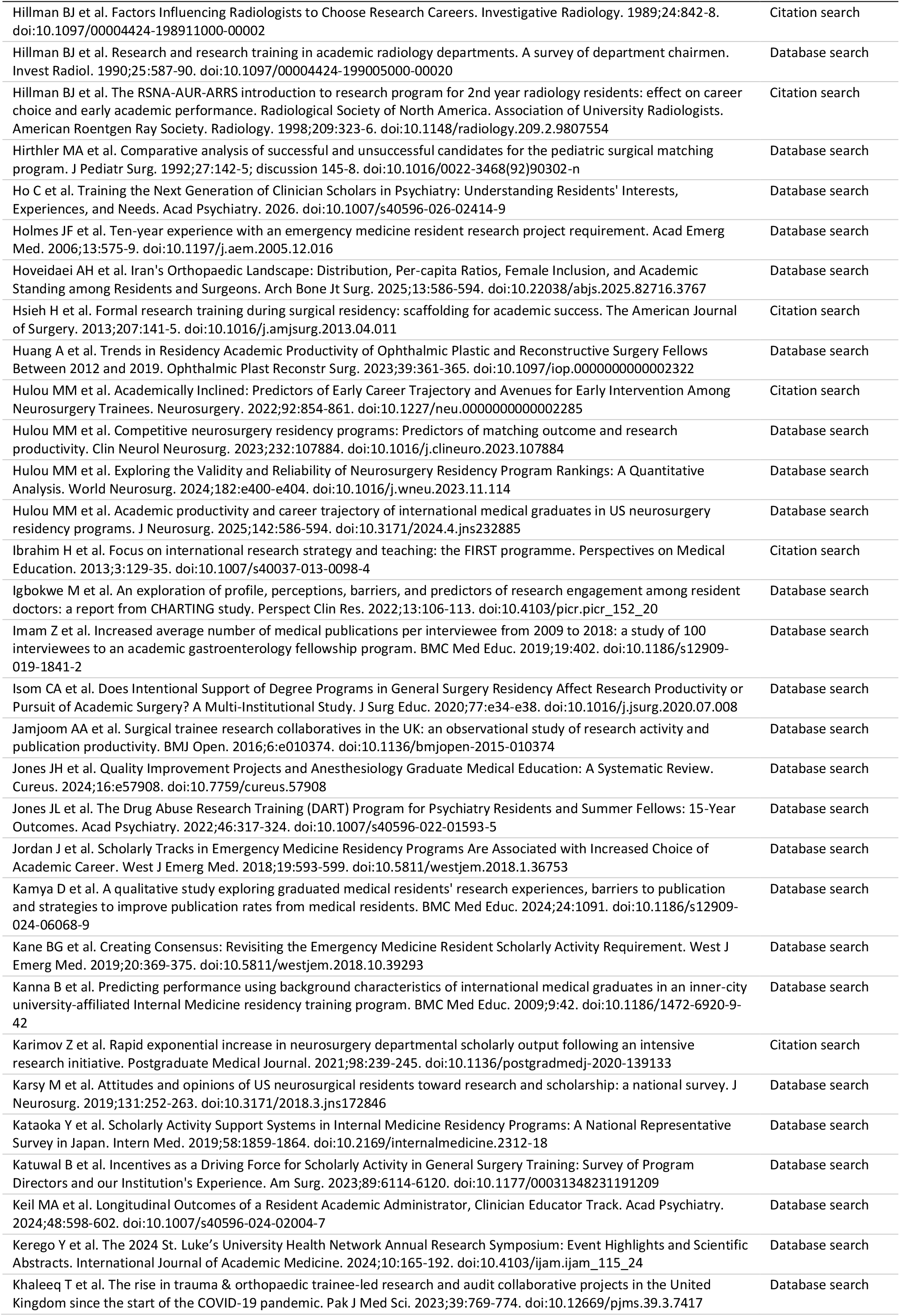
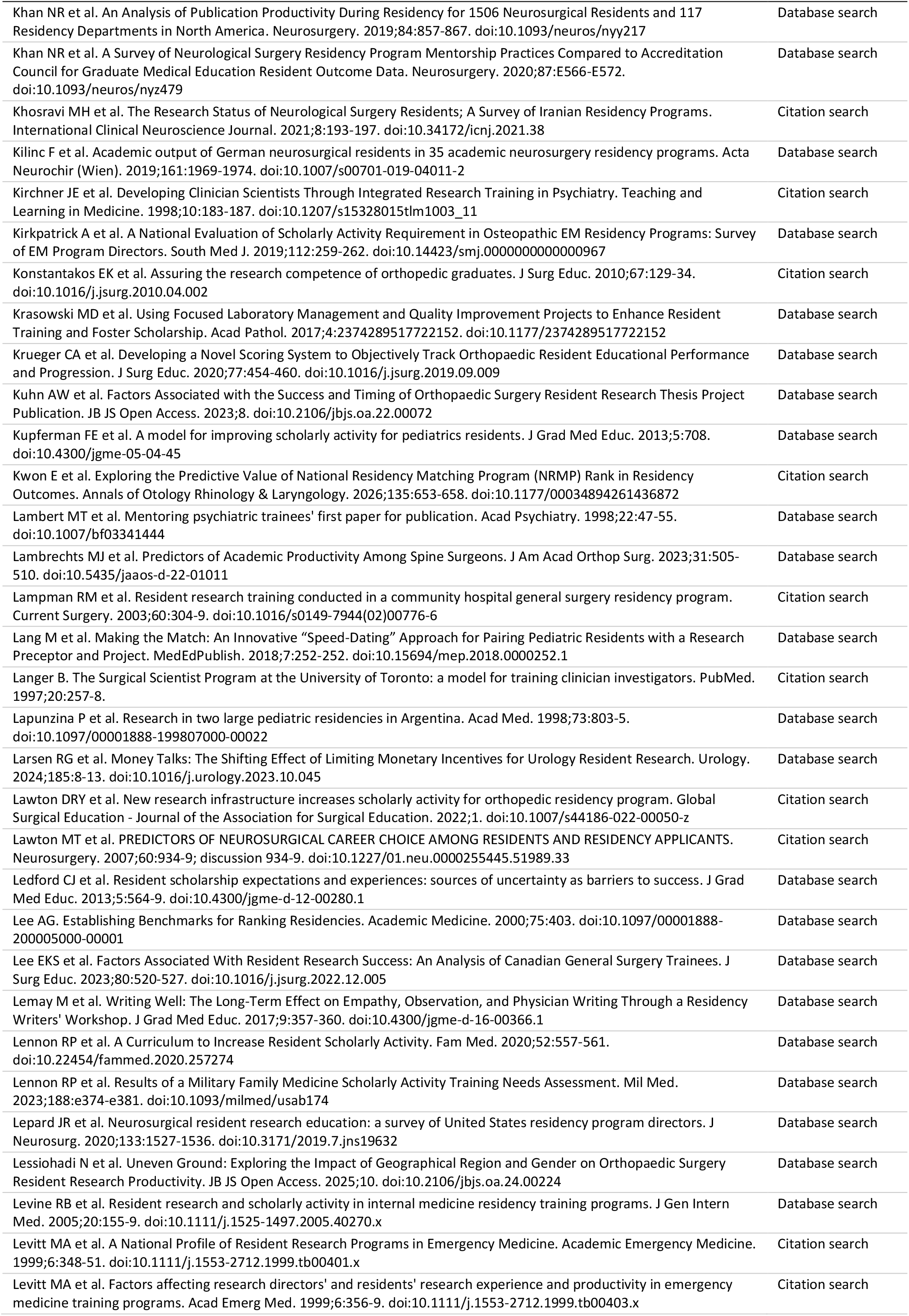
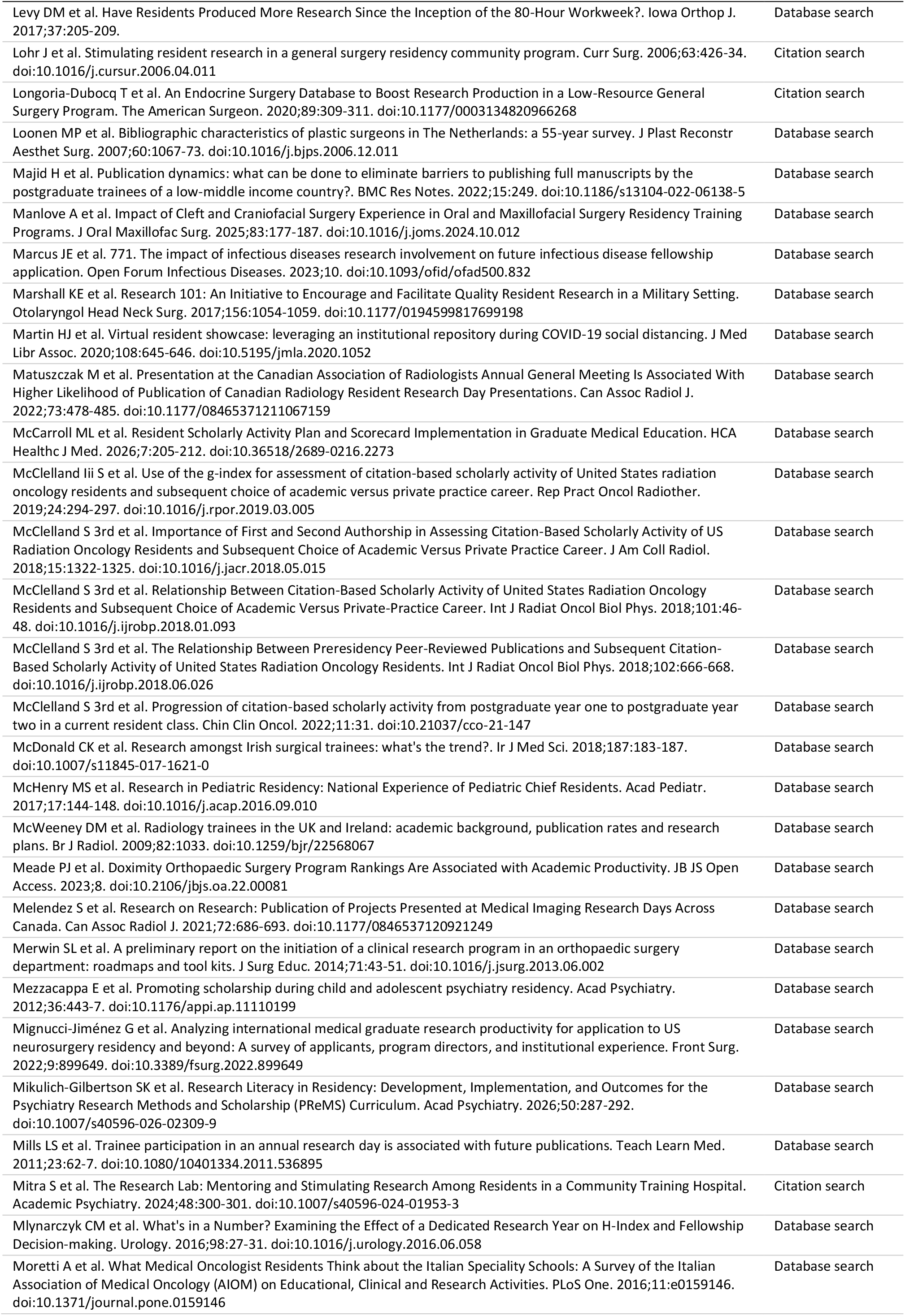
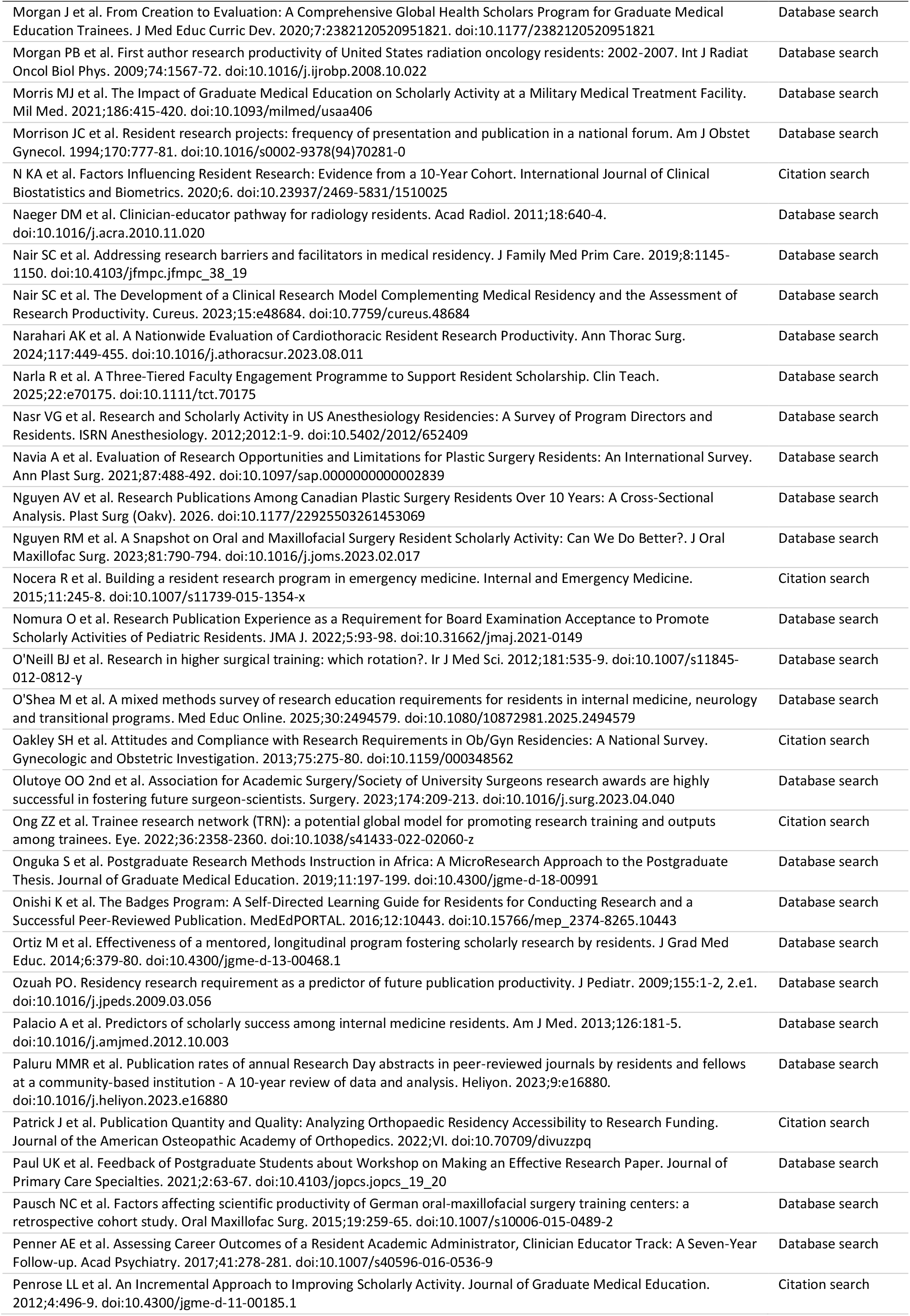
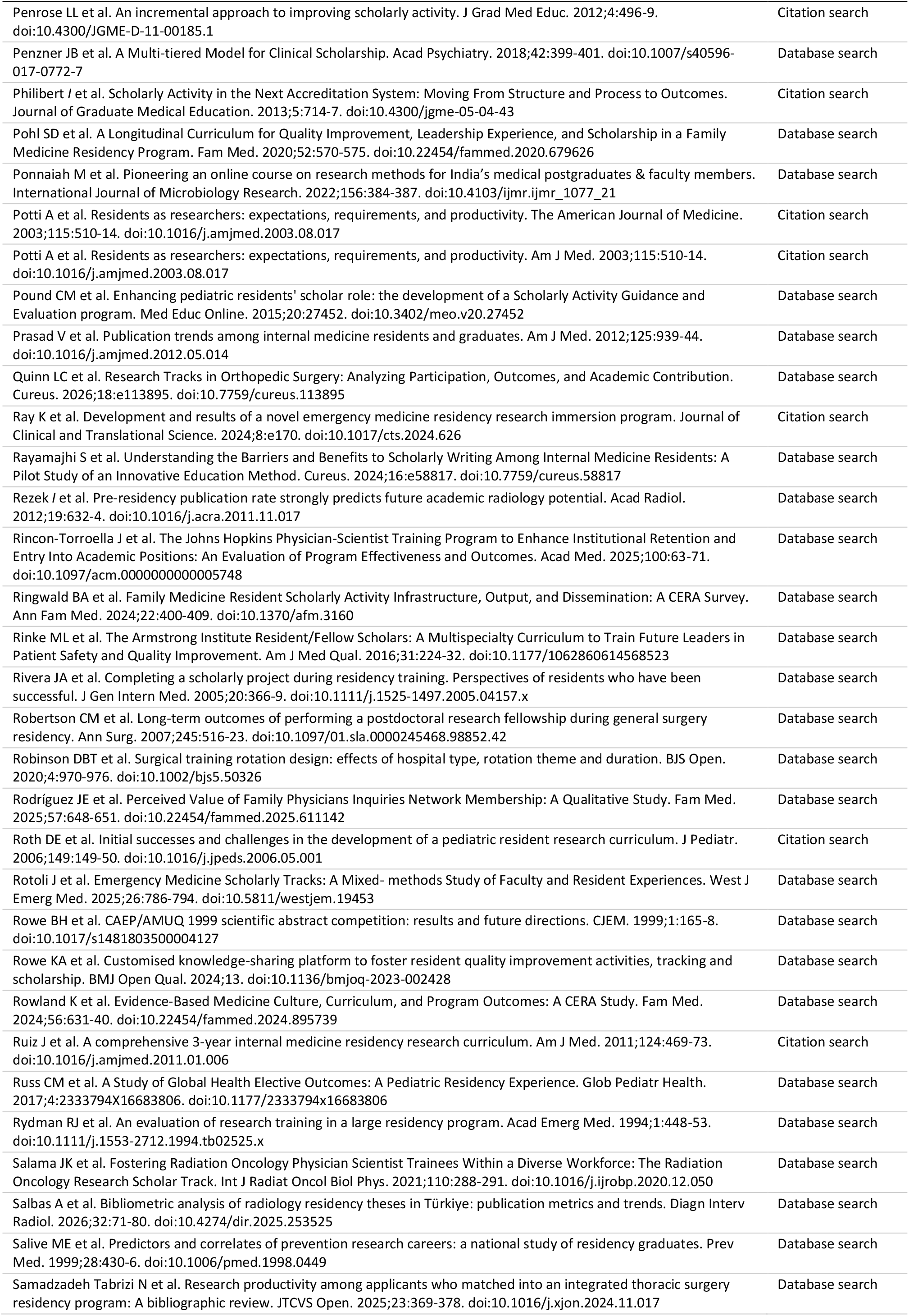
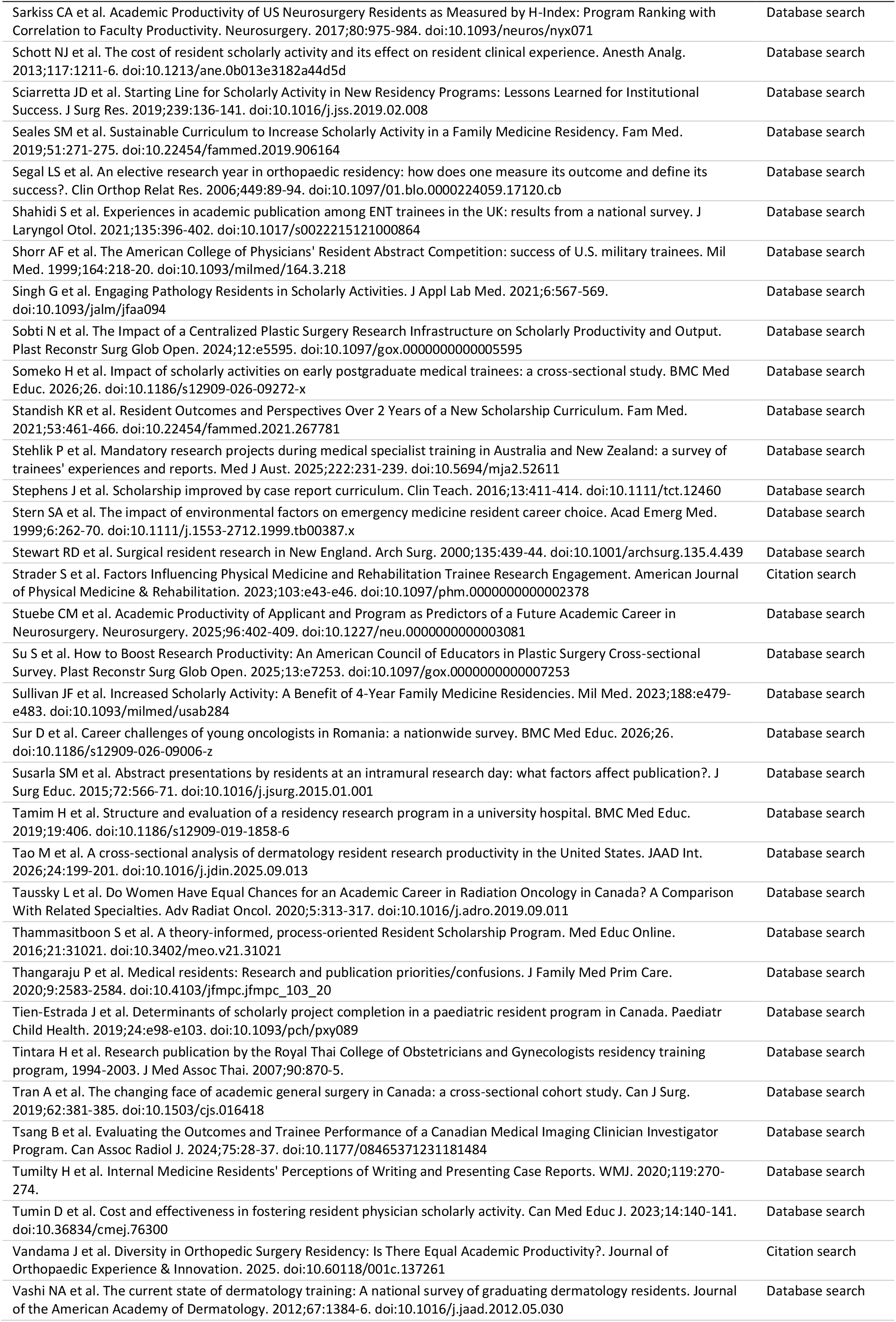
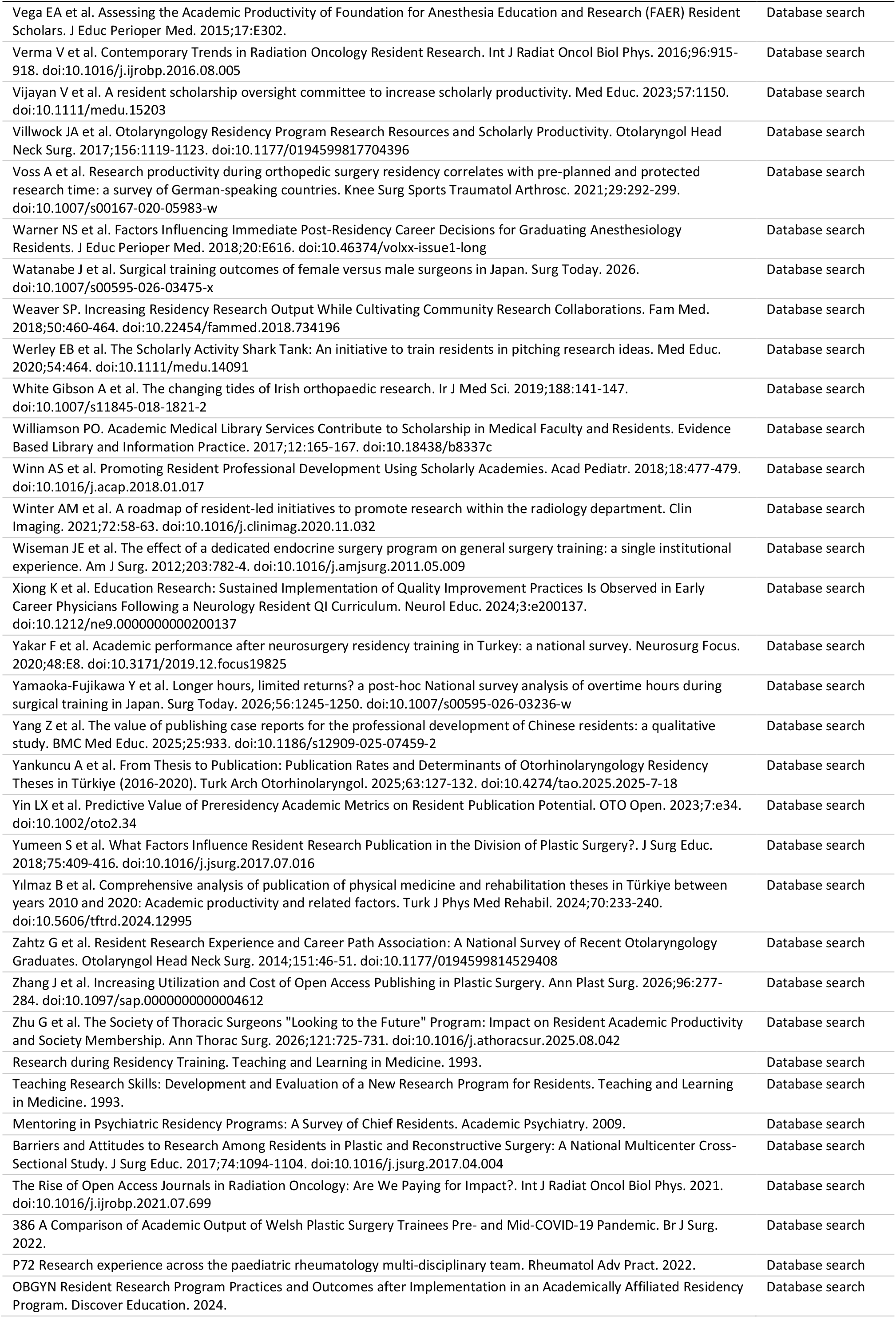
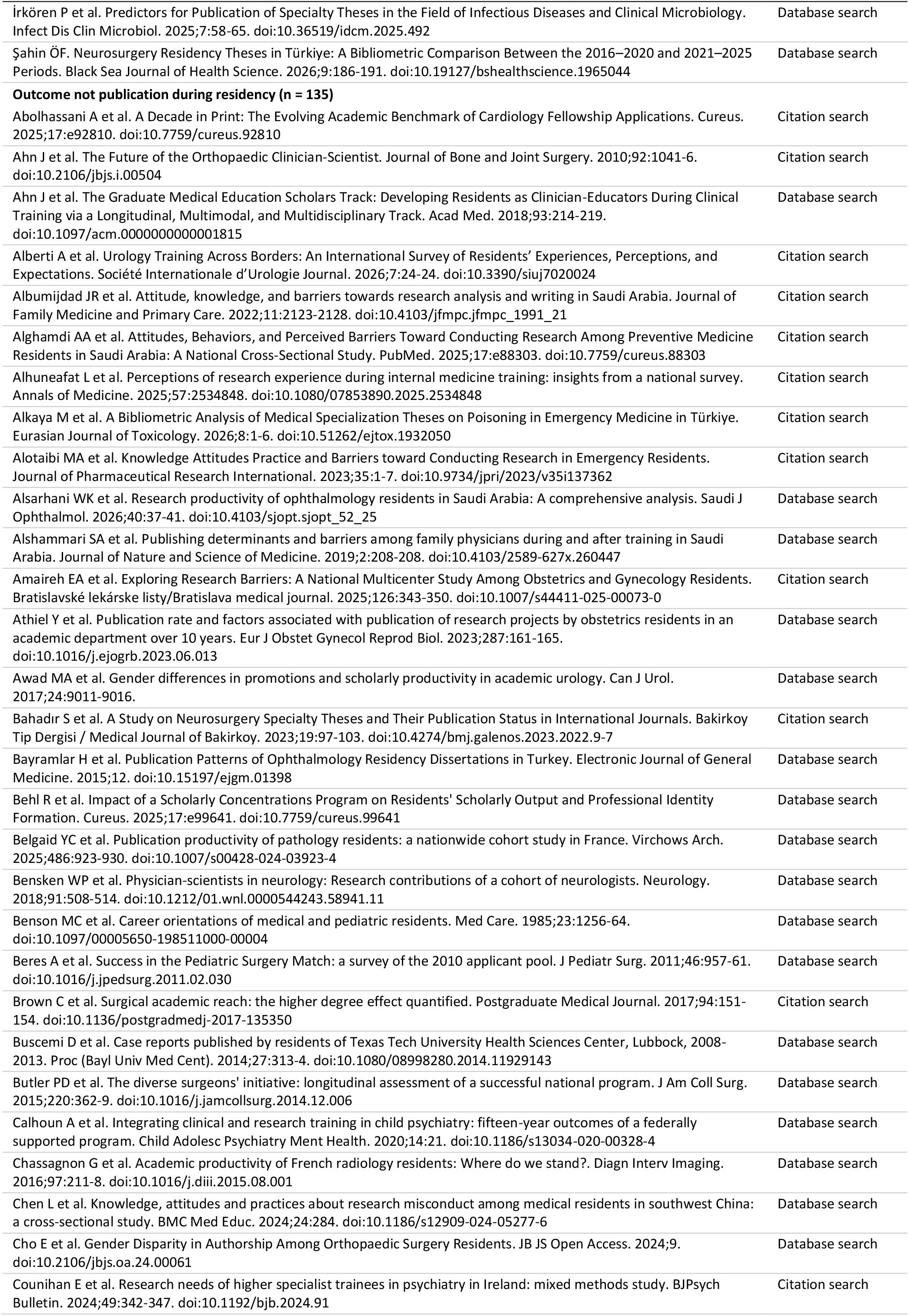
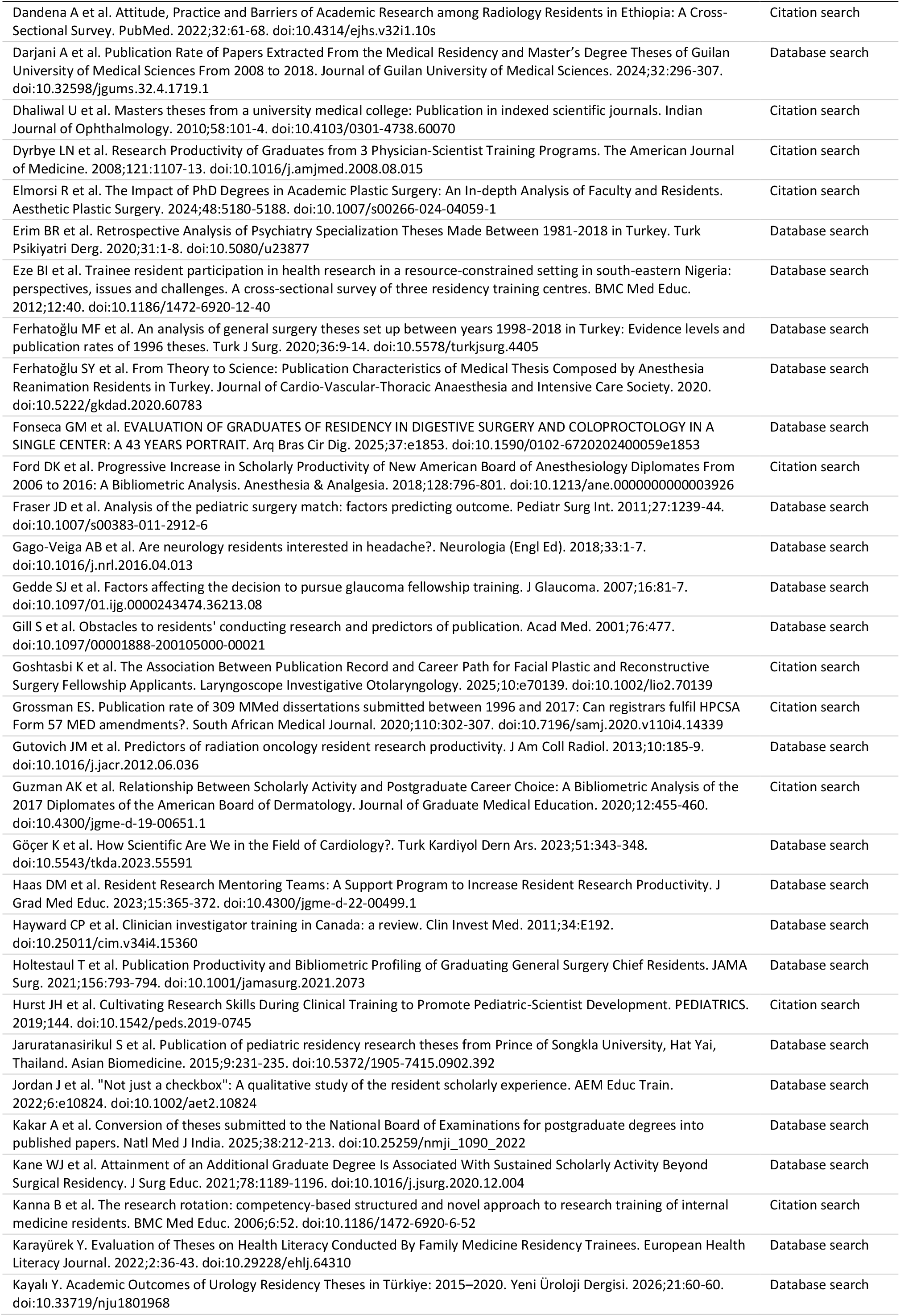
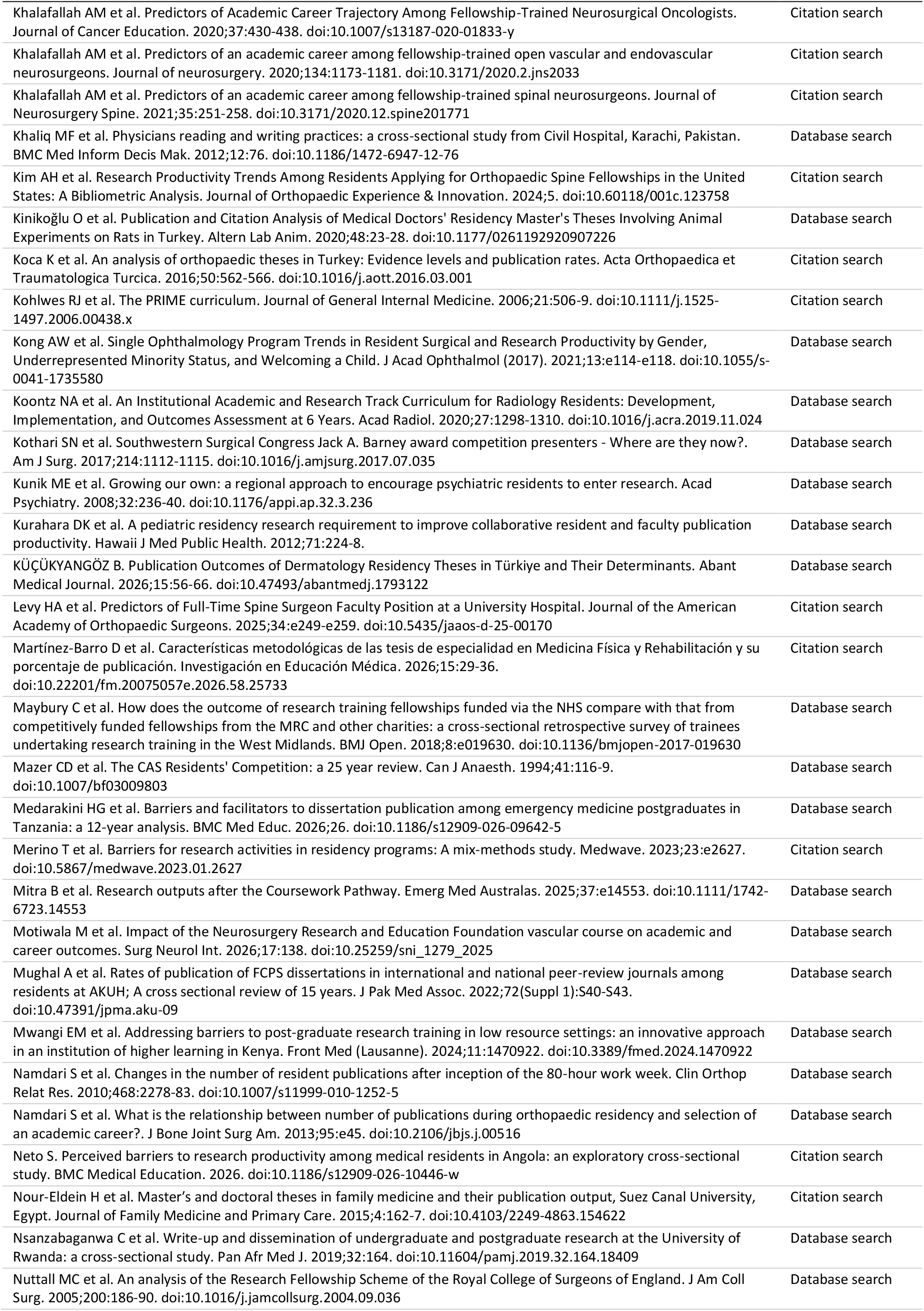
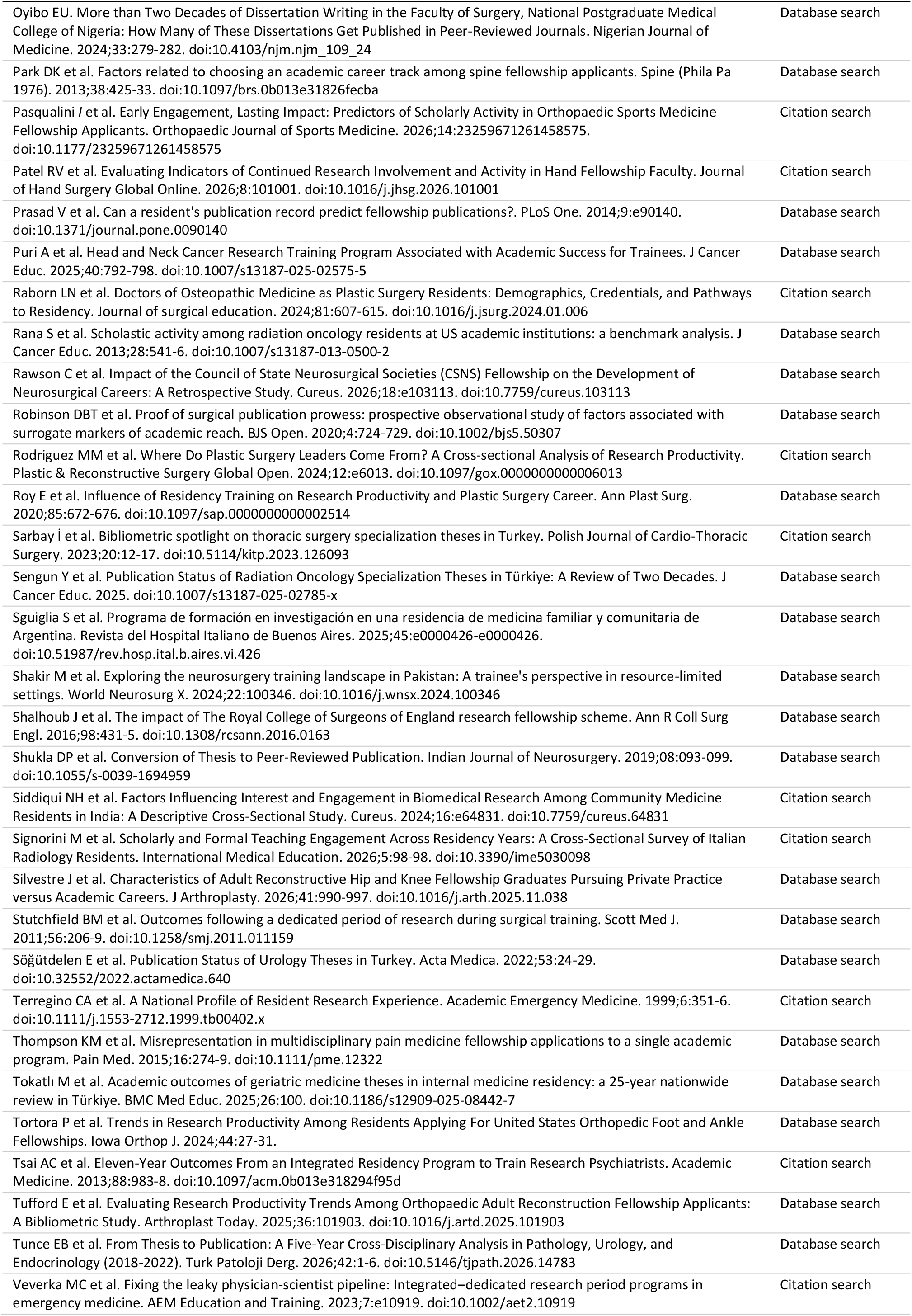
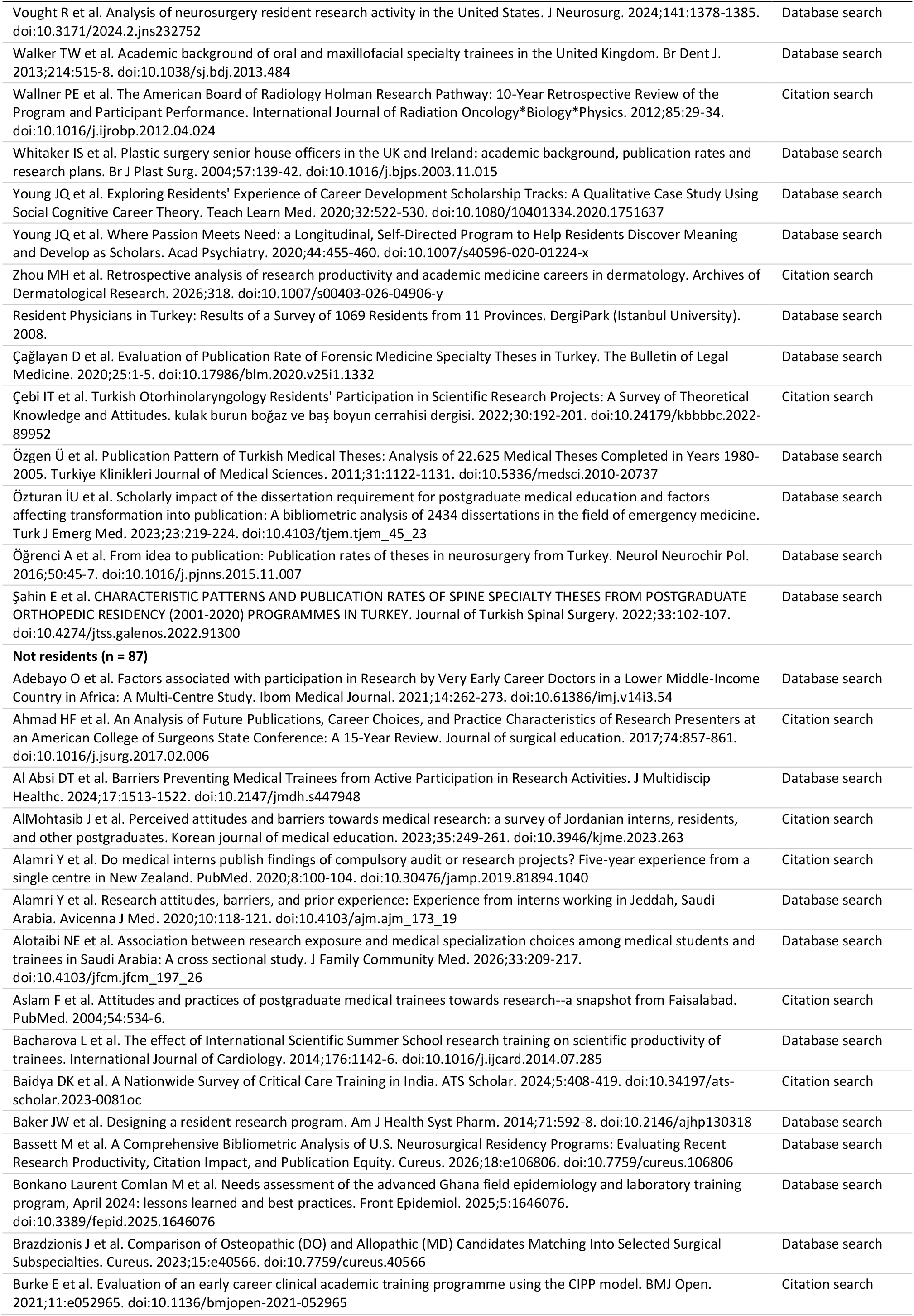
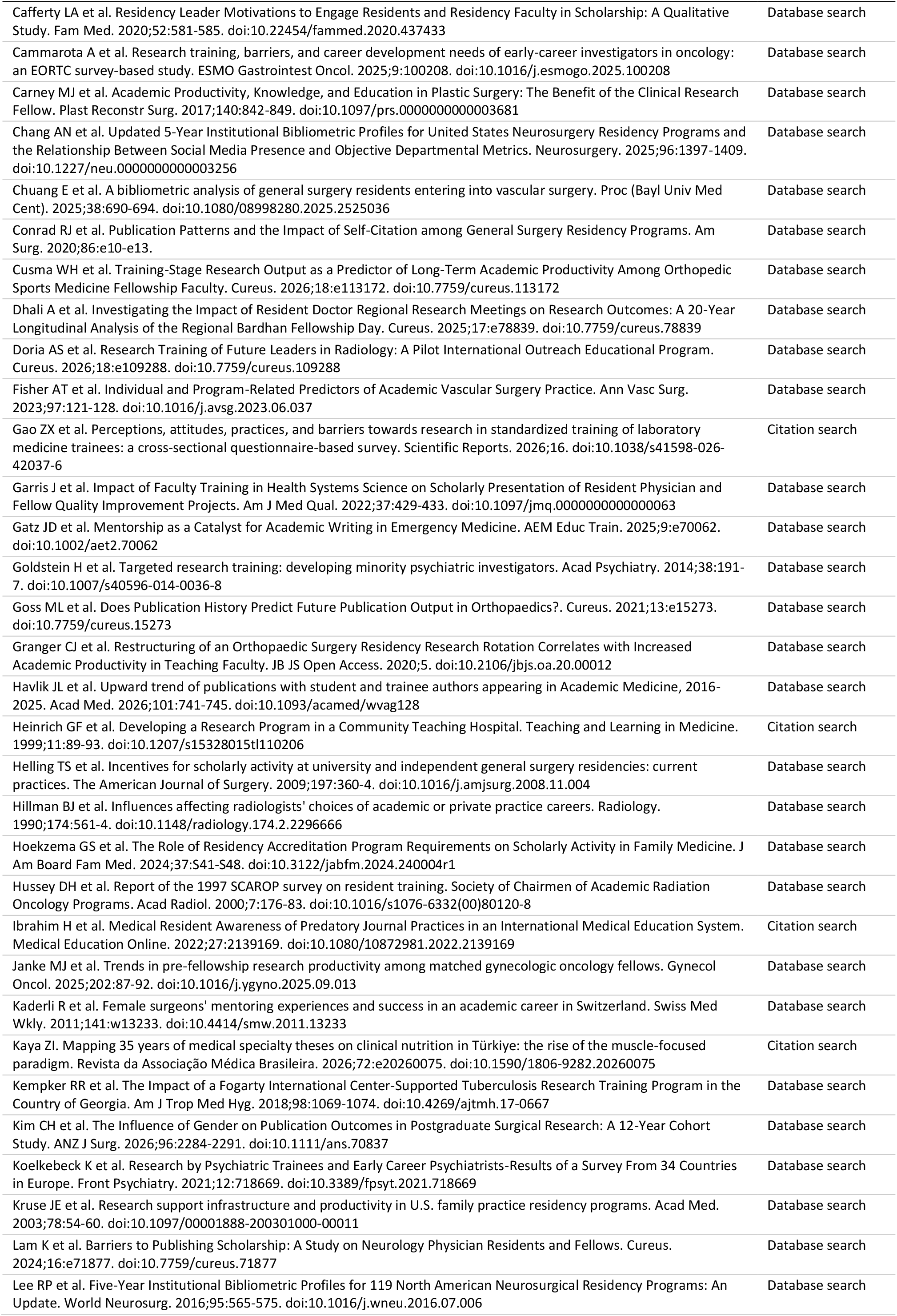
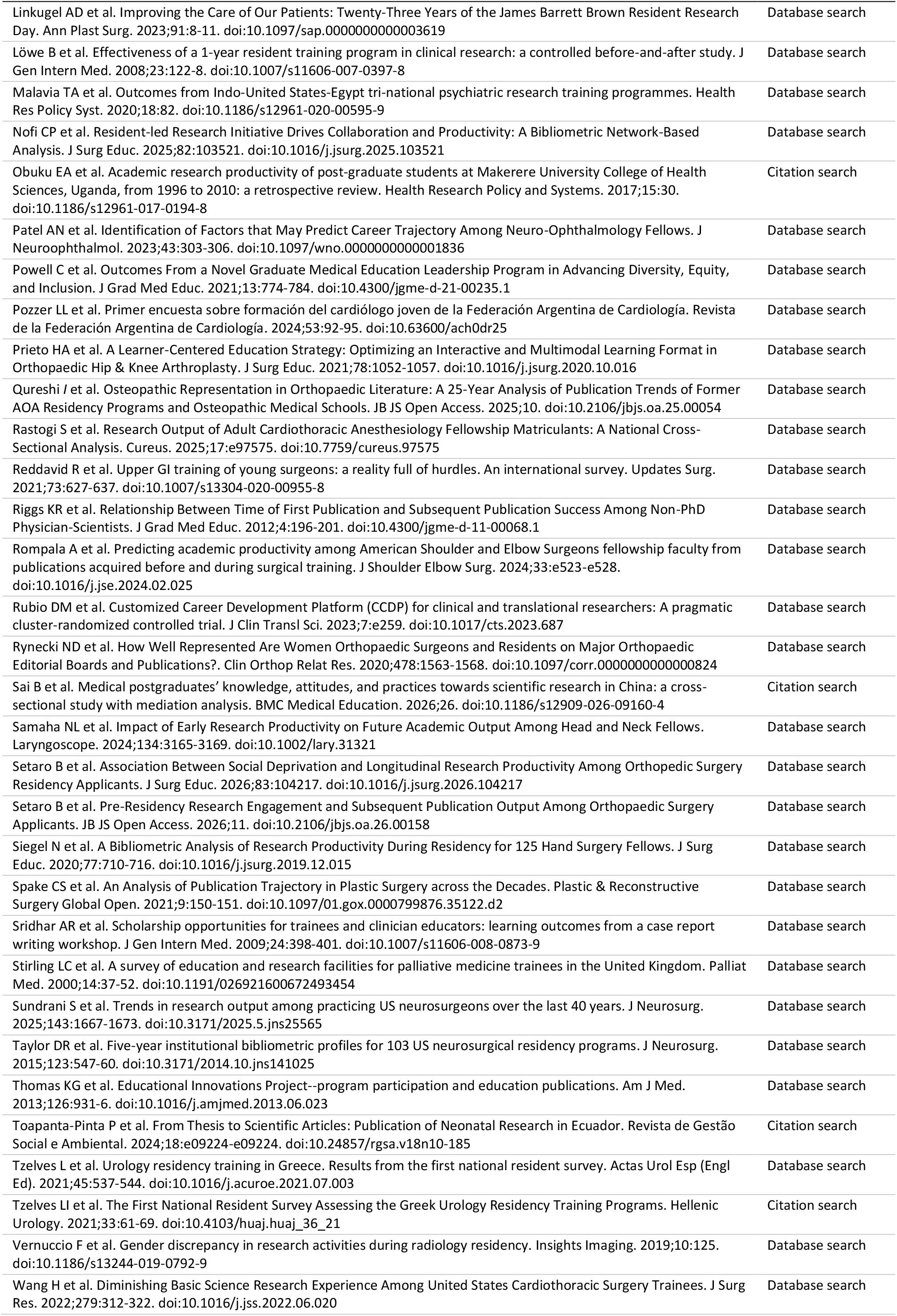
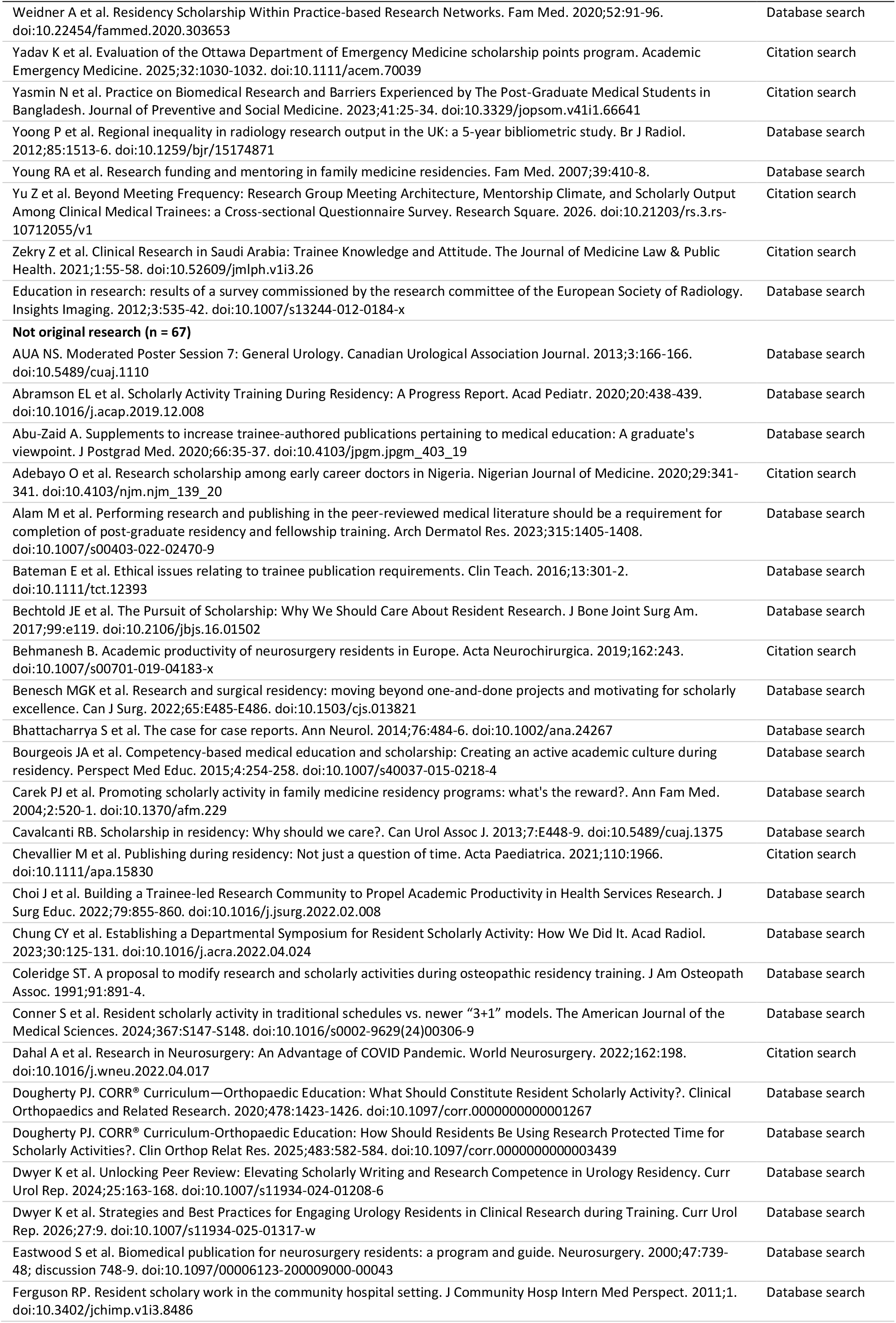
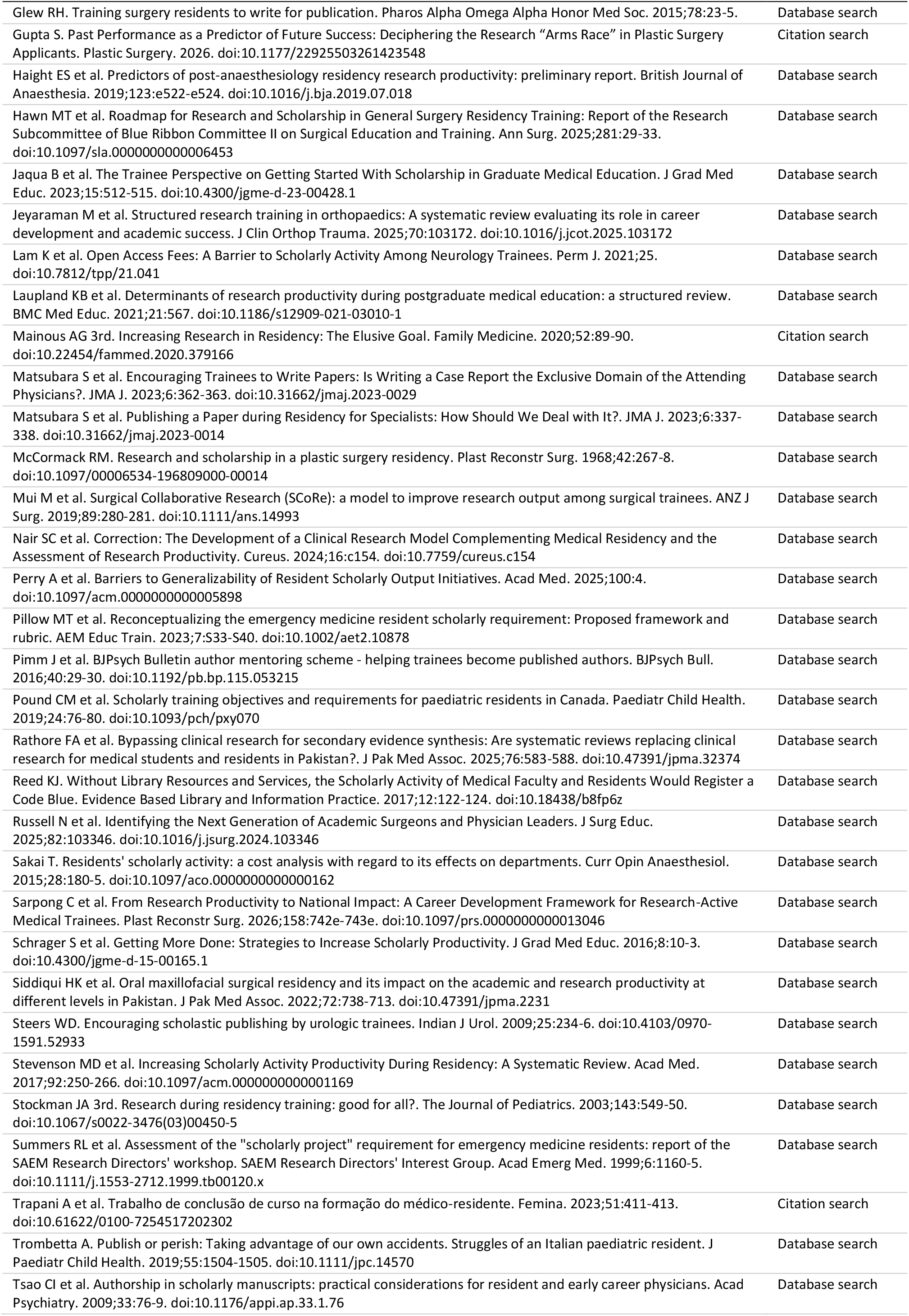
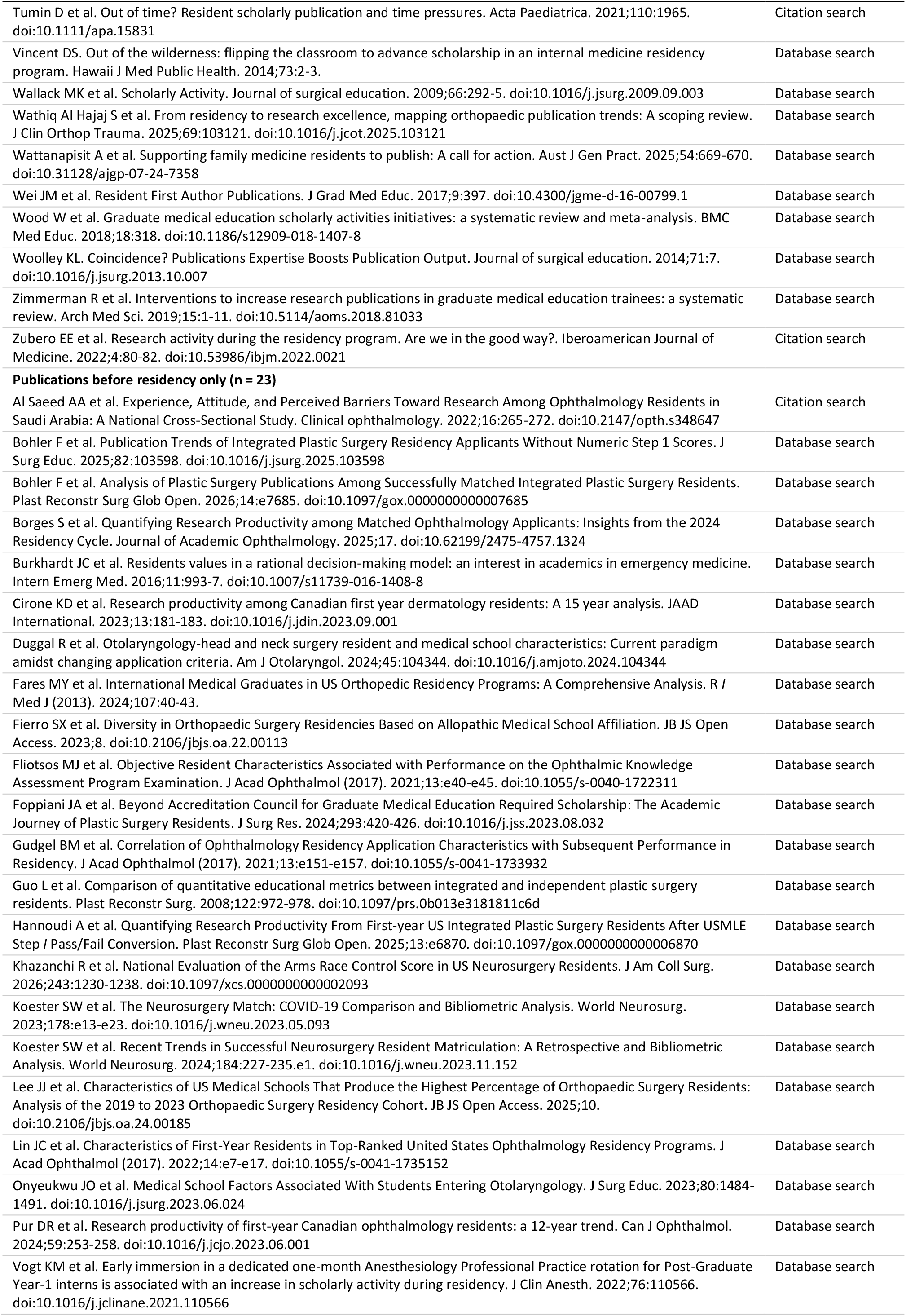
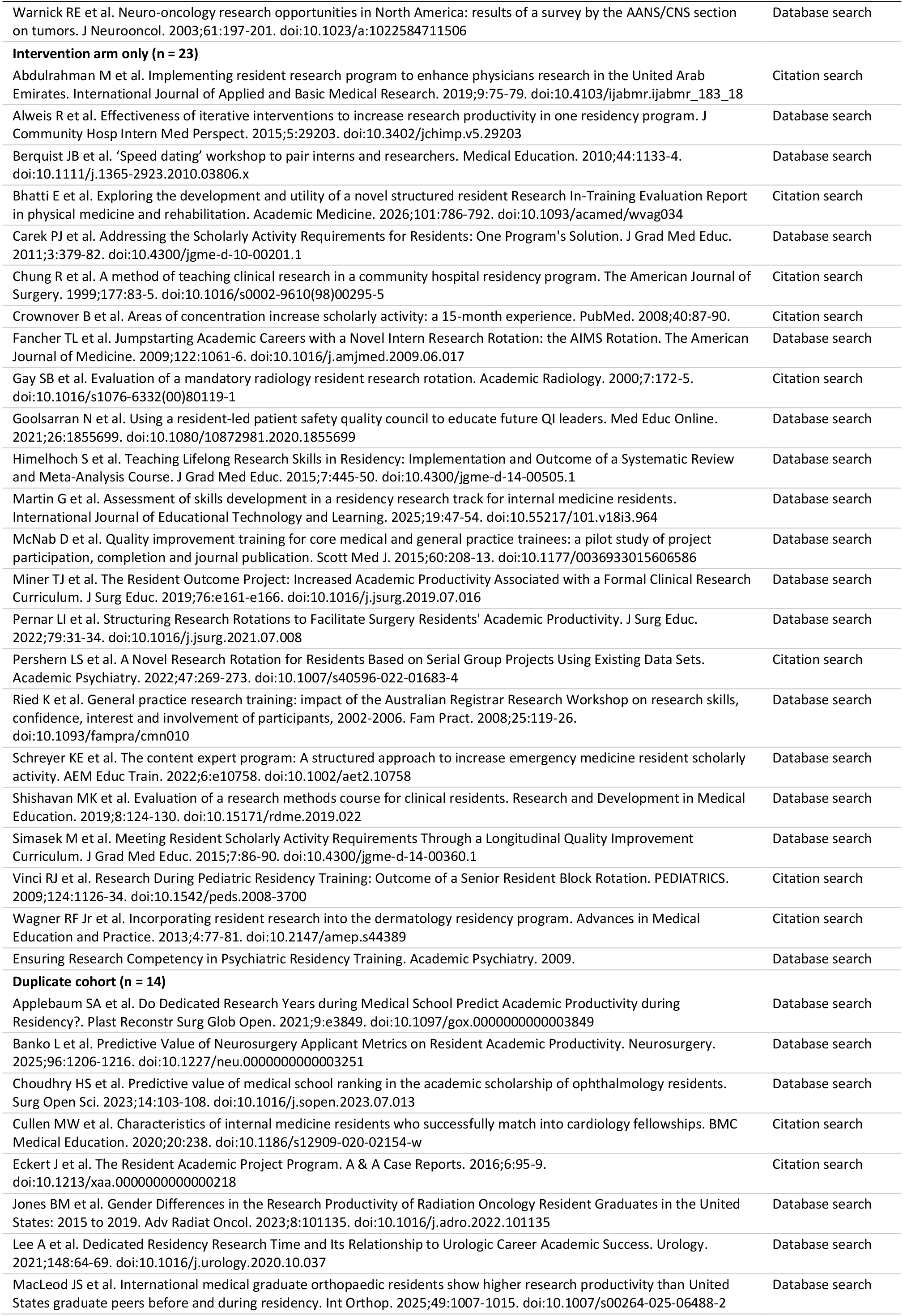
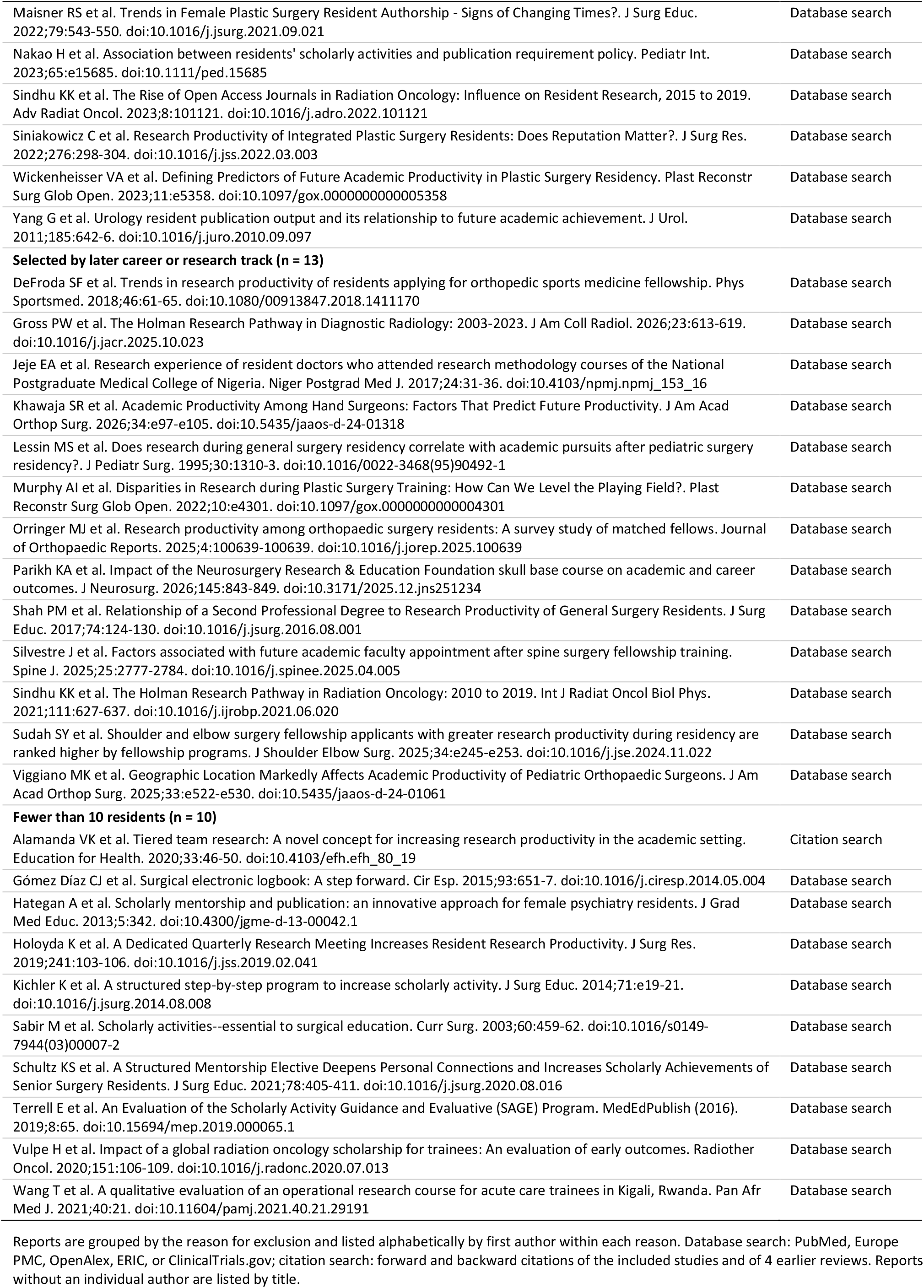
Reports excluded at full-text review, with reasons. All reports excluded after full-text assessment, from database searches and citation searching, with the reason for exclusion.

**Figure S1.**
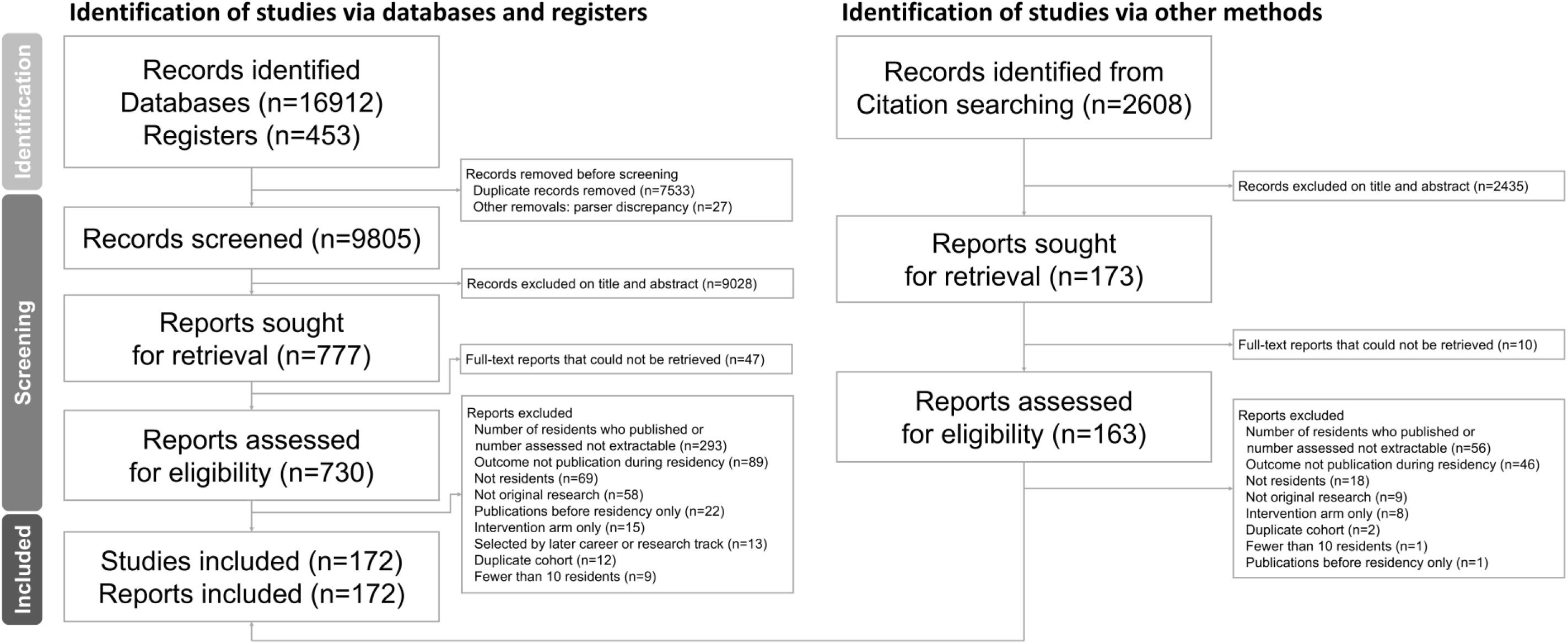
PRISMA 2020 flow diagram. Identification, screening, and inclusion of studies. Records were identified from databases and registers (left) and by citation searching (right). The 27 records removed for other reasons are the difference between the number of PubMed records identified and the number retrieved, which could not be matched to individual records. PRISMA, Preferred Reporting Items for Systematic Reviews and Meta-Analyses.

**Figure S2.**
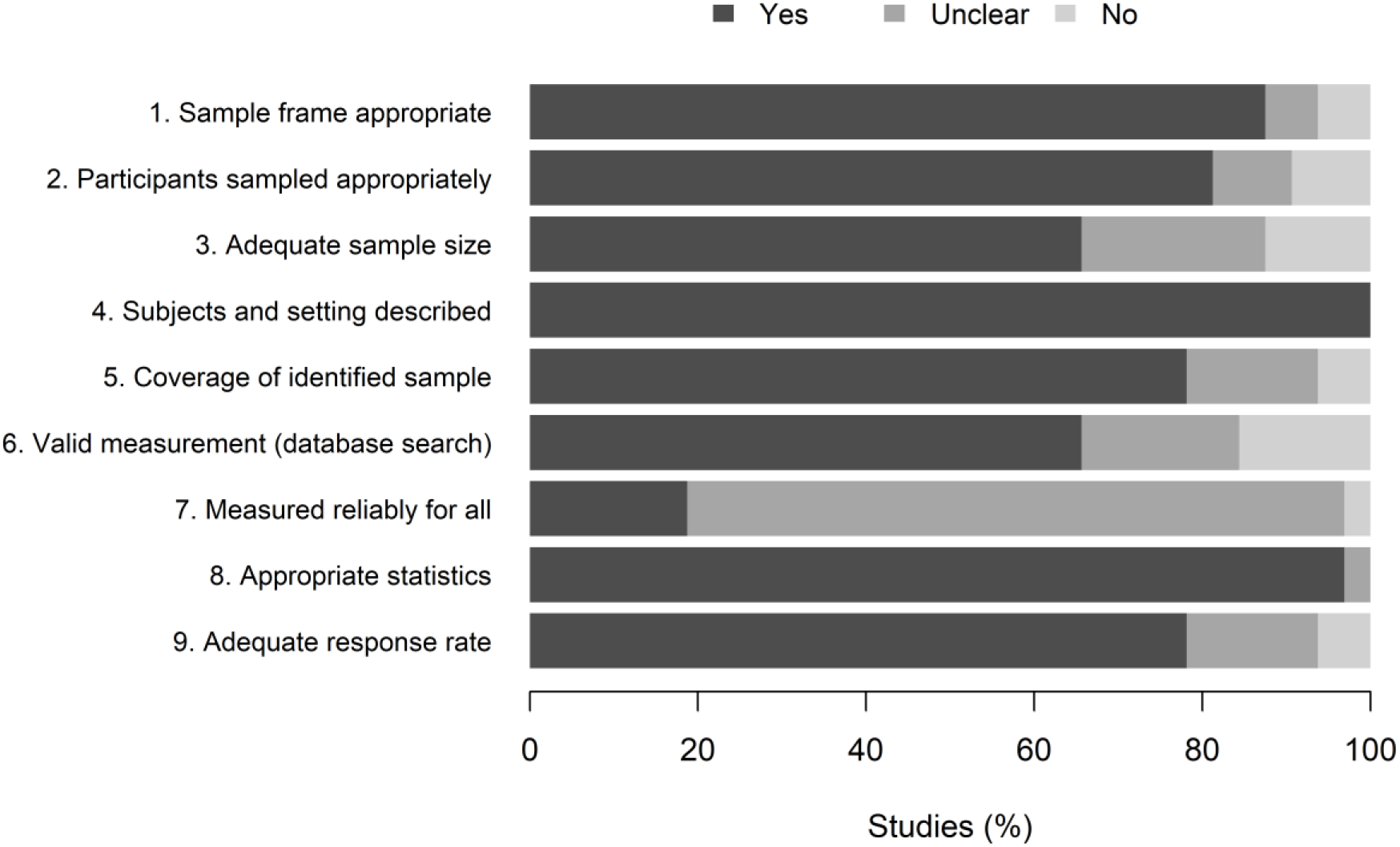
Risk of bias summary (JBI checklist for prevalence studies). Risk of bias in the 32 US studies of the primary analysis, assessed with the Joanna Briggs Institute (JBI) checklist for prevalence studies, adapted to emphasize the denominator, cohort completeness, coverage of the training window, and verification of authorship. Bars show the percentage of studies rated yes, unclear, or no for each of the 9 items. Ratings were made independently by the 2 AI screening agents with the source passage for each item; disagreements were resolved against the source and confirmed by I.-C.T.; ratings for each study are given in Table S7.

**Figure S3.**
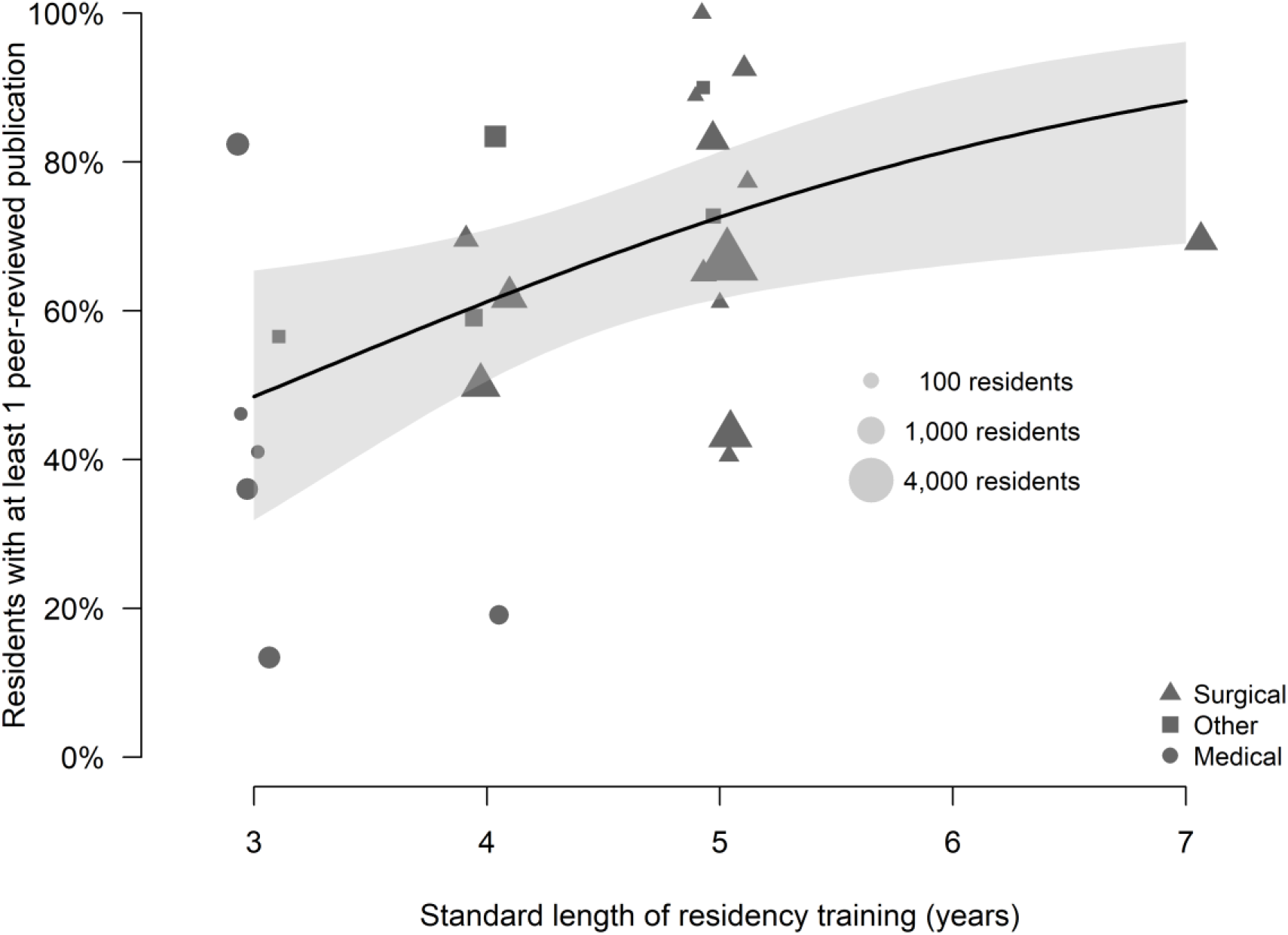
Proportion of residents with at least 1 publication by standard length of residency training. 26 US studies, excluding pre-intervention cohorts. Each marker is a study; marker area increases with the number of residents (18 to 4,320; reference sizes shown), and marker shape shows the specialty group. Markers are offset horizontally to reduce overlap. The line shows the meta-regression on training length (OR per year, 1.68; 95% CI, 1.10–2.57), and the shaded band its 95% CI. Estimates are similar from 3 to 4 years and higher at 5 years. Training length is fixed by specialty, so the 2 cannot be separated in these data.

**Figure S4.**
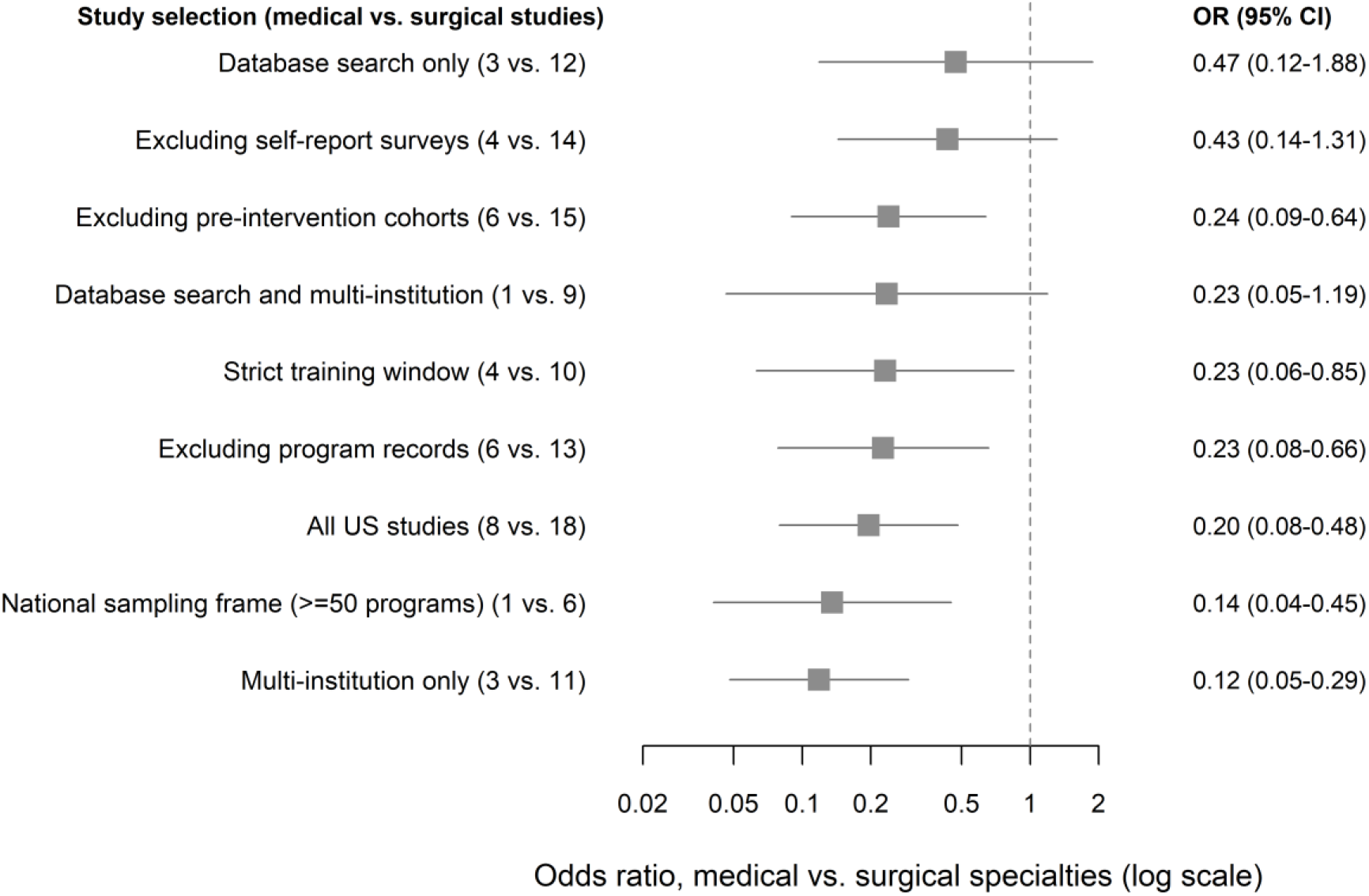
Medical vs. surgical odds ratio under study selections. Odds ratio (OR) for publication in medical vs. surgical specialties under 9 nested study selections of US studies (numbers of medical vs. surgical studies in parentheses). Squares show ORs and horizontal lines their 95% CIs, from mixed-effects logistic meta-regression with specialty as the only moderator; log scale. The dashed line marks no difference. Every selection except all US studies excludes pre-intervention cohorts. Rows are sorted by OR, with the selection in which medical specialties came closest to surgical specialties at the top. ORs below 1 mean lower odds of publication in medical specialties.

**Figure S5.**
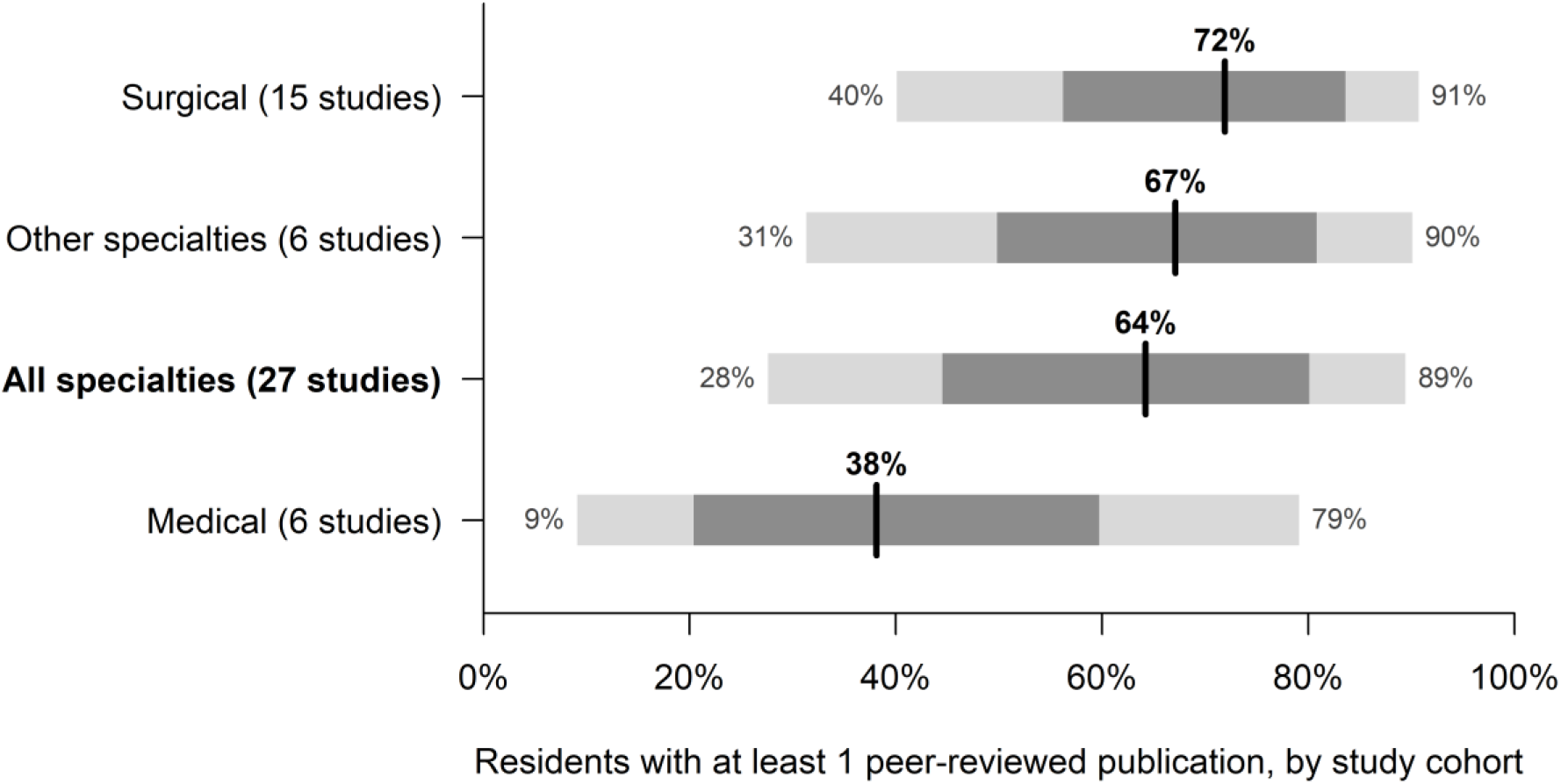
Distribution of published US cohort estimates. Distribution of published US study estimates of the proportion of residents with at least 1 peer-reviewed publication, excluding pre-intervention cohorts. Rows are sorted by median. Bars show percentiles of the predictive distribution from random-effects models, using a t distribution: the light gray bar spans the 10th to 90th percentiles, the dark gray bar the 25th to 75th percentiles, and the black line the median. The distribution describes study cohorts, many of which pooled several programs; it does not rank individual programs. Numbers of studies are given in parentheses.

**Figure S6.**
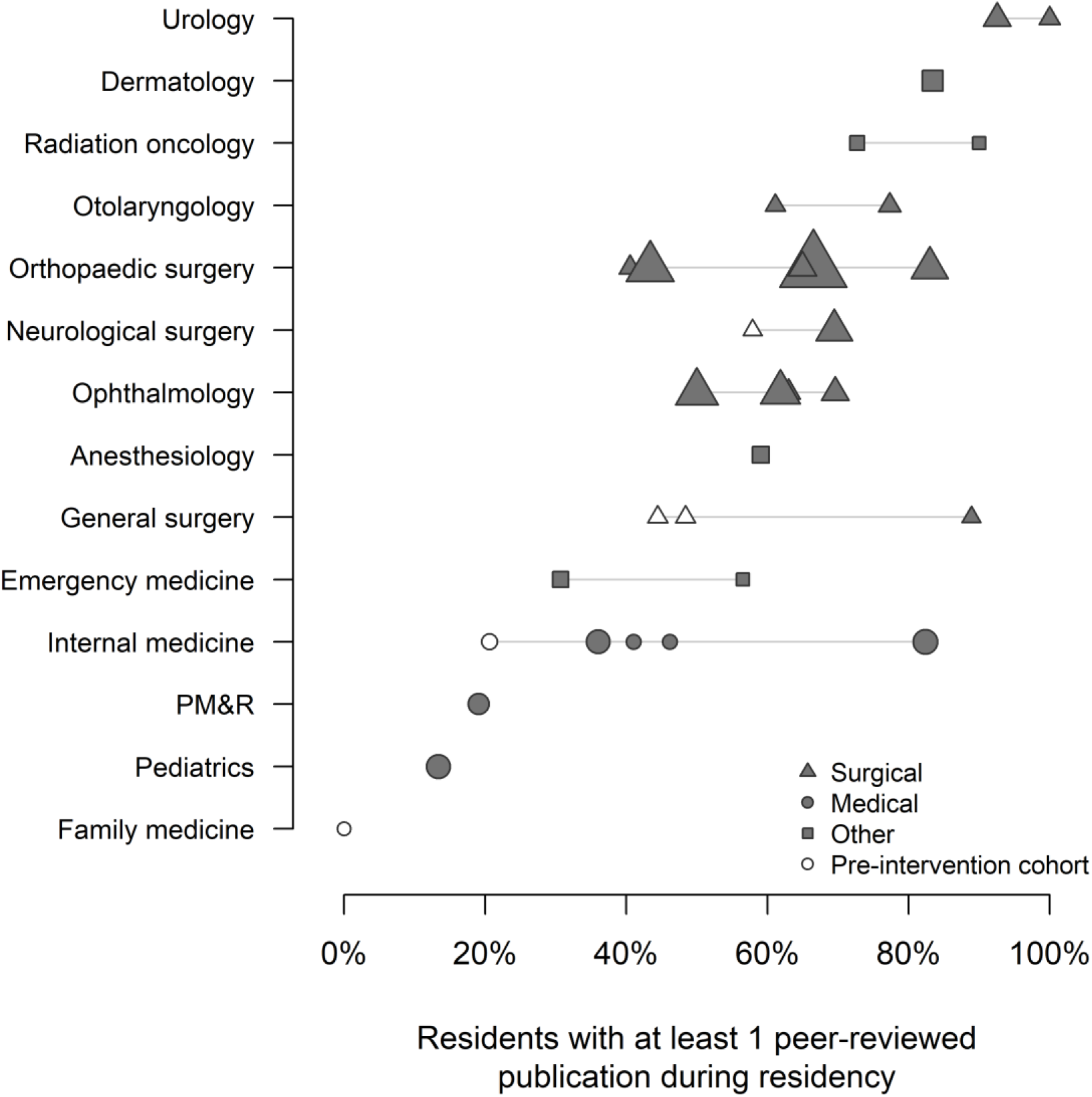
Proportion of residents with at least 1 publication by specialty, US studies. The 32 US studies of the primary analysis, one row per specialty, sorted by the median of its studies. Each marker is a study; marker area increases with the number of residents, marker shape shows the specialty group, and open markers are single-program pre-intervention cohorts. Lines join the studies of a specialty. Many specialties have only 1 or 2 studies, so the figure is descriptive and specialties were not pooled separately. PM&R, physical medicine and rehabilitation.

